# Disentangling phenotype scale, amplification, and effect heterogeneity underlying polygenic GxE architecture of complex traits

**DOI:** 10.64898/2026.10.15.26365681

**Authors:** Zhengtong Liu, Aakarsh Anand, Xinyue Qie, Manuela Costantino, Moonseong Jeong, Aditya Gorla, Noah Zaitlen, Andy Dahl, Sriram Sankararaman

## Abstract

While polygenic gene-by-environment (G×E) signals are increasingly detected in biobank data, it is often unclear whether they reflect the phenotype’s measurement scale or one of two biologically distinct mechanisms: heterogeneous genetic effects across environmental strata (*effect heterogeneity*) or systematically larger but correlated effects in one stratum (*amplification*). We introduce REGENA (Robust Estimator of Gene-ENvironment Architectures), a variance-component method that disentangles these contributions at biobank scale by learning a phenotype transformation that minimizes scale-induced G×E, thereby assessing scale-dependence while jointly estimating the cross-environment genetic correlation and environment-specific genetic variances. In simulations, REGENA is well-calibrated and powerful while recovering the transformation underlying scale-dependent G×E. Applied to 43 quantitative traits and seven environmental exposures in 291,273 UK Biobank individuals, REGENA finds that roughly half of trait-environment pairs with significant G×E are scale-dependent. Among the remaining scale-independent trait-environment pairs, G×E heritability accounted for an average of 15% of additive heritability and was driven primarily by effect heterogeneity (accounting for 71% of G×E heritability on average) although there was substantial variation in its relative contribution across trait-environment pairs. Environment-specific effects tend to make a larger contribution at lower allele frequencies, accounting for 6% of genetic variance in low-frequency variants versus 3.9% at common variants (signed test *p* = 1.7 × 10^−24^). Modeling these distinctions improves environment-specific polygenic prediction (14% on average), with the largest gains for pairs whose genetic effects are least correlated across environments (*p* = 4.0 × 10^−3^; 3.8-fold for testosterone stratified by sex and 2.32-fold for waist-hip ratio stratified by alcohol consumption). Further, environment-stratified GWAS detects 174 loci missed by pooled GWAS with 67 having opposite-signed effects across strata.

## Introduction

Gene-environment (G×E) interactions, the modulation of genetic influences by environmental exposures, have been hypothesized to play an important role in complex traits and diseases^1,2,3^. Analyses of environmental exposures and traits paired with genetic data from large-scale biobanks have led to mounting evidence for G×E, both at the single variant^4,5,6^ and polygenic^7,8,9,10^ levels. This increasing evidence has been accompanied by concerns that these G×E signals may simply reflect the scale on which the phenotype is analyzed^11,12,13,14^. In practice, phenotypes are often transformed to improve adherence to model assumptions but nonlinear transformations can potentially generate spurious interaction signals (wherein an additive phenotype appears to harbor interaction signals due to a non-linear transformation of the phenotype) or can mask scale-specific effects^11,12,15^. This issue is particularly problematic as the appropriate scale is seldom known for most trait-environment pairs analyzed in large-scale studies.

Resolving whether a polygenic G×E signal is scale-dependent is only a first step. The observation of such a signal is compatible with different genetic architectures: genetic effects may differ across environmental strata (effect heterogeneity), or they may be perfectly correlated but systematically larger in one stratum (amplification). Distinguishing between these architectures can reveal whether the environment modifies individual variants or acts on the pathways that aggregate them^16,17^.

Among recent attempts at understanding polygenic G×E, Huang et al. ^14^ proposed an approach to learn a scale transformation of the phenotype based on non-genetic covariates or polygenic scores (PGS) that minimizes scale-dependent heterogeneity. This approach, based on non-genetic covariates or polygenic scores, however, does not provide an unbiased estimation of the underlying genetic variance nor the genetic architecture. On the other hand, Durvasula and Price^16^ proposed an approach to test for distinct genetic architectures underlying G×E by combining disparate tools (such as heritability and genetic correlation estimation and PGS × E regression) and reconciling the results post hoc. Relying on summary statistics, the impact of scale cannot be assessed in their approach. Whether polygenic G×E signals underlying complex traits are robust to phenotype scaling and, if so, how different genetic architectures contribute to polygenic G×E remain open questions.

Here we introduce REGENA (Robust Estimator of Gene-ENvironment Architectures), a statistical model that provides a principled approach to answer each of these questions. REGENA fits a unified linear mixed model that jointly estimates parameters (the genetic correlation across environments and genetic variances and heritabilities within each environment) for a range of transformed phenotypes. These parameters first allow us to identify G×E signals that are invariant to phenotype scaling across monotone transformations of the phenotype and then assess whether amplification or effect heterogeneity (or some combination of the two) contribute to those signals. While REGENA can investigate a broad set of phenotypic transformations, we choose the family of Box-Cox transformations^18^ to assess scale dependence based on its simplicity, interpretability, and on theoretical grounds. Searching across transformations typically requires refitting the variance components model, which is computationally infeasible for biobank-scale datasets. REGENA instead uses a randomized method-of-moments (MoM) estimator that makes this process highly efficient. REGENA further supports partitioning of G×E signals by genomic annotations. Beyond estimation of the phenotype scale and genetic architecture, REGENA also computes a model-based G×E polygenic score (using a best linear unbiased predictor or BLUP) that adapts, in a data-driven manner, to the heterogeneity across environmental strata and predicts individual-level phenotype in an environment-specific manner.

In simulations, REGENA effectively removes G×E effects induced by phenotype scaling and is calibrated and well-powered to detect amplification and effect heterogeneity. Applying REGENA to 43 quantitative traits and seven environmental exposures in 291,273 unrelated white British individuals in the UK Biobank, we find that roughly half of significant G×E on the observed scale are attributable to phenotype scaling. For the trait-environment pairs robust to phenotype scaling (scale-independent pairs), REGENA estimates the G×E heritability and the specificity—the fraction of genetic variance that is environment-specific—and partitions each into amplification and effect heterogeneity. We find that the relative contribution of these mechanisms varies across trait-environment pairs, with most G×E effects driven by effect heterogeneity. Moreover, low-frequency variants show greater environment-specific genetic effects. A G×E polygenic score derived from the REGENA model improved prediction over an additive PGS, with larger gains for trait-environment pairs with low cross-environment genetic correlation. Finally, environment-stratified GWAS of scale-independent pairs recovers loci missed by the pooled GWAS, with one third of stratified GWAS showing opposite-signed effects in the two environments. Overall, these results establish REGENA as a unified framework for identifying polygenic G×E that is robust to phenotype scaling, resolving the underlying G×E mechanisms, translating the G×E architecture into improved locus discovery and stratified prediction.

## Results

### Overview of REGENA

REGENA is a variance-component model that jointly characterizes genome-wide genetic effects on a quantitative phenotype across discrete environments. For simplicity, we describe the model for a two-environment setting defined by a binary exposure ***E*** ∈ {0,1}^*N*^, where *N* is the number of individuals. We denote the corresponding environment-indicator vectors by ***E***_*1*_ and **E**_2_. Given a standardized genotype matrix ***X*** for N individuals genotyped at *M* variants and a phenotype vector ***y***, REGENA allows the SNP effects *β*_*1*_ and *β*_*2*_ in the two environments to follow a joint Gaussian distribution with environment-specific genetic variances 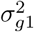 and 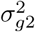 and cross-environment genetic covariance *γ*_*g*_. The corresponding linear mixed model is

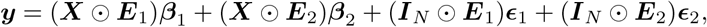

where ⊙ denotes row-wise multiplication. The environment-specific residual effects are ϵ_1_ and ϵ_2_ with variances 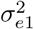 and 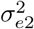 respectively. These variance components induce the cross-environment genetic correlation 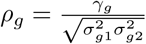 The full covariance decomposition and variance-component estimator are described in the Methods: “Environment-specific linear mixed model” and “Estimation of variance components”. The parameters 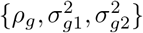 then summarize the architecture of G×E and distinguish two main mechanisms underlying polygenic G×E^16^. Effect heterogeneity is characterized by imperfect cross-environment genetic correlation (*ρ*_*g*_ *<* 1). Amplification is characterized by differential genetic variance 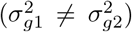. When *ρ*_*g*_ = 1 and 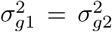, there is no evidence of G×E. Rather than assigning each trait-environment pair exclusively to a discrete category, we describe G×E as a continuous departure from this null along two axes. Effect heterogeneity is quantified by 1 − *ρ*_*g*_, whereas amplification is quantified by the genetic-variance ratio 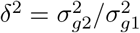. Effect heterogeneity shifts a trait-environment pair along the genetic-correlation axis, whereas amplification shifts it along the genetic-variance axis.

Because apparent G×E can depend on the scale on which a phenotype is analyzed, we first determine whether a detected G×E signal is scale-independent before interpreting its underlying mechanism. To do so, we scan a family of candidate monotone phenotype transformations and re-estimate the model parameters on each transformed scale. A naive implementation would require fitting the variance-component model separately for every transformed phenotype, which is computationally prohibitive at biobank scale. REGENA extends the previous randomized method-of-moments framework (randomized MoM)^9^ to evaluate hundreds of candidate phenotype scales in parallel across hundreds of thousands of individuals and millions of genetic variants (Methods: “Randomized method-of-moments estimation”).

Although REGENA can accommodate diverse phenotype transformations, in this study we focus on the interpretable one-parameter Box–Cox family {***f***_*λ*_}. Within this family, *λ* = 1 corresponds to the original, untransformed phenotype scale, whereas *λ* = 0 corresponds to a log transformation. Additionally, we mathematically justify the use of this family of transformations due to its ability to remove apparent amplification (Notes S3). For each transformed phenotype indexed by *λ*, we test the null hypotheses *ρ*_*g*_(*λ*) = 1 and 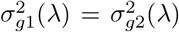. We construct Wald-type statistics for these hypotheses, yielding two-sided *p*-values 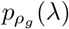 and 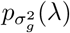, respectively. In practice, we evaluate a grid of *L* candidate values 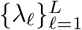. A trait– environment pair is classified as scale-dependent if at least one transformation removes evidence of both effect heterogeneity and amplification.

Among scale-dependent pairs, we select a working transformation that preserves the largest sample-size-weighted genetic variance. For scale-independent pairs, we attempt to select the working transformation nearest to the measured scale at which only one mechanism remains statistically detectable, using the measured scale (*λ* = 1) when no such transformation exists (Figure 1; see Methods: “Scale-independent G×E across transformations”). Finally, REGENA provides environment-specific SNP-effect estimates for a trait-environment pair, enabling the construction of environment-specific polygenic scores. These environment-specific polygenic scores are based on BLUP and use the estimated variance components to adapt to the G×E architecture underlying the trait (see Methods: “Partitioned G×E and polygenic prediction”).

**Figure 1:**
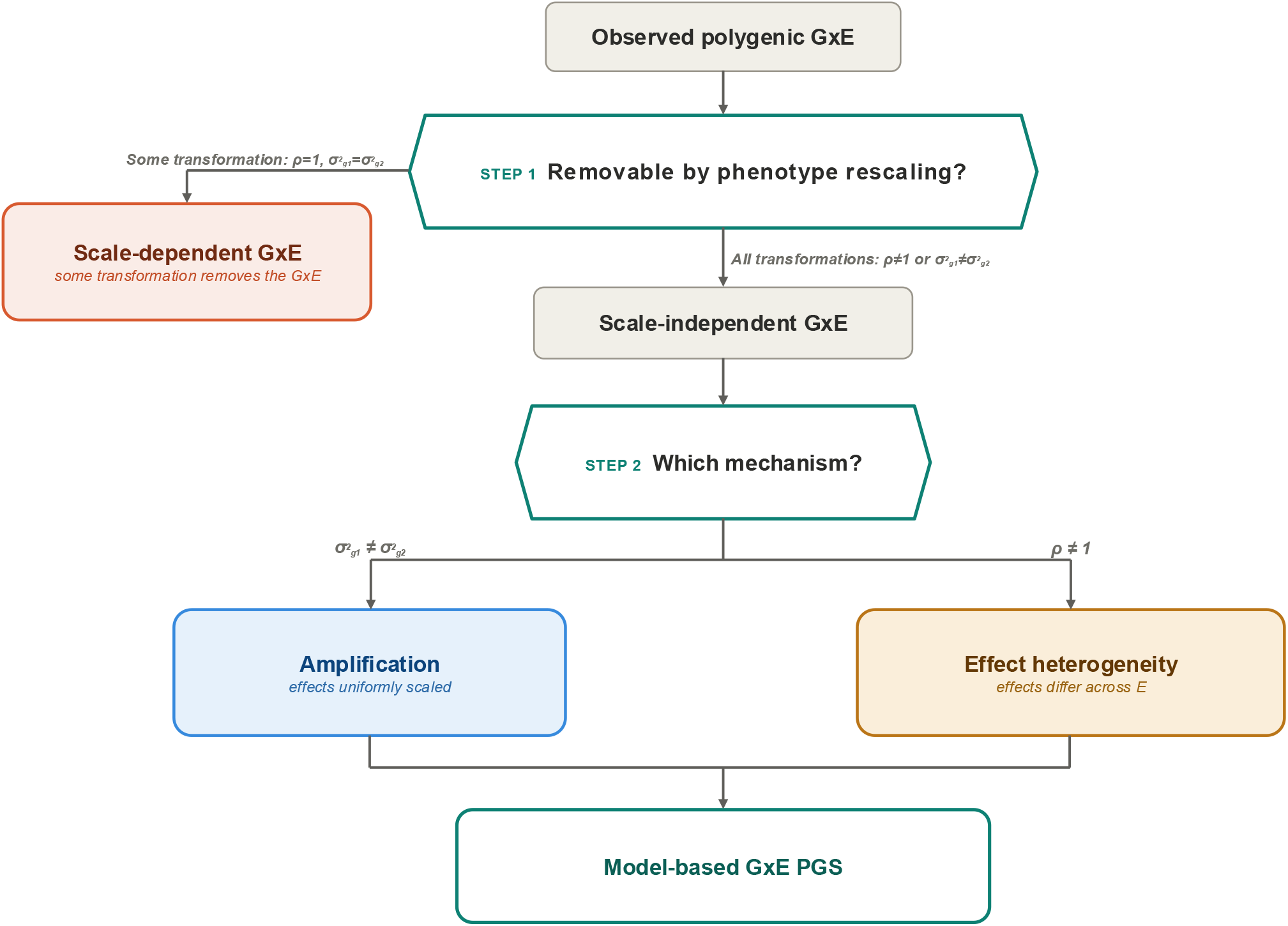
Overview of the REGENA framework. For an observed polygenic G×E signal, REGENA first assesses scale dependence across a set of phenotype transformations. The signal is classified as scaledependent if, for at least one transformation, neither *ρ*_*g*_ = 1 nor 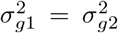 is rejected, consistent with the apparent interaction being removable by a change of phenotype scale. Signals that persist across all transformations are classified as scale-independent and further decomposed by mechanism. Amplification corresponds to genetic effects that are proportional across environments and is therefore indicated by unequal environment-specific genetic variances 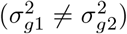. Effect heterogeneity corresponds to environment-specific differences in the relative effects of individual variants and is indicated by an imperfect genetic correlation (*ρ*_***g***_ ≠ 1). The two mechanisms are not mutually exclusive: a signal can show both unequal genetic variances and *ρ*_*g*_ *<* 1. For scale-independent signals, REGENA computes a model-based G×E polygenic score.

### REGENA is calibrated and well-powered in simulations

#### Calibration and power

We assessed the accuracy and calibration of REGENA under the null hypothesis of no G×E by simulating phenotypes with identical genetic variance components across two environmental strata. Unless otherwise specified, we used UK Biobank array SNPs from unrelated White British individuals (*M* = 454,207; *N* = 291,273) and sex as the environmental exposure. We generated 100 replicates for each setting with 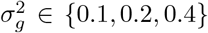. Across settings, REGENA produced accurate estimates of environment-specific genetic variance components and the genetic correlation between strata with controlled false-positive rates (defined as the rejection rate at *α* = 0.05) (Figure 2a,b,c; Figure S2), and calibration was maintained when the environment was phenotypically correlated with the trait, when the environment was heritable and shared causal variants with the trait, under collider bias from an unobserved confounder, and under heavy-tailed residuals (Notes S5; Figure S8).

**Figure 2:**
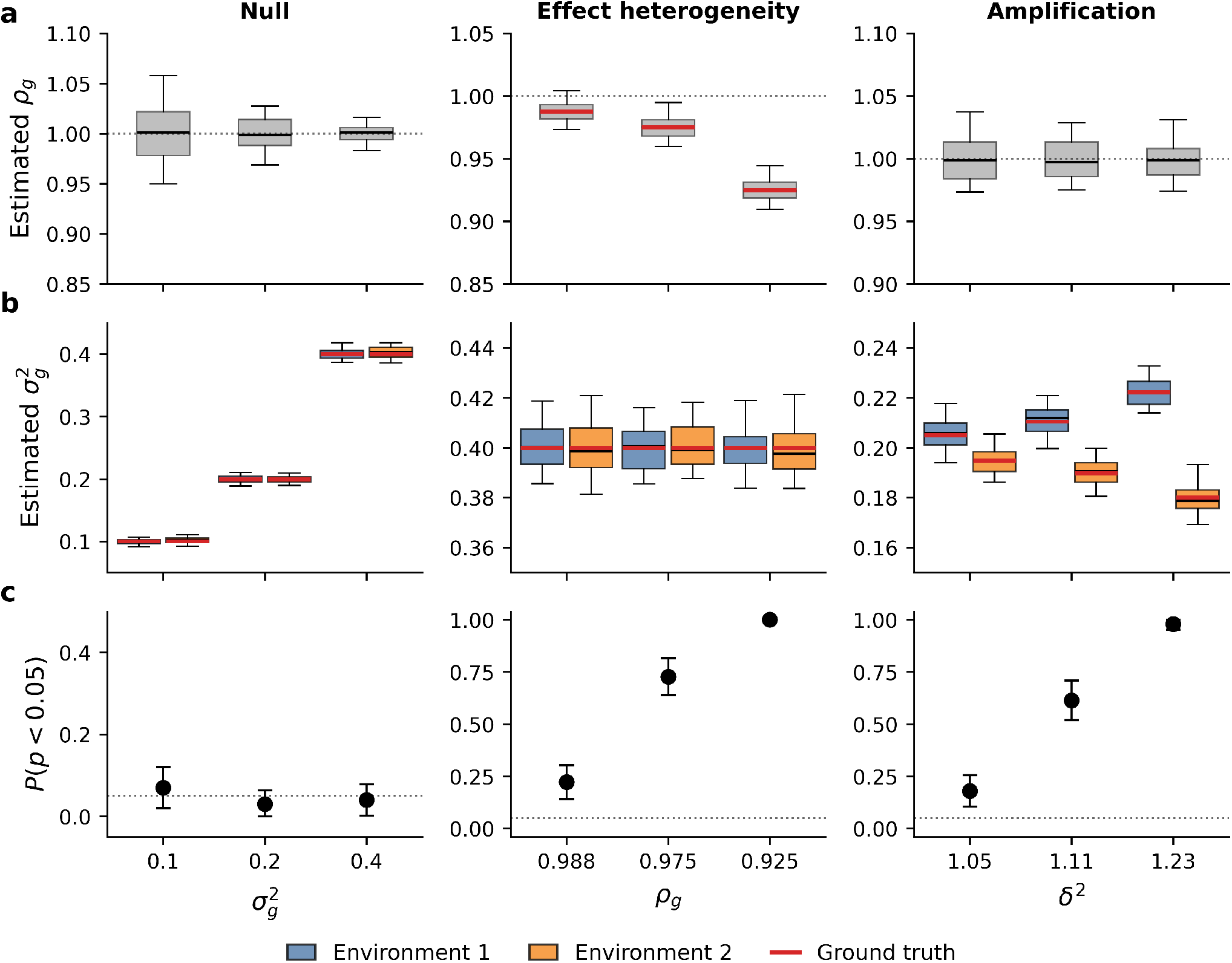
Calibration and power of REGENA in simulation. Phenotypes were simulated using UK Biobank array SNPs from unrelated White British individuals (*M* = 454,207 SNPs, *N* = 291,273 individuals), with sex as the binary environment and 100 replicates per parameter setting. Columns correspond to three simulation regimes: a null model with no G×E and equal environment-specific genetic variances; imperfect cross-environment genetic correlation (effect heterogeneity) with fixed environment-specific genetic variances; and amplification, in which environment-specific genetic variances differ while the genetic correlation is fixed at *ρ*_*g*_ = 1. The x-axis denotes the regime-specific simulation parameter: genetic variance 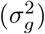 under the null, genetic correlation (*ρ*_*g*_) under effect heterogeneity, and amplification factor (*δ*^2^) under amplification. **a**, Estimated cross-environment genetic correlation, 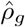. The dotted line denotes *ρ*_*g*_ = 1. **b**, Estimated environment-specific genetic variances, 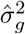, for environment 1 and environment 2. **c**, Rejection rate of the joint test *H*_***0***_ : *ρ*_*g*_ = 1 and 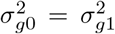 at *α* = 0.05. Points indicate rejection rates across 100 replicates and error bars denote 95% binomial confidence intervals; the dotted line denotes the nominal type I error rate.

We next evaluated REGENA in settings with true G×E by simulating G×E effects arising from either effect heterogeneity or proportional amplification. In the effect heterogeneity setting, we fixed 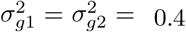 and varied *ρ*_*g*_ from 0.93 to 0.99. In the amplification setting, we fixed *ρ*_*g*_ = 1 and varied the ratio of environment-specific genetic variances, parameterized by the amplification factor *δ*^2^, which ranged from 1.05 to 1.23. REGENA achieved high power even for relatively mild effect heterogeneity and amplification effects (*ρ*_*g*_ *≤* 0.98 or *δ*^2^ *≥* 1.11; Figure 2). Further, it produced accurate parameter estimates in these settings. We also demonstrated that REGENA generalizes GENIE to accommodate G×E effects arising from proportional amplification (Notes S6; Figure S9).

#### Detection of scale-dependent G×E

We next evaluated whether REGENA distinguishes G×E induced by phenotype scale from scale-independent G×E. In null simulations with matched genetic variances across strata, 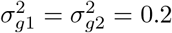, we applied inverse Box–Cox transformations with *λ* ∈ {−1, −0.5, 0, 0.5} to generate phenotypes in which G×E was absent on the generating scale but could appear after transformation. Across simulations, REGENA detected scale-induced G×E for 11% of transformed phenotypes on the observed scale and identified a transformation under which the signal was removed according to the pipeline (Figure 3).

**Figure 3:**
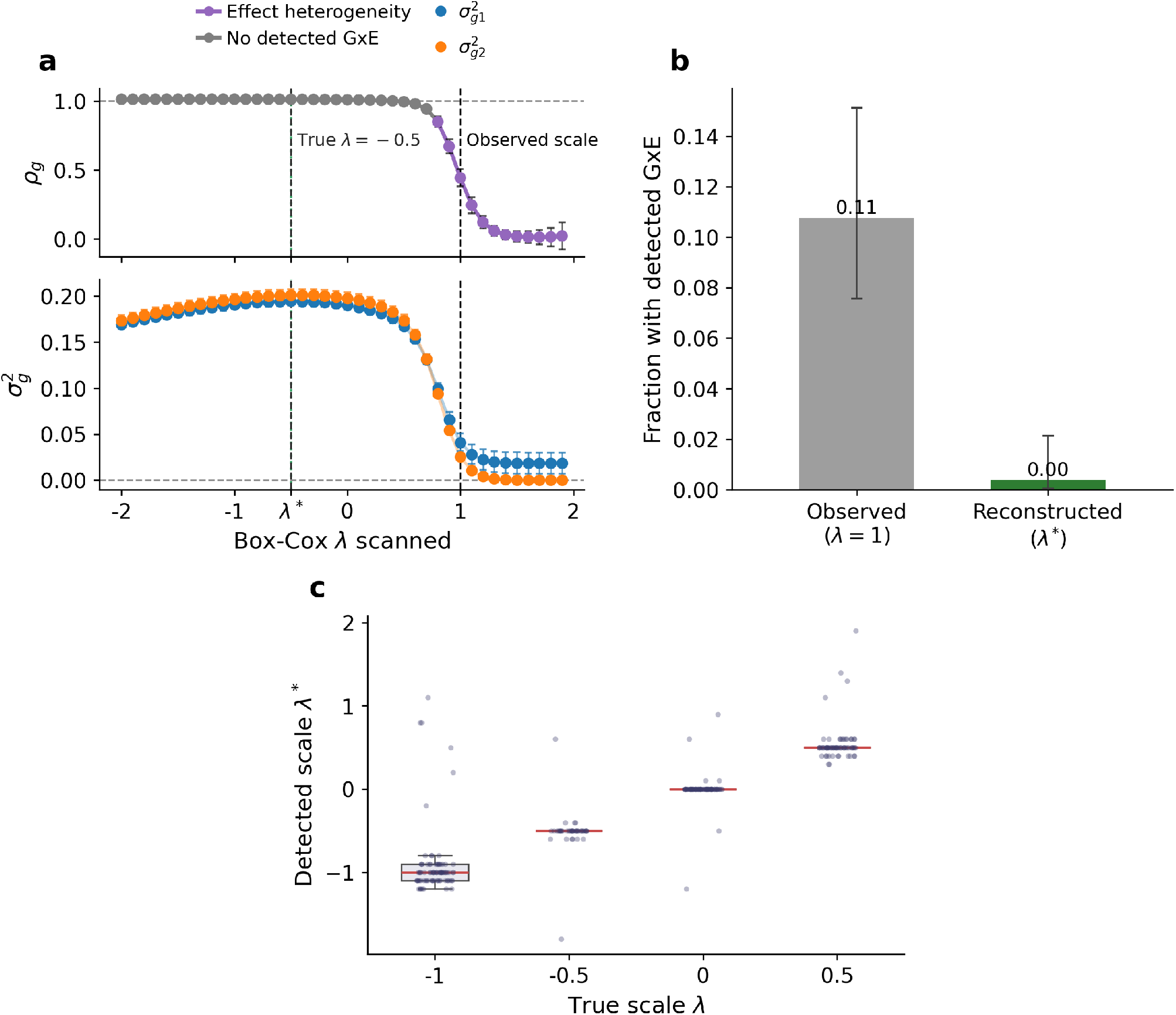
REGENA identifies scale-dependent G×E and recovers the underlying phenotype scale. Phenotypes were simulated on a working scale with equal environment-specific genetic variances,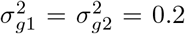, and perfect cross-environment genetic correlation, *α*_*g*_ = 1, and were then mapped to the observed scale using inverse Box-Cox transformations with λ ∈ {−1, −0.5, 0,0.5}. REGENA scanned BoxCox transformations over λ ∈ [−2, 2] and selected a detected scale, 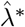, for each replicate. **a**, Representative scan for a phenotype generated using an inverse Box-Cox transformation with λ = −0.5. The estimated genetic correlation, *α*_*g*_, and environment-specific genetic variances, 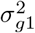 and 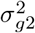, are shown across the scanned transformations. Vertical lines indicate the generating scale, the observed scale (λ = 1) and the detected scale 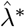. **b**, Fraction of 100 simulation replicates with detected G×E at the observed scale and at the reconstructed scale for phenotypes generated with λ = −0.5. G×E was tested at a nominal threshold of *p <* 0.05; error bars denote 95% Wilson binomial confidence intervals. **c**, Distribution of 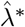 across 100 replicates for each generating value of A.

Since REGENA is based on the randomized MoM method, we evaluated the accuracy and stability of the estimated Box–Cox parameter; the estimates closely tracked the simulated values and remained consistent across random seeds (Figure S3). Importantly, this behavior was not limited to generative transformations within the Box–Cox family. We simulated phenotypes under six nonlinear maps (Supplementary Notes S7; Figure S4) in which no G×E was present before the transformation, but an environment-specific shift induced apparent amplification-type G×E on the observed scale. Although *λ*\* is not directly interpretable under model misspecification, the Box–Cox scan identified a working scale that removed the apparent signal in every case (Figure S5 and S6). The scan is not universal: under strong saturation, *λ*\* was pinned to the scan boundary and the signal persisted (Figure S7). Together, these results show that REGENA identifies and removes a broad class of scale-dependent G×E.

#### Computational scalability

We benchmarked REGENA using UK Biobank array SNPs (*M* = 454,207) varying the sample size and number of phenotypes across ten replicates per configuration. Runtime scaled approximately proportionally with sample size (Figure S1a) and with the number of phenotypes (analyzing as many as 500 phenotypes took about 7 hours), reflecting a fixed genotype-streaming cost and a marginal cost of only approximately 45 seconds per additional phenotype (Figure S1b). Thus, increasing the number of phenotypes fivefold, from *L* = 10 to *L* = 50, increased runtime by only 1.6-fold, enabling phenome-wide G×E scans at biobank scale.

### Half of UK Biobank G×E signals are attributable to phenotype scale

We applied REGENA to 43 quantitative traits across seven binary environments in the UK Biobank (see “UK Biobank data” in Methods for details). For each trait–environment pair, we first tested for G×E on the original phenotype scale. For pairs with significant original-scale G×E, REGENA then assessed whether the signal was scale-dependent by scanning Box–Cox transformations, and classified any remaining scale-independent G×E into effect heterogeneity, amplification, or both. We restricted the Box–Cox parameter *λ* to [*−*3, 3], which encompasses a broad range of monotone transformations commonly used for quantitative traits.

We identified 39 trait–environment pairs with significant G×E on the original scale (*p<* 0.05*/*(43 × 7)) with 29/39 showing evidence of effect heterogeneity and 25/39 showing evidence of amplification with a significant difference in genetic variance between strata. After running REGENA, 19/39 pairs (48.7%) were no longer significant on at least one transformed scale, indicating that nearly half of the original-scale signals could be explained, at least in part, by phenotype scale. This attenuation was observed for both mechanisms: 15/29 pairs with significant effect heterogeneity and 12/25 pairs with significant amplification lost significance on a transformed scale (Figures 5 and 6; Figures S10 to S14). We find that testosterone stratified by sex shows scale-independent G×E: consistent with its well-established sex-specific genetic architecture ^15,19,20^, the estimated genetic correlation remained near zero across all valid tested transformations (Figure 4a,b).

**Figure 4:**
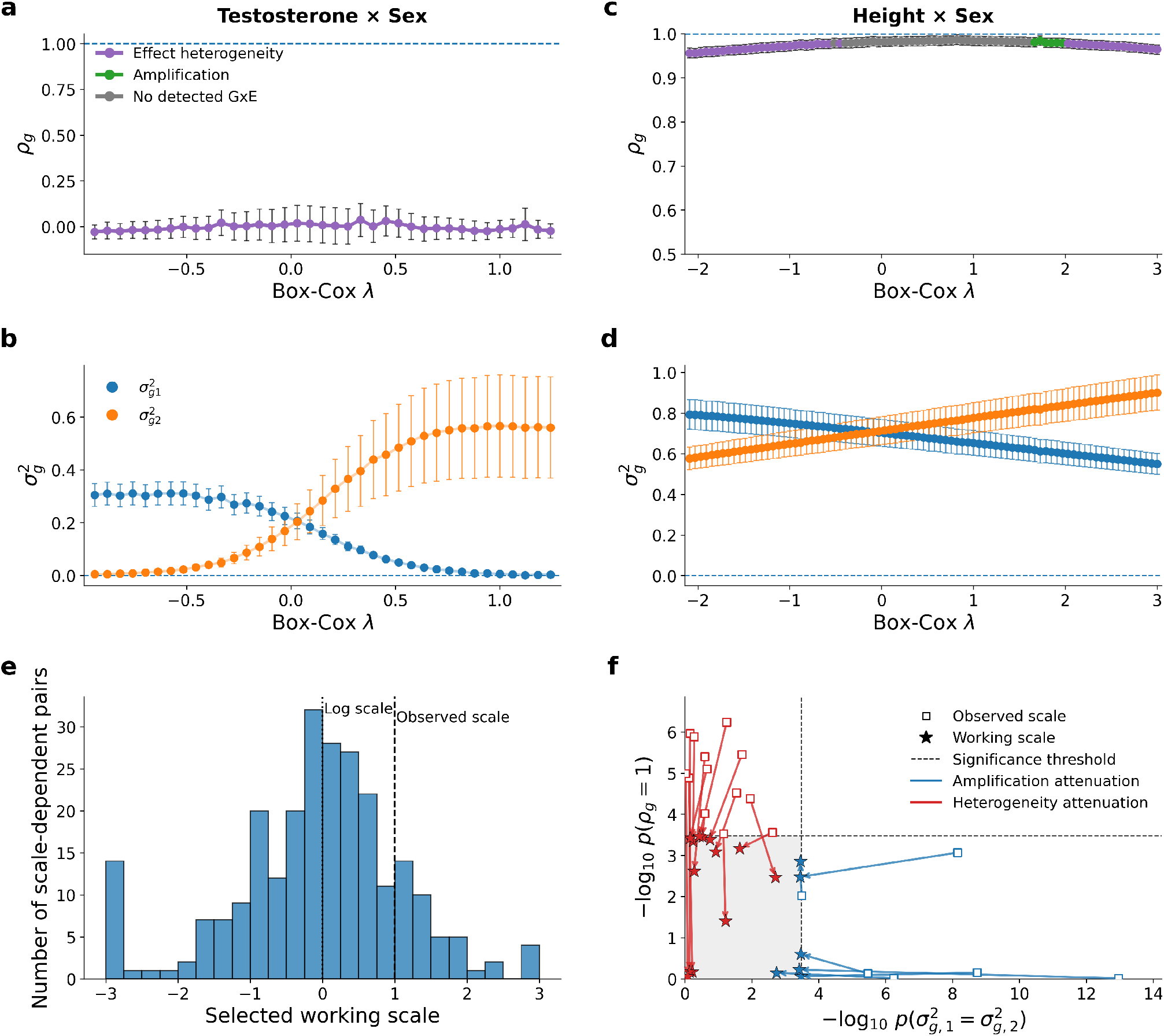
Phenotype transformation distinguishes scale-dependent from scale-independent G×E. **a**, Genetic correlation and **b**, environment-specific genetic variances across valid Box-Cox transformations for testosterone stratified by sex. Colors in the upper panel indicate effect heterogeneity, amplification or no detected G×E. **c**, Genetic correlation and **d**, environment-specific genetic variances across Box-Cox transformations for height stratified by sex. In **a**-**d**, points show parameter estimates and error bars or shaded bands indicate 95% confidence intervals. **e**, Distribution of selected working scales across all trait-environment pairs. The working scale maximized the prevalence-weighted additive genetic variance across the two environmental strata. Selected scales were centered near the log transformation, with mean λ* = −0.08. Dashed lines indicate the log (λ = 0) and observed (λ = 1) scales. **f**, Attenuation of G×E evidence from the observed to the working scale. Each trajectory connects the observed-scale estimate (open square) to the working-scale estimate (star) for a pair with significant observed-scale G×E that was attenuated after transformation. Amplification and effect-heterogeneity evidence are quantified as — 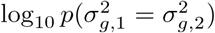 and — log_10_ *p(ρ*_*g*_ = 1), respectively. Dashed lines indicate the significance thresholds, and the shaded lower-left region indicates that neither test is significant. Blue and red trajectories indicate attenuation of amplification and effect heterogeneity, respectively.

**Figure 5:**
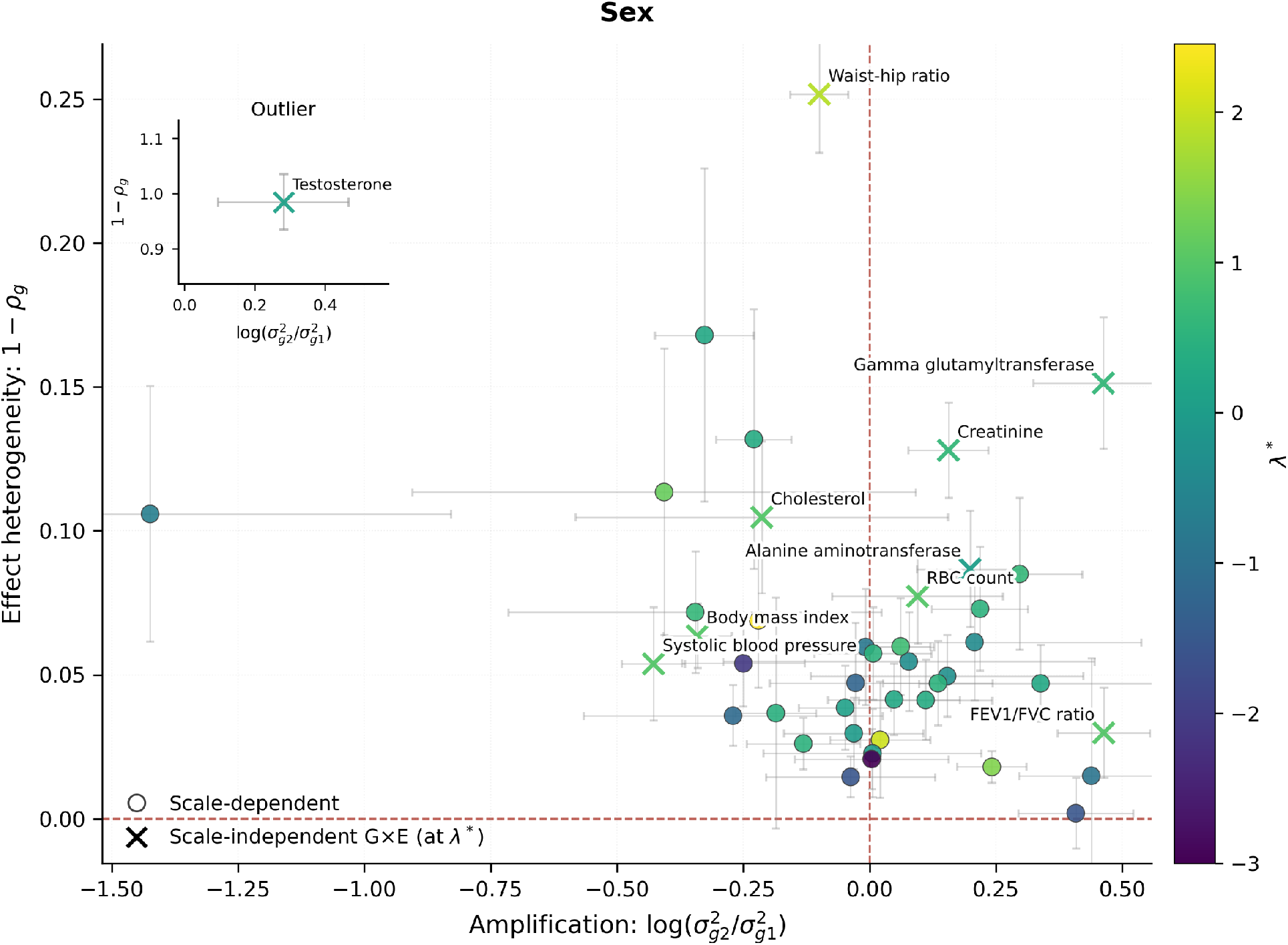
Mechanisms of gene-by-sex interaction across 43 quantitative traits in the UK Biobank. REGENA was applied to each trait-sex pair with a Box-Cox scan over λ ∈ [−3,3]. Each trait is summarized on two axes evaluated at the optimal λ* : effect heterogeneity (1 − *ρ*_*g*_; y-axis) and amplification 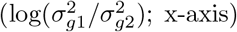. Scale-dependent pairs are shown as dots and scale-independent pairs as markers. Colors indicate the selected λ. Points near the origin indicate weaker G×E, whereas departures along the x- or y-axis indicate amplification- or heterogeneity-type G×E, respectively.

**Figure 6:**
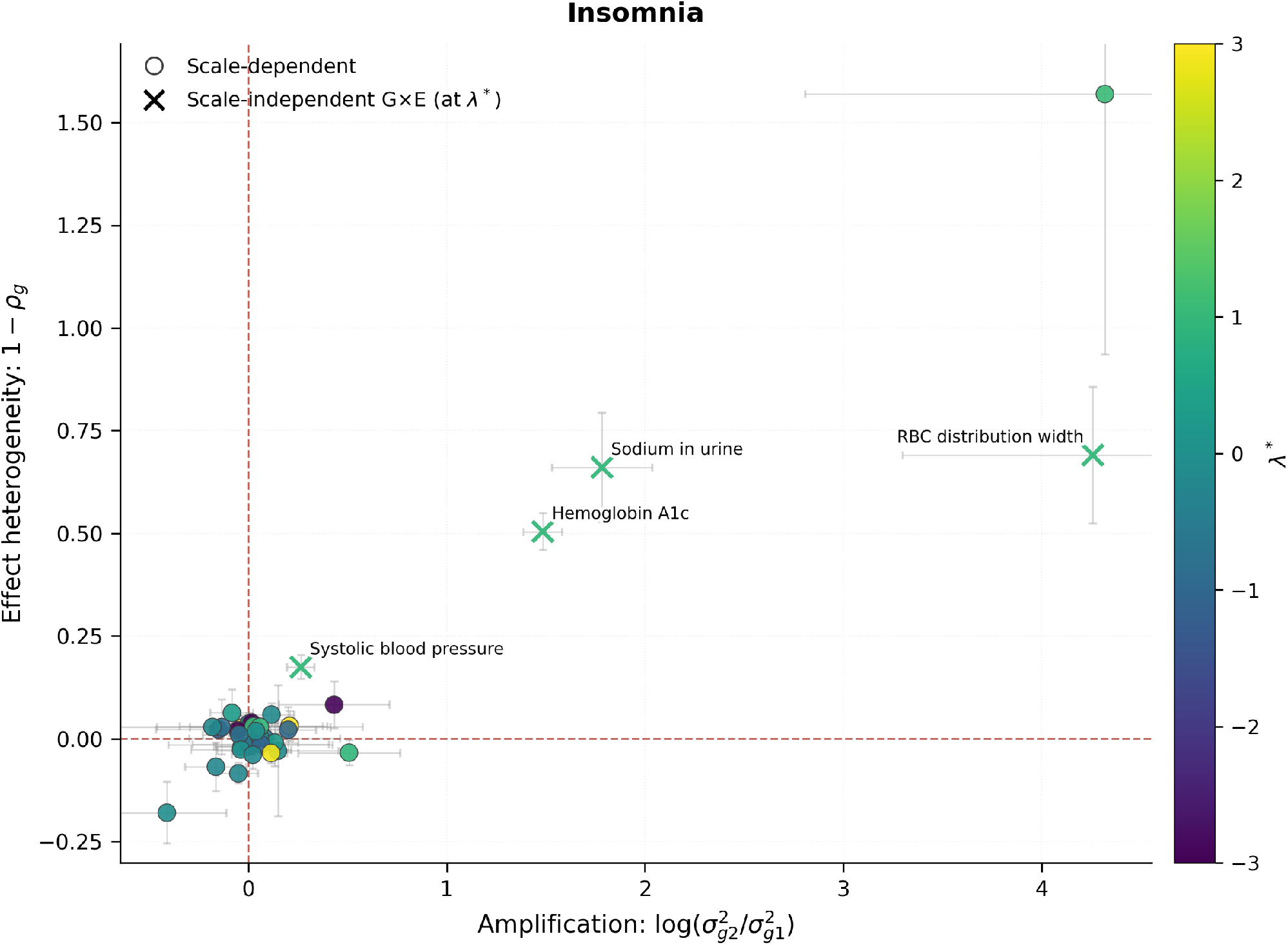
Mechanisms of gene-by-insomnia interaction across 43 quantitative traits in the UK Biobank. REGENA was applied to each trait–insomnia pair with a Box–Cox scan over λ ∈ [−3, 3]. Each trait is summarized on two axes evaluated at the optimal λ*: effect heterogeneity (1 − *ρ*_*g*_; y-axis) and amplification 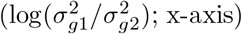. Scale-dependent pairs are shown as dots and scale-independent pairs as markers. Colors indicate the selected λ.

By contrast, for height stratified by sex, the genetic correlation remained close to one across all tested transformations, and the genetic variances did not differ significantly for some scales, providing no evidence of scale-independent G×E overall (Figure 4c,d). The prevalence of scale-independent G×E also varied across environments. Among traits stratified by smoking status, for example, FEV1/FVC was the only trait that retained significant G×E across the tested transformations, indicating that the remaining smoking-associated signals were attributable to phenotype scale.

### Characterizing scale-dependent GxE attenuation

We first examined the working scales for scale-dependent GxE signals. For each trait–environment pair, we defined the working scale as the transformation that maximized the total additive genetic variance. Across 19 scale-dependent GxE traits, we found that the working scale was typically close to the log transformation (mean *λ*\* = *−*0.08; Figure 4e). Although attenuation of GxE on a transformed scale does not imply that the raw-scale signal is a statistical artifact, these results are consistent with the hypothesis that many multiplicative GxE patterns on the observed scale can be represented as additive genetic and environmental effects on an alternative phenotype scale ^11^. We then examine how the transformation affected signals of effect heterogeneity or amplification on the original scale (Figure 4f). Among the 19 trait–environment pairs, 13 showed effect heterogeneity whereas 6 showed amplification that were attenuated on the working scale. For example, glucose stratified by statin use showed amplification-type G×E on the original scale, but the genetic-variance ratio 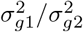 decreased by 4.39-fold after transformation, eliminating evidence for amplification-type G×E and resulting in no significant G×E at *λ*^*\*;*^ = *−*2.40. The largest attenuation of effect heterogeneity was observed for LDL direct stratified by statin use with a transformed scale at *λ*^*\**^ = *−*1.85 removing evidence for scale-independent effect heterogeneity.

### Quantifying the magnitude and mode of G×E heritability

To compare the magnitude of G×E across trait–environment pairs, we quantified total G×E heritability as the variance attributable to environment-specific deviations in genetic effects, denoted 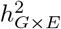 (Notes S8). Scale-independent G×E signals carried substantially greater 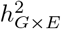 than the scale-dependent pairs. Of the 20 scale-independent pairs, 14 showed significant G×E heritability, with a mean 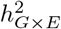 of 2.6%. By comparison, only three of the 19 scale-dependent pairs showed significant G×E heritability on their working scales, with a mean of 0.95%. Among the 14 significant scale-independent pairs, G×E heritability increased total heritability by an average of 15% (Figure 7a). As a sensitivity analysis, we also estimated G×E heritability after inverse-normal transformation of each phenotype. These estimates were broadly concordant with those from the primary analysis (Pearson’s *r* = 0.82 among trait–environment pairs that were marginally significant in both analyses; Figure S15).

**Figure 7:**
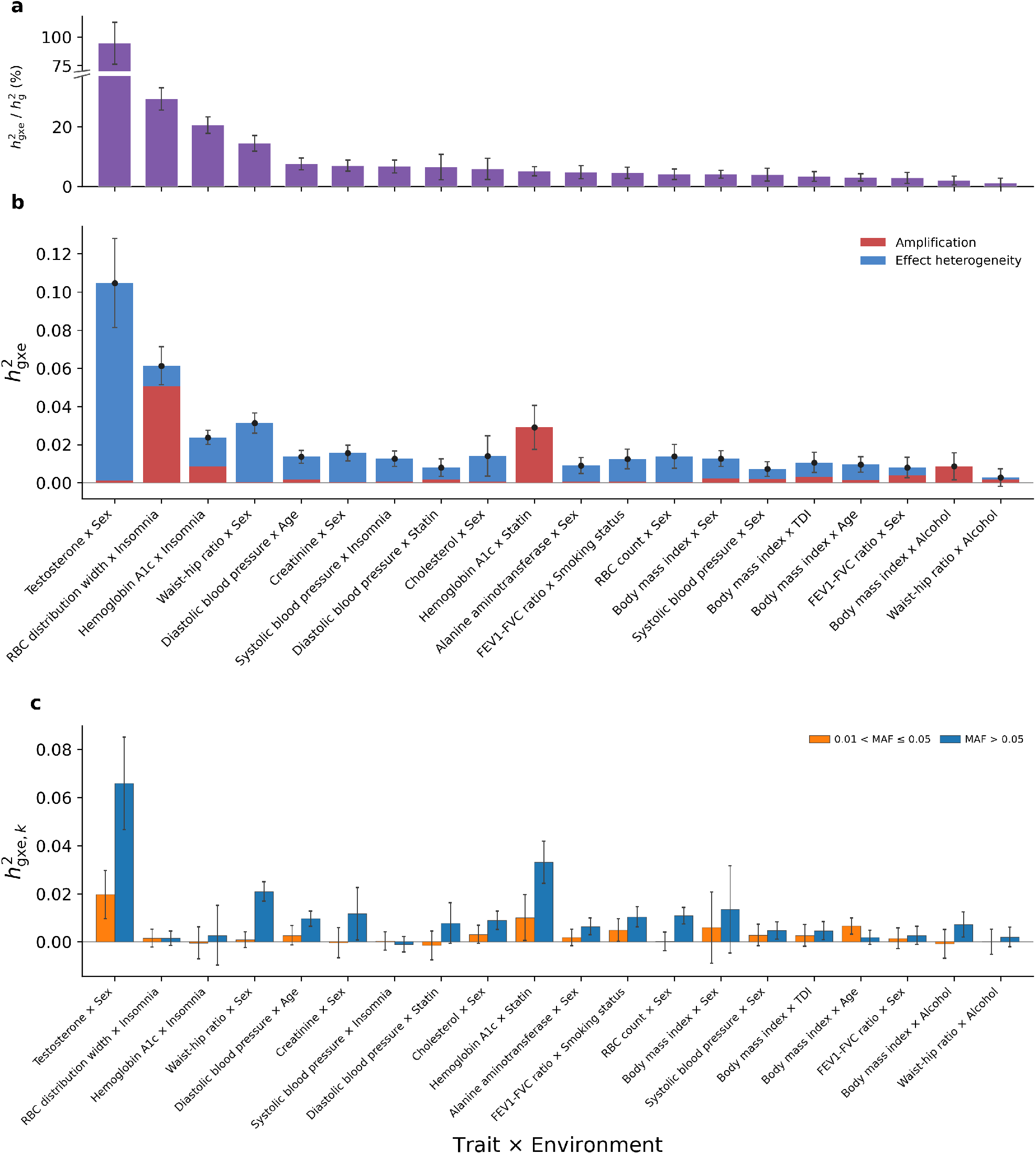
G×E heritability of scale-independent trait-environment pairs. **a**, Relative G×E heritability for the 20 trait-environment pairs with scale-independent G×E, expressed as the increase of total heritability attributable to 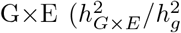 in percentage). Error bars indicate 95% confidence intervals. **b**, Estimated G×E heritability, 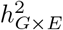, for the 20 scale-independent pairs. Stacked bars partition total G×E heritability into components attributable to amplification and effect heterogeneity. Points indicate total estimates, and error bars indicate 95% confidence intervals. **c**, Estimated partitioned G×E for the scale-independent trait-environment pairs. The genotype is partitioned into common (MAF > 0.05) and low-frequency variants (0.01 < MAF ≤ 0.05).

We further decomposed 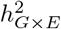 into amplification and effect heterogeneity components (Figure 7b; Notes S8). The relative contributions of these components varied substantially across trait–environment pairs: the amplification proportion ranged from 0.5% to 100%, with a mean of 29%. For HbA1c stratified by statin use and BMI stratified by alcohol consumption, G×E heritability arose almost entirely from amplification (*>* 99%). By contrast, for cholesterol stratified by sex and FEV1/FVC stratified by smoking status, G×E heritability was driven predominantly by effect heterogeneity (*>* 94%).

We also note that apparent amplification can be a consequence of the measurement scale, whereas effect heterogeneity is invariant to monotone transformation (Notes S3). Testosterone stratified by sex, for example, appears to be the most amplification-dominated pair in the set on the original scale (80%), but a mild transformation of the phenotype (*λ*^*\**^ = 0.09) equalizes the sex-specific genetic variances entirely, leaving 99% of G×E heritability as effect heterogeneity, with a cross-sex genetic correlation indistinguishable from zero 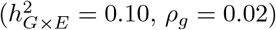.

### Mechanisms of scale-independent G×E

The heritability decomposition suggests that amplification and effect heterogeneity contribute in different proportions across trait–environment pairs. We next examined these mechanisms on the 20 scale-independent trait–environment pairs directly. Among these, 12 showed significant differences in genetic variance while 14 showed cross-environment genetic correlations significantly less than one (*p<* 0.05*/*(2 × 43 × 7)). Overall, genetic effects were substantially shared across environments, with a mean cross-environment genetic correlation of 0.82 and the amplification factor *δ*^2^ = 1.23. Individual pairs differed markedly in whether their G×E signal reflected amplification, imperfect genetic correlation, or both (Figures 5 and 6; Figures S10 to S14; Data S1).

#### Effect heterogeneity

Among trait-environment pairs at the lower end of the genetic-correlation distribution, testosterone stratified by sex displayed a genetic correlation indistinguishable from zero, indicating largely non-overlapping genetic architectures in males and females. Insomnia-stratified traits also showed strong evidence of effect heterogeneity: HbA1c and systolic blood pressure (SBP) had cross-environment genetic correlations ranging from 0.34 to 0.83. The HbA1c–insomnia signal is consistent with prior evidence linking sleep traits to glycemic biology. Mendelian randomization studies have shown that frequent insomnia symptoms causally elevate HbA1c ^21^, and a genome-wide gene-by-sleep-duration meta-analysis identified 16 loci interacting with short or long sleep duration to influence glycemic traits ^22^. Such locus-level effect heterogeneity is consistent with the genome-wide cross-environment genetic correlation of 0.50 *±* 0.04 that we estimate for HbA1c stratified by insomnia.

#### Amplification

At the upper end of the genetic-correlation spectrum, scale-independent G×E was driven primarily by differences in genetic variance, consistent with proportional amplification. HbA1c stratified by statin use showed an approximately five-fold difference in genetic variance (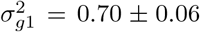 in non-users versus 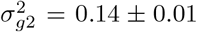 in users; *p* = 3.30 × 10^*−*19^), despite no significant difference in heritability between strata (*p* = 0.14). More broadly, of the 12 scale-independent pairs with significant differences in genetic variance, seven showed no significant difference in heritability and four had cross-environment genetic correlations indistinguishable from one (*p >* 0.05*/*(2 × 43 × 7)). These patterns are consistent with pure proportional amplification, in which the same genetic effects are expressed at different magnitudes across environmental contexts.

#### Mixed mechanisms

Other trait–environment pairs showed both modes of interaction. Diastolic blood pressure (DBP) stratified by age showed evidence for imperfect cross-environment genetic correlation (*ρ*_*g*_ = 0.88*±*0.017; test against *H*_0_ : *ρ*_*g*_ = 1, *p* = 8.7×10^*−*14^), together with unequal genetic variances across younger and older individuals 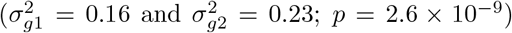 indicating that gene–age interactions operate through both mechanisms.

#### Mechanism varies across exposures within a trait

BMI provides an illustration that the mechanism of G×E can differ across environmental exposures for the same trait. BMI showed imperfect genetic correlation between strata for sex and age (*ρ*_*g*_ *=* 0.94 ± 0.01 and 0.95 ± 0.01, respectively), and differences in genetic variance for age, sex, the Townsend deprivation index, and alcohol consumption. The largest variance difference was observed across alcohol consumption strata 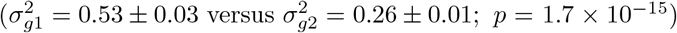. These results indicate that a single trait can exhibit distinct forms of scale-independent G×E depending on the environmental exposure considered.

### Partitioned G×E across MAF and LD

To investigate whether there are systematic differences in polygenic G×E architecture across variant classes, we re-fit REGENA for the 20 scale-independent trait-environment pairs after partitioning variants into eight bins defined by two MAF strata (common, MAF > 0.05; low-frequency, 0.01 < MAF ≤ 0.05) and four LD quartiles within each MAF bin. Total partitioned G×E heritability was highly concordant with genomewide estimates (Pearson’s *r* = 0.95; Figure S17). Common variants accounted for 78% of G×E heritability meta-analyzed across these trait-environment pairs, with the contribution of low-frequency variants ranging from 0.5% to 78% across pairs (Figure 7c; Figure S16). In particular, 77% of the G×E heritability for testosterone stratified by sex was attributed to common variants (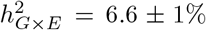 for common versus 2.0 ± 0.5% for low-frequency variants; Figure 7c), whereas heritability for BMI stratified by age was driven by low-frequency variation (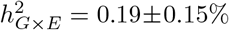 for common versus 0.66±0.17% for low-frequency variants). For variants partitioned by LD quartiles, the highest-LD quartile accounted for 41% of meta-analyzed G×E heritability, compared with 15% for the lowest. Testosterone by sex concentrated 44% of G×E heritability in the highest-LD quartile (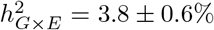 for the variants in highest LD quartile versus 1.1 ± 1.1% in the lowest), whereas BMI by age concentrated 43% in the lowest (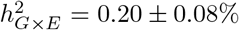 for the variants in highest LD quartile versus 0.37 ± 0.15% in the lowest). The variation of G×E heritability in MAF and LD bins motivates us to investigate the environment-specific effects across annotations in the next section.

### Environment-specific genetic effects are enriched at lower allele frequencies

Motivated by recent work examining the specificity of genetic effects across traits and cell types^23,24^, we hypothesized that genetic effects at lower frequencies are likely to be more environment-specific. To test this hypothesis, we applied REGENA to 7.7 million imputed SNPs (MAF > 0.01%), partitioned into five MAF bins and two LD strata within each MAF bin across all trait-environment pairs. As above, estimates are reported on each pair’s working scale (Algorithm 1). Analogous to measures of the cell-type specificity for eQTLs^24^, we defined G×E specificity as the fraction of genetic variance attributable to environment-specific effects, or equivalently, the ratio of squared per-allele G×E effects to total squared per-allele effects (Notes S9).

Polygenic G×E effects were substantially more specific at low-frequency variants. Meta-analyzed across trait-environment pairs, overall specificity was 6.0% for low-frequency (MAF ≤ 0.05) variants compared with 3.9% for common (MAF > 0.05) variants (sign test, *p* = 1.68 × 10^−24^, *n* = 43×7 = 301). Specificity increased with decreasing minor allele frequency, rising from 3.7% in the most common to 19.3% in the lowest frequency bin (Figure 8a), consistent with stronger selection against highly pleiotropic variants^25,23^. Specificity also varied across environments: for variants with MAF > 0.05, specificity ranged from 1.7% (insomnia) to 7.1% (sex). Notably, the low-frequency specificity was consistently higher than common-variant specificity across all seven environments (Figure 8b).

**Figure 8:**
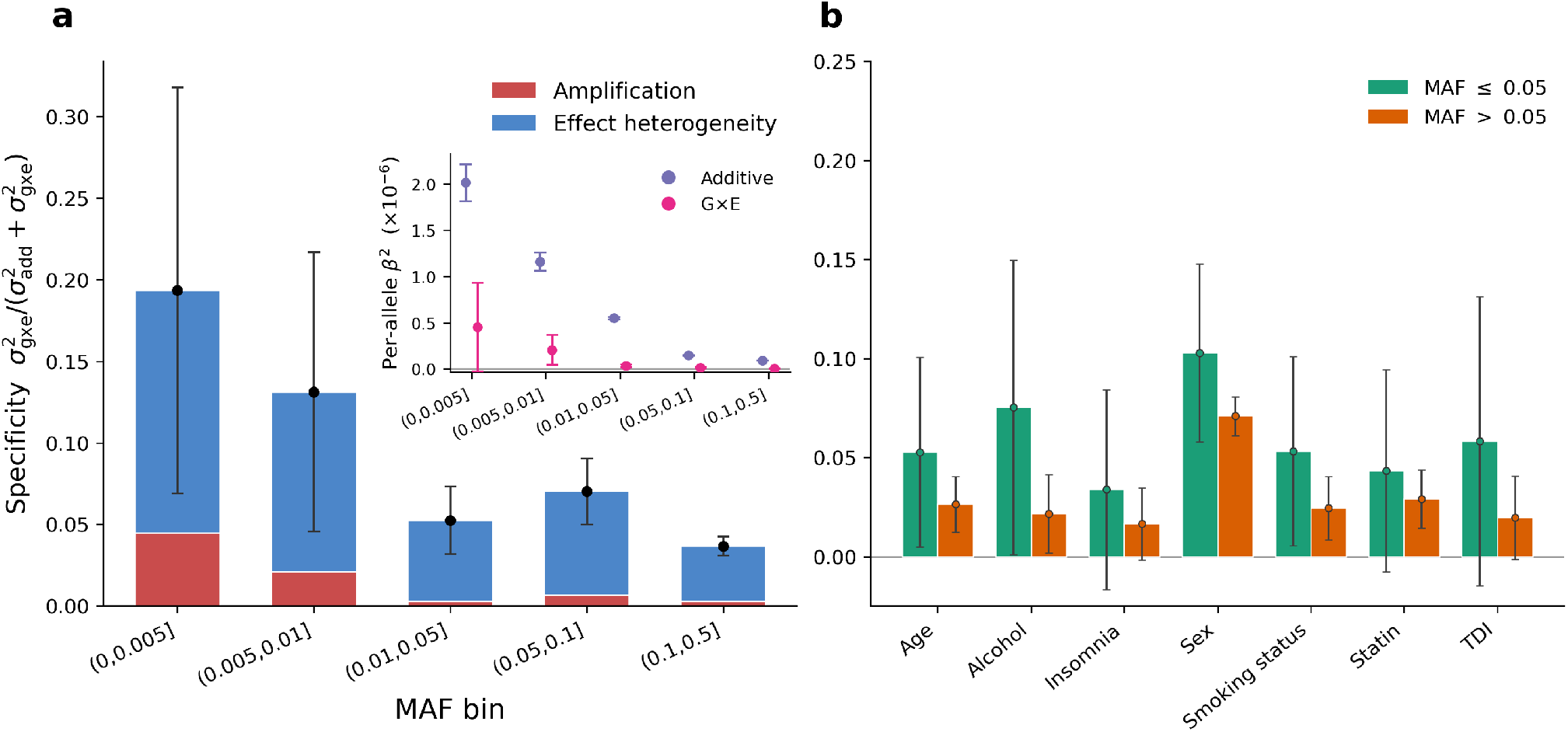
Decomposition of G×E specificity. **a**, MAF-stratified specificity of G×E heritability, metaanalyzed across trait-environment pairs. Specificity was evaluated on the selected working scale for scaleindependent and scale-dependent pairs. Stacked bars show the contributions of amplification specificity, *S*_**amp**_, and effect-heterogeneity specificity, *S*_**het**_. The inset shows MAF-stratified squared per-allele effect sizes for the additive and G×E components. Within each MAF bin, the meta-analyzed additive heritability 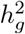 and G×E heritability 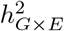 were divided by 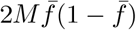, where *M* is the number of variants in the bin and 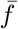 their mean MAF. Points denote the point estimates and error bars indicate 95% confidence intervals in all panels. **b**, Environment-specific G×E specificity meta-analyzed across traits and summarized separately for variants with MAF ≤ 0.05 and MAF > 0.05.

We next decomposed specificity into amplification and effect-heterogeneity components. At the working scale, effect heterogeneity accounted for the majority of specificity in every MAF bin, from 91.8% in the most common bin to 76.8% in the rarest bin. Thus, environment-specific genetic effects arise primarily through imperfect cross-environment genetic correlation across the allele-frequency spectrum. On the observed scale, the amplification component was larger (Figure S18), accounting for roughly half of specificity in the intermediate bins. This component was attenuated by the Box-Cox rescaling, consistent with our previous observation that some of the apparent amplification reflects the measurement scale.

### Modeling G×Eimproves environment-specific polygenic prediction

The model underlying REGENA enables us to construct environment-specific polygenic scores (PGS). To assess whether the architectural distinctions captured by REGENA translate into improved prediction, we compared two scores for each trait-environment pair: a baseline BLUP PGS fit across pooled environments and a G×E PGS with environment-specific BLUP weights derived from REGENA’s stratum-specific genetic variances and cross-environment covariance (Methods, “Partitioned G×E and polygenic prediction”). Both scores were partitioned across the same eight MAF × LD bins on common SNPs, differing only in whether genetic effects were allowed to vary between environments. Individuals were randomly split 60/20/20 into training, calibration, and test sets. We quantified incremental *R*^2^ over a covariates-only model that included the environment and summarized the performance of the G×E score relative to the baseline. Analyses were restricted to the 20 trait-environment pairs showing scale-independent G×E.

On the held-out test set, mean incremental *R*^2^ across the 20 pairs increased from 0.051 for the baseline PGS to 0.058 for the G×E PGS (Δ*R*^2^ = 0.007; 1.14-fold, paired t-test *p* = 0.036) (Figure 9a). Within environment strata, mean incremental *R*^2^ increased from 0.054 to 0.057 (1.06-fold; Figure S19). Specifically, testosterone by sex showed the largest improvement: incremental *R*^2^ increased from 0.010 to 0.040 in males (3.82-fold). Whereas the baseline score showed no detectable predictive value in females (Δ*R*^2^ = −0.001, 95% CI, [−0.003,0.001]); the G×E score instead achieved an incremental *R*^2^ of 0.009 (95% CI, [0.004, 0.014]). For WHR, the G×E score improved prediction by 2.32-fold and 1.89-fold across the two alcohol strata and by 1.62-fold in males (*R*^2^ : 0.022 to 0.037) and 1.51-fold in females (R^2^ : 0.035 to 0.052) for sex. The magnitude of the predictive gain also tracked the degree of genetic divergence between environments: per-stratum improvement increased with 1 − *ρ*_*g*_ (Pearson’s r = 0.44, p = 0.004; Figure 9b).

**Figure 9:**
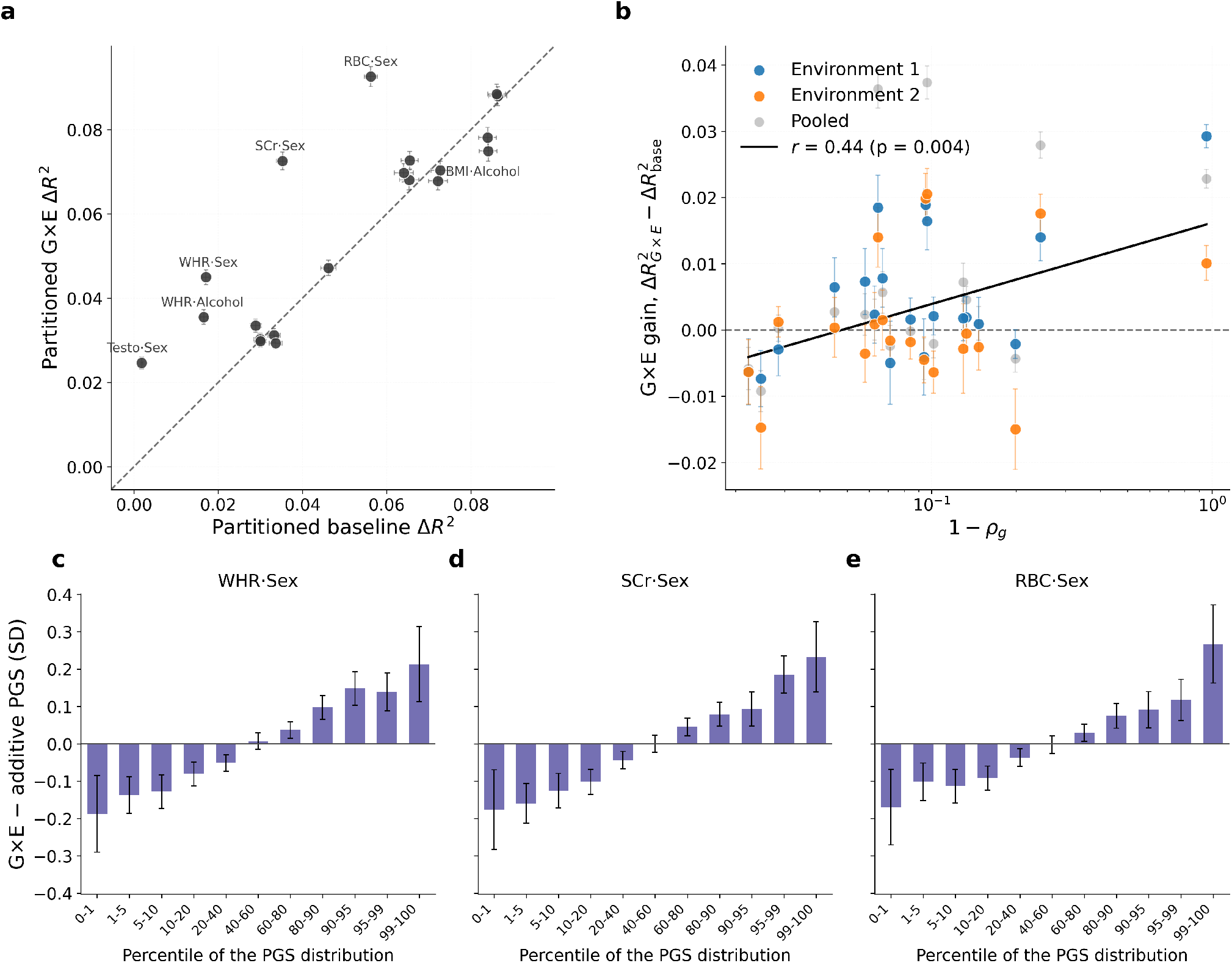
Environment-specific polygenic scores improve prediction for scale-independent G×E. Both panels use the 20 scale-independent trait-environment pairs and compare a MAF/LD-partitioned G×E score with a non-G×E baseline that pools the environmental strata. Incremental *R*^2^ (Δ*R*^2^) is the out-of-sample gain over a covariates-only model (sex, 20 genetic PCs, environment indicator); both scores share the covariate set and the train/test split. The error bar denotes one standard error from a nonparametric bootstrap over held-out individuals. **a**, G×E versus baseline ΔR^2^ on the full held-out set, one point per pair. The dashed line is *y* = *x*; points above it indicate pairs where the G×E score achieves higher ΔR^2^ than the baseline. **b**, G×E gain 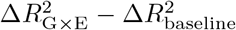 against 1 − *ρ*_*g*_ (log_**10**_ scale). Larger 1 − *ρ*_*g*_ indicates more effect heterogeneity. The solid line denotes the OLS fit (Pearson *r* = 0.44; *p* = 0.004). **c**-**e**, waist-hip ratio by sex, creatinine in urine by sex, and RBC count by sex: held-out individuals were binned by percentile of the additive score and, separately, of the G×E score, and bars give the mean covariate-adjusted phenotype under the G×E score minus that under the additive score in each bin, in phenotype SD units; error bars are 95% confidence intervals within the pair. Panels **c**-**e** share the *y* axis.

The G×E PGS also improved discrimination at the tails of the score distribution. Among the five trait-environment pairs with the largest gains in incremental *R*^2^ over the baseline PGS, the difference in mean covariate-adjusted phenotype between individuals in the top and bottom 1% of the score distribution was 0.35-0.44 SD greater under the G×E PGS than under the baseline PGS: 1.22 to 1.66 SD for red blood cell count by sex, 1.03 to 1.43 SD for creatinine by sex, 0.70 to 1.10 SD for waist-to-hip ratio by sex, 0.66 to 1.01 SD for waist-to-hip ratio by alcohol intake, and 0.27 to 0.68 SD for testosterone by sex (Figure 9c-e; Figure S20; Table S3).

Thus, explicitly modeling cross-environment genetic covariance yields greater gains in polygenic prediction when genetic effects tend to be less correlated across environments.

### Environment-stratified GWAS recovers loci missed by pooling

To determine whether G×E also improves genetic discovery, we performed GWAS in the pooled sample and separately within each environment stratum for the 20 scale-independent trait-environment pairs (Notes S13; Table S2). Stratification identified 174 loci not detected in the pooled analysis (8.7 per pair; *p* < 5 × 10^−8^; Figure 10a), with 67 of 174 showing opposite-sign effects between environments (Figure S21). As a negative control, height stratified by sex (a pair with no significant GxE) yielded only four novel loci: 0.27% of pooled loci, in contrast to the median of 3.0% across the 20 scale-independent pairs.

**Figure 10:**
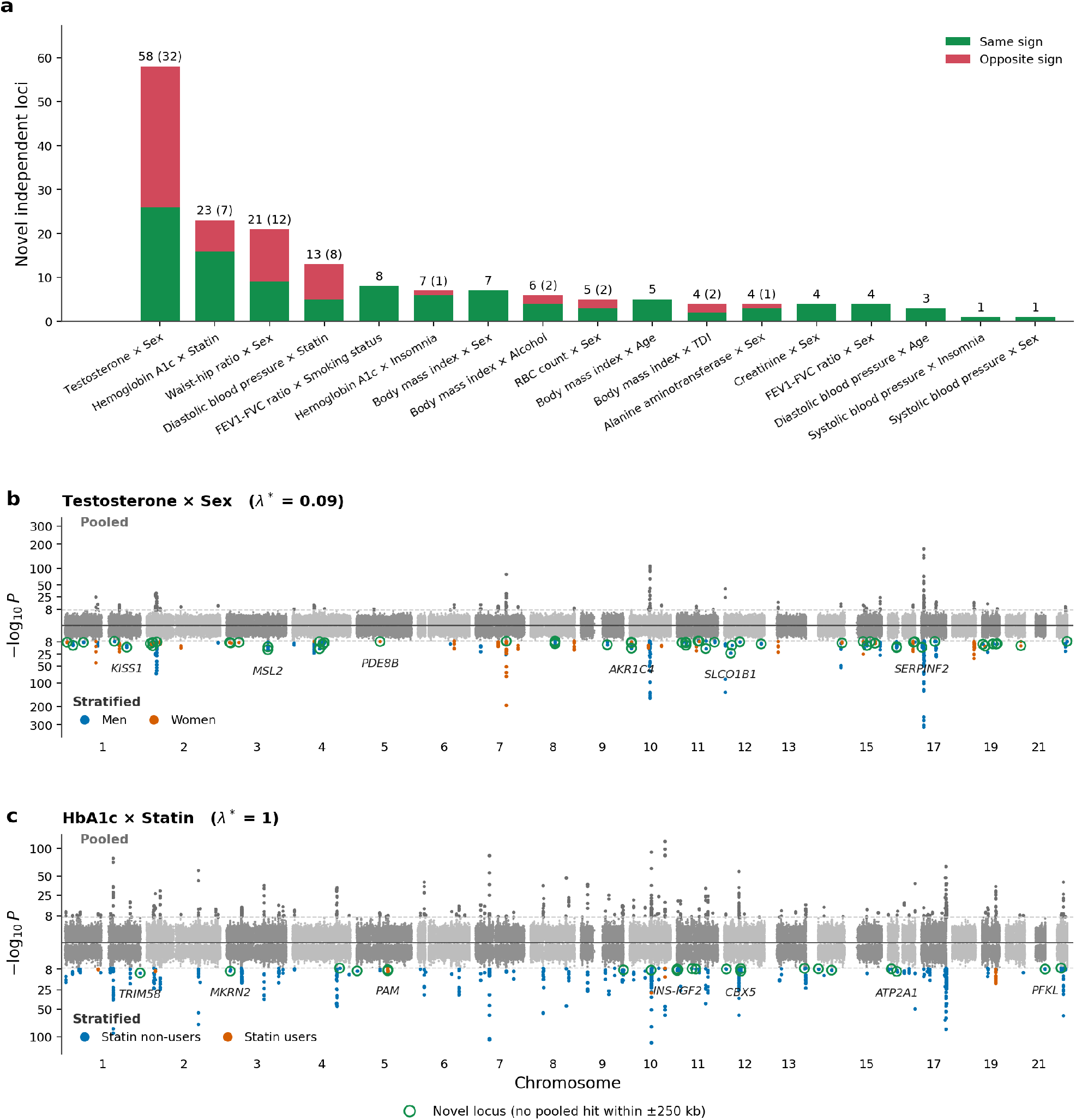
Loci recovered only by environment-stratified GWAS. **a**, Number of novel independent loci for each of the 20 trait-environment pairs with scale-independent G×E. Loci were considered novel if they reached genome-wide significance (P < 5 × 10^−8^, two-sided) in one environment while the pooled scan showed no genome-wide-significant association within ±250 kb. Stacked bars partition novel loci by whether the effects in the two environments share the same sign or have opposite signs; numbers above bars give the total, with opposite-sign loci in parentheses. Manhattan plots for **b**, testosterone stratified by sex, and **c**, HbA1c stratified by statin usage, analyzed at the optimal scale. Above the axis, we showed the results for the pooled scan over all individuals; below the axis, we showed the environment-stratified scan, in which each variant is plotted with the smaller of its two within-environment *p*-values. Genome-wide significant variants (*p* < 5 × 10^−8^) are colored by the environment for the stratified GWAS. Open green circles mark the novel loci, annotated within the nearest gene for the strongest non-overlapping ones.

The strongest effect was observed for testosterone stratified by sex, for which stratification identified 58 novel loci (41 male-specific and 17 female-specific), increasing the number of independent loci from 101 in the pooled analysis to 159 in the union of the two sex-stratified scans. This is consistent with the largely distinct genetic architectures of testosterone between sexes^20^. Male-specific signals included *KISS1* (1:204161591; *β* = −0.010, *p* = 3.1 × 10^−15^ in men versus *β* = −0.001, *p* = 0.6 in women; pooled *β* = 0.006, *p* = 7.1 × 10^−8^; Data S2), a regulator of the hypothalamic-pituitary-gonadal axis^26^. Among female-specific signals, *PDE8B* (5:76483059; *β* = −0.008, p =1.2 × 10^−8^ in women versus *β* = 0.002, p = 0.10 in men; pooled *β* = 0.010, p = 3.2 × 10^−5^) regulates cAMP signaling in the adrenal cortex^27^ (Figure 10b). Similar patterns were observed for other trait-environment pairs. WHR stratified by sex yielded 21 novel, predominantly female-driven loci, including the imprinted adipose regulator *KLF14*, which has sex-specific effects on fat distribution and type 2 diabetes risk^28^ (Figure S22). HbA1c stratified by statin use yielded 23 novel loci spanning both major biological determinants of the trait^29^. Glycemia-related signals included *INS-IGF2* ^30^ (p = 6.8 × 10^−9^ in non-users, with an opposite-sign effect in users) and the *β*-cell secretion gene *PAM* (*p* = 5.3 × 10^−9^ in users)^31^. Erythrocyte-related signals included the late-erythropoiesis regulator *TRIM58* ^32^ (*p* = 3.6 × 10^−11^) and the glycolytic enzyme *PFKL*^***33***^ (*p* = 9.9 × 10^−9^; Figure 10c).

## Discussion

We introduced REGENA, a unified variance-component framework that separates gene-by-environment interaction from interaction induced by the choice of phenotype scale *and* resolves GxE architecture by estimating the cross-environment genetic correlation and the environment-specific genetic variances across a set of phenotype transformations. Its randomized MoM estimator allows a dense grid of phenotype transformations to be scanned at little additional cost, making systematic analysis of the effects of scale tractable at biobank sizes.

Applying REGENA to quantitative trait-environment pairs in the UK Biobank, we find that a substantial number of interactions are scale-dependent and can be re-expressed as additive genetic and environmental effects under a suitable transformation. Scale-dependence does not by itself render an interaction to be artifactual, since the measured scale might be biologically or clinically relevant. Rather, distinguishing scale-dependent from scale-invariant interaction is a prerequisite for interpretation, and REGENA additionally localizes the parameter—genetic correlation or genetic variance—through which scale dependence operates. Among the interactions that persist across scales, most trait-environment pairs lie on a continuum between effect heterogeneity and amplification, and the dominant mechanism can differ across exposures even within a single trait.

The GxE architecture inferred by REGENA also refines the picture of context-specific genetic effects. Modeling GxE interactions recovers heritability that an additive model leaves unexplained: across scale-independent trait-environment pairs, the GxE heritability attributable to common SNPs increased the total heritability by 15% on average. Partitioning additive and GxE heritability by allele frequency, we find that polygenic GxE effects tend to be more specific at lower MAF so that interaction accounts for a larger share of genetic variance at low-frequency relative to common SNPs. This observation predicts that the relative contribution of GxE to trait variance is likely to increase at lower frequencies, a prediction that can be tested from whole-exome and whole-genome sequencing data^34,35^ and is expected under a model of pleiotropic stabilizing selection ^36^.

These architectural distinctions have consequences for both genetic prediction and gene discovery. Constructing GxE PGS from REGENA improves accuracy over an additive PGS, with the gains concentrated in trait-environment pairs whose genetic correlations differ across environments. Thus insights into architecture can indicate when stratified scoring is worthwhile, offering a principled criterion for departing from a pooled additive PGS. Stratifying by environments that show substantial polygenic GxE can also help define strata that are more homogeneous in genetic architecture than the pooled sample, which could, in turn, increase power to detect loci whose effects are heterogeneous across contexts. Our discovery of hundreds of novel genetic loci across the scale-independent trait-environment pairs, with one-third of the loci showing opposite signs across strata, supports the utility of this approach.

Our study has limitations that motivate future work. First, the use of the Box–Cox family – motivated by its simplicity, interpretability, and theoretical considerations - represents a limited set of transformations that may not correct for more complex types of scale dependence. REGENA is, however, flexible in the types of scale dependence that can be explored, although more sophisticated approaches to explore the space of transformations beyond a grid search might be needed. Going forward, the computational efficiency of REGENA allows sensitivity analyses, quantitatively using our framework for testing scale dependence but also more qualitatively (as in Figures 4a-d and Figure 5), to now be performed routinely which should serve as a useful starting point in assessing the robustness of GxE signals. Even more broadly, the basic framework underlying REGENA could be employed to test for scale-dependence of other aspects of genetic architecture. Second, our primary analyses encode environments as binary. This discretization preserves a direct interpretation of each variance component as a stratum-specific quantity, which is not available under continuous encoding. REGENA nevertheless supports finer categorical encodings, as illustrated for diastolic blood pressure across age quintiles (Figure S23); systematic application of such encodings remains an avenue for future work. Third, REGENA is currently restricted to quantitative traits. Extending the MoM formalism to binary traits, including disease status, while accounting for the role of ascertainment and covariates^37^ and being applicable at biobank scale is an important direction for future work. Fourth, we have not directly assessed the role of GxE in rare variants. This requires the REGENA framework to be extended directly to sequencing datasets and to handle the challenges arising from rare variant analyses. Finally, integrating REGENA’s environment-specific weights with summary-statistic methods^38^ could further improve the accuracy and cross-context portability of polygenic scores.

## Methods

### Environment-specific linear mixed model

We consider a binary environmental exposure vector ***E*** ∈ {0,1}^*N*^ across *N* individuals. Let ***x***_***j***_ ∈ ℝ^*NxM*^ denote the standardized genotypes of individual *i* across *M* variants, and let ***X*** ∈ ℝ^*N x M*^ be the corresponding genotype matrix (rows index individuals, columns index variants). We assume the phenotype ***y*** ∈ ℝ^*N*^ follows an environment-specific linear mixed model:

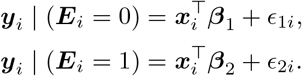

This model can be written in matrix form as

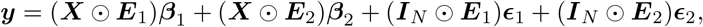

where ***E***_**2**_ = ***E, E***_**1**_ = **1** − ***E, I***_*N*_ is the *N × N* identity matrix, and ⊙ denotes element-wise multiplication with broadcasting over rows. We assume

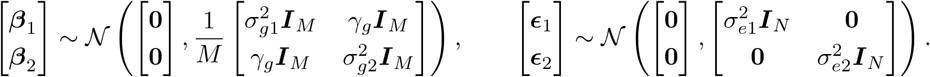

where 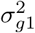 and 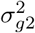 denote the environment-specific genetic variances, *γ*_*g*_ denotes the cross-environment genetic covariance, and 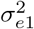 and 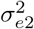 are the environment-specific residual variances. The total phenotypic covariance is

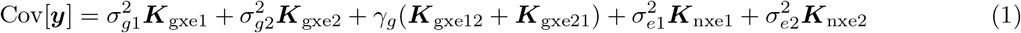

where 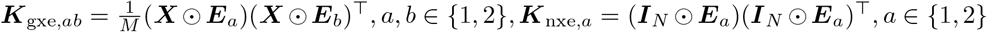 (see Notes S1). The cross-environment genetic correlation is

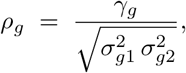

and environment-specific SNP-heritability estimates are

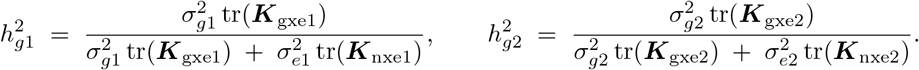

### Estimation of variance components

To estimate variance components, we use a method-of-moments (MoM) estimator^39,40,41^, which yields the normal equations

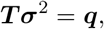

where 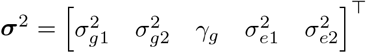. Each entry in ***T*** is

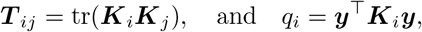

where {***K***_***i***_} are the kernels appearing in Equation 1, with the cross-covariance parameter *y*_*g*_ associated with the symmetric kernel (**K**_**gxe12**_ + **K**_**gxe21**_). Under the observation that the environment vectors are binary indicators (see Notes S2), the normal equations decouple into two independent linear systems for the environment-specific components:

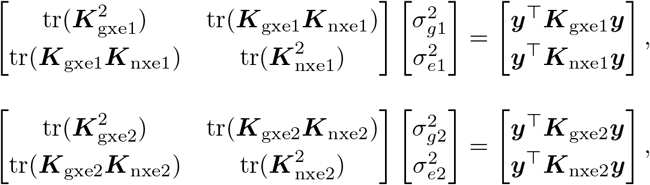

and, for the cross-environment covariance,

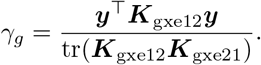

### Randomized method-of-moments estimation

The dominant cost in solving **T*σ***^***2***^ = ***q*** lies in assembling the trace matrix **T**, whose entries **T**_**i**_**j** = tr [**K**_**i**_**Kj**] each require forming the GRMs and multiplying two *N* x *N* matrices at a cost of *O(N*^***3***^), the principal computational bottleneck. To avoid this, we estimate each trace with the randomized trace estimator^42^. Let ***z***_*k*_ be an N-vector with zero mean and identity covariance; then tr 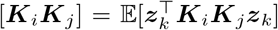, so that 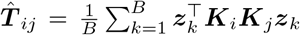. Writing ***X***_*a*_ = ***X ⊙ E***_*a*_, each genetic GRM factors as 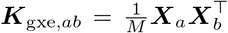 with *a,b* ∈ {1, 2}, so the product 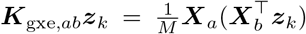 evaluates as two matrix-vector products and we never form or store any GRM explicitly. In practice, we draw the *B* probe vectors **z**_**k**_ from a standard normal distribution, which lowers the cost of computing **T**_***i***_***j*** from O(N^**3**^) to *O(NMB*). We further exploit the structure of the genotype matrix, whose entries take only the values 0, 1, or 2, via the Mailman algorithm^43^, reducing the cost of each matrix-vector product from *O(NM*) to *O(NM/* max(log_3_ N, log_3_ M)). The resulting runtime for assembling the normal equations is 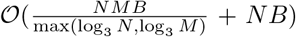 a substantial improvement over the *O*(*N*^3^) cost of direct computation when *B* is small. We set *B* = 10 by default, which we have shown in prior work balances statistical and computational effciency^9^.

### Efficient estimation of variance components over phenotype transformations

Inferences about genetic architecture can depend on transformations applied to the analyzed phenotype, especially in the presence of scale dependence, floor or ceiling effects, or nonlinear exposure-response relationships. To understand the effect of scale transformations, we pick a set of *L* transformations 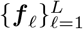 and attempt to estimate the variance components corresponding to each of the transformed phenotypes ***f***_***ℓ***_ (***y***). For example, for a parametric family of transformation functions ***f***_***θ***_, we choose parameters across a grid of size *L* and estimate the variance components across this grid. A naive approach to inference involves estimating the variance components separately for each transformation on this grid. However, such an approach is computationally intensive particularly for biobank-scale data.

The challenge that we aim to solve is to enable estimation of variance components across a set of transformations in an efficient manner. We compute **q**(**f**) for each transformation and stack them as ***Q*** = [***q***(***f*** 1),…, ***q***(***f***_***L***_)] ∈ ℝ^*PxL*^, where *P* is the number of variance components. We estimate variance components using the MoM framework, which solves the system 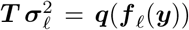, where 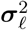 collects the variance components, ***T***_*i j*_ = tr(**K**_*i*_***K***_*j*_) depends only on the genotype and environment kernels {***K***_***i***_}, and ***q***(***f***_***ℓ***_ (***y***)) collects quadratic forms of the transformed phenotype. Importantly, ***T*** is independent of the phenotype and can be precomputed once. All variance components *Σ*_(1:*L*)_ are then obtained in parallel via ***Σ*** ^(1:***L***^) = ***T***^−1^***Q***. Here, Σ _(1:*L*)_ ∈ ℝ ^*PxL*^, with its *i*-th column giving the stacked variance-component estimates for the *i*-th transformation.

Because **T** is fixed and estimated efficiently through randomized MoM, the cost of computing variance components per-transformation is small making it feasible to scan over hundreds or thousands of transformations thereby enabling systematic exploration of phenotype scaling at biobank-scale.

### Box-Cox transformation

While REGENA can explore the impact of a variety of transformations, in this work, we consider transformations from the one-parameter Box-Cox family^18^, which includes the identity, logarithmic, and reciprocal

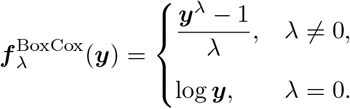

This family is continuous in *λ* and captures many commonly used scale adjustments. For each trait-environment pair, we evaluate the transformation on a grid of *λ* values, after applying a constant shift to ensure *y*_*i*_ > 0 when necessary.

Extreme values of *λ* can induce heavy tails in the transformed phenotype, creating outliers and destabilizing downstream inference. To mitigate this, we restrict attention to values of *λ* under which the transformed data are not inconsistent with a Gaussian distribution (see Notes S4).

### Scale-independent GxE across transformations

We summarize the GxE mechanisms in terms of the parameters of Equation 1. Following Durvasula and Price ^16^, we consider three mechanisms:

1. **Imperfect genetic correlation** (*ρ*_*g*_ *<* 1). Genetic effects differ between environments, either because different loci contribute or because effect sizes differ at shared loci.
2. **Varying genetic variance** 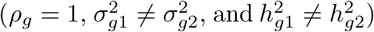. Genetic effects are perfectly correlated across environments (same loci and proportional effect sizes), but the marginal genetic variances and heritabilities differ.
3. **Proportional amplification** 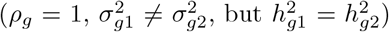. Genetic variances differ, but this difference is exactly offset by a difference in total phenotypic variance, so that heritability remains constant.

The latter two mechanisms together constitute *amplification* (*ρ*_*g*_ = 1 with 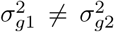, distinguished by whether heritability is preserved.

To study scale-robust GxE effects, we apply the Box-Cox transformation over a grid of *L* values 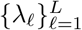 and jointly estimate the variance components for all transformed phenotypes using the batched method-of-moments procedure above. The outputs across the L transformations are 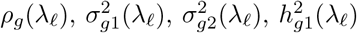 and 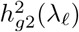. At each A we compute Wald-type statistics

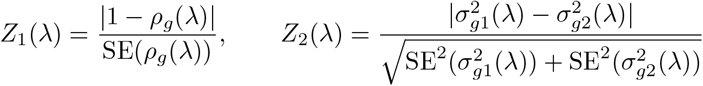

associated with two-sided Wald p-values 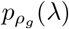 and 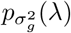.

We define a trait-environment pair as scale-dependent if there exists at least one Box-Cox parameter *λ* such that (i) the genetic correlation is not significantly different from 1 and (ii) the genetic variances do not differ significantly between environments. Formally, for each transformation, we test the composite null 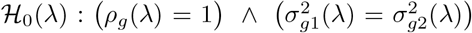. For scale-independent pairs, we further identify the GxE mechanisms on the reporting scale. We therefore outlined the scale-based GxE analysis pipeline below (Algorithm 1).

### Multiple-testing control for scale-independent GxE

We first focus on the test for a single transformation. Given *λ*, let *S*_***1***_(*λ*) and *S*_***2***_(*λ*) denote the rejection regions for *Z*_***1***_(*λ*) and *Z*_***2***_(*λ*), and define the rejection event *R(λ)* = 1{*Z*_***1***_(*λ*) ∈ *S*_***1***_(*λ*) V *Z*_***2***_(*λ*) ∈ *S*_***2***_(*λ*)}. By the union bound, under *ℋ*_**0**_(*λ*), Pr(*R*(*λ*)) ≤ Pr(*Z*_**1**_ (*λ*) *∈ S*_1_(*λ*)) + Pr(*Z*_**2**_(*λ*) *∈ S*_**2**_(*λ*)), so choosing per-test thresholds α/2 controls Pr(R(*λ*)) ≤ *α*.

We now combine the tests across transformations. Consistent with the definition above, a pair is scaledependent if ℋ_0_ (*λ*) holds for some *λ*, so the null is

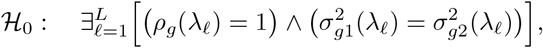

which we reject only if every per-*λ* test is rejected. Let p(*λ*_*ℓ*_) be the per-*λ p*-value and let *L*_**o**_ − {*ℓ* : *ℋ*_**0**_(*λ*_ℓ_) holds}. At threshold *t*,

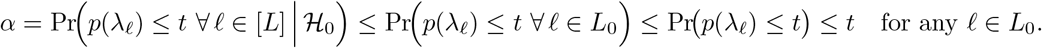

Therefore, rejecting when *p*(λ_ℓ_) ≤ *t* for all ℓ ensures the combined test has type I error at most *t*.

Accounting for multiple testing across *n*_*t*_ traits and *n*_*e*_ environments, we test each component of the composite null at level α/(2*n*_***t***_*n*_***e***_). Single-parameter tests reported in subsequent analyses (for example, per-bin tests of *ρ*_*g*_(λ) =1 or of 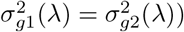 are Bonferroni-corrected across the *n*_*t*_*n*_*e*_ trait-environment pairs at level α/(*n*_***t***_*n*_***e***_), and tissue-stratified analyses are corrected across the number of tissue annotations tested.

#### Algorithm 1

**Scale-based classification of gene—environment interaction**.

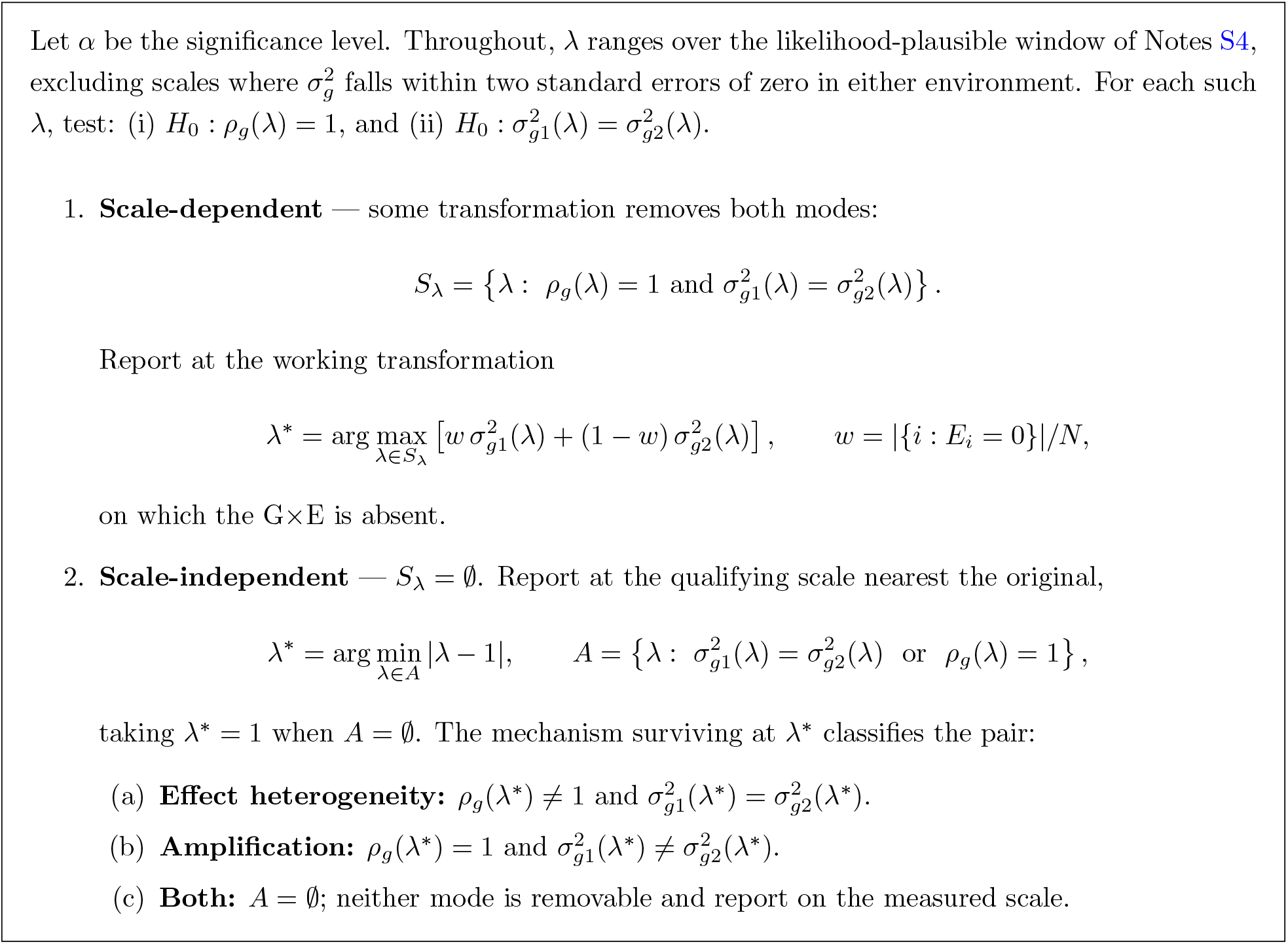

### Partitioned G ×E and polygenic prediction

#### Functional partitioning

We partition the genotype matrix into *K* non-overlapping annotations 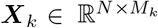 and assign each annotation independent, environment-correlated effects with annotation-specific variances 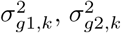 and cross-environment covariance *ϒ*_*g*_,_*k*_ (Notes S10). The phenotypic covariance is then a sum of annotation-specific G ×E kernels,

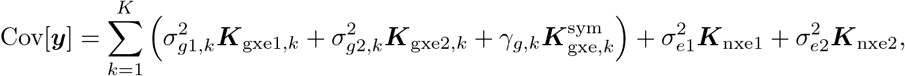

with per-annotation genetic correlation 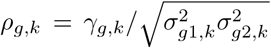. The MoM system expands analogously with annotation-specific kernels, retaining the block-diagonal structure induced by the disjoint environments, which lets us partition G ×E heritability across annotations and test enrichment in trait-relevant functional categories.

#### Categorical environments

For a categorical environment with *K* mutually exclusive categories (one-hot indicators ***E***_1_,…, ***E***_*K*_ with Σ_***k***_ ***E***_*k*_ = **1**), the two-environment model generalizes to a *K*-category effect prior with genetic covariance **Σ**_***g***_(diagonal 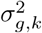 off-diagonal *ϒ*_*g,kl*_; Notes S11), yielding

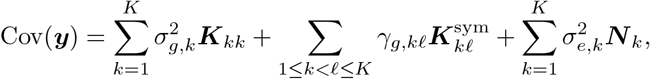

with pairwise genetic correlations 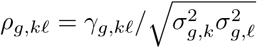.

#### Environment-specific polygenic scores

Given the estimated variance components, we derive polygenic scores whose SNP weights are the posterior-mean (BLUP) effects^44^ (Notes S12). These polygenic scores adapt to the architecture of G ×E underlying the trait in a data-driven manner. When the estimated cross-environment genetic covariance *ϒ*_*g*_ is close to zero, indicating that SNP effects in one environmental stratum are uncorrelated with those in the other, the polygenic scores are estimated separately for each environmental stratum. On the other hand, when *ϒ*_*g*_ is large in magnitude, SNP effects are shared across the two strata. Because forming these weights requires inverting the phenotypic covariance, which is infeasible to materialize at biobank scale, we obtain them by iteratively solving the corresponding linear system with preconditioned conjugate gradients^45,46^, accelerated by a preconditioner that rescales residuals separately within each environment stratum. Predictive accuracy is assessed out-of-sample through the gain in variance explained when the score is added to a covariate-only baseline, Δ*R*^2^ = *R*^2^(covariates+PGS) — *R*^***2***^ (covariates), reported both across all individuals and within strata, with the environment indicator admitted as an optional covariate; under the stratified scheme each individual receives the score matched to their own environment. Standard errors and percentile confidence intervals for the two *R*^***2***^ terms and their difference are obtained by resampling individuals in the held-out set with replacement, holding the trained weights fixed so that the quantified uncertainty pertains to evaluation alone and not to refitting the genetic model. To assess discrimination in the tails, we form a covariate-adjusted phenotype *z*_*i*_ by regressing the phenotype on the covariates and the environment indicator and scaling the residual to standard-deviation (SD) units; each score is evaluated against the phenotype on which it was trained. Ranking the *n* held-out individuals by the score s, we take the ⌈*pn* ⌉highest and lowest as *T*_*p*_ and *B*_*p*_ (for example, *p* = 1%) and record 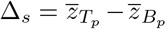. We compare the two scores through Δ_G× E_ — Δ_baseline_, computed separately for each trait-environment pair.

### UK Biobank Data

We applied REGENA to the UK Biobank^47^ across 43 quantitative phenotypes and seven binary environmental exposures: sex, age, statin use, smoking status, alcohol consumption, insomnia, and the Townsend deprivation index. For each trait-environment pair, we fit a model that included the environmental variable as a main effect and allowed environment-specific residual variance. The analyses used the UK Biobank Resource under application 33127, with participants’ informed consent verified by the UK Biobank^48^.

#### Genotype quality control

Our primary analyses were restricted to SNPs present on the UK Biobank Axiom genotyping array. We removed SNPs with greater than 1% missingness or minor allele frequency below 1%, and SNPs failing a Hardy-Weinberg equilibrium test at *p <* 10^−7^. We restricted the sample to self-reported British white ancestry individuals who were greater than third-degree relatives, defined as pairs with kinship coefficient below 1/2^9/247^, and we further removed individuals flagged as outliers for genotype heterozygosity or missingness. This yielded *N* = 291,273 individuals and *M* = 454,207 SNPs.

In an additional analysis, we included high-quality imputed variants with MAF ≥ 0.1% in *N* = 291, 273 unrelated white British individuals. Variants were required to have an INFO score ≥ 0.8, a hard-call threshold of 0.2 and genotype calls available in more than 99% of individuals. We further excluded variants deviating from Hardy-Weinberg equilibrium (p < 10^−7^) and those located within the major histocompatibility complex region, yielding a final set of M = 7, 774, 235 variants.

#### Covariates

For all traits, we included age, sex, age^2^, age×sex, age^2^×sex, and the top 20 genetic principal components (PCs) as covariates, and we always additionally included the focal environmental variable as a covariate. We used PCs precomputed by the UK Biobank from a superset of 488,295 individuals. Each phenotype was retained on its measured (raw) scale prior to the Box-Cox scan.

#### Environmental exposure definitions

Statin use was defined as a binary variable based on the Anatomical Therapeutic Chemical code C10AA, corresponding to any statin subtype. Smoking status was encoded as never, previous, or current use, and dichotomized into ever versus never use for the binary analyses. The remaining continuous exposures (age, alcohol consumption, insomnia, and the Townsend deprivation index) were dichotomized into lower and higher strata. For the categorical-environment analysis, age was instead discretized into quintiles.

#### Annotations

For heritability partitioned by MAF and LD, we divided variants into eight annotations formed by crossing four quartiles of in-sample LD score (computed with GCTA^49^) with two MAF bins (MAF < 5% and MAF ≥ 5%).

## Data Availability

This study used individual-level genotype and phenotype data from the UK Biobank, accessed under Application 33127. These data are available to researchers upon application to UK Biobank (https://www.ukbiobank.ac.uk/enable-your-research/apply-for-access). Software implementing REGENA is available at https://github.com/sriramlab/REGENA.

## Software availability

https://github.com/sriramlab/REGENA

## Acknowledgments

This research was conducted using data from the UK Biobank Resource under application number 33127. We gratefully acknowledge the UK Biobank participants for their contributions to this research. This work was supported in part by NIH grant R35GM153406 (Z.L., A.A., M.J., and S.S.) and NSF CAREER award 1943497 (S.S.).

We thank Francesco Paolo Casale for valuable feedback on this project. We would also like to acknowledge stimulating discussions at the UCLA Computational Genomics Summer Institute, funded by NIH grants GM135043 and GM112625.

## References

[1] David J Hunter. Gene–environment interactions in human diseases. Nature reviews genetics, 6(4): 287–298, 2005.

[2] Esther Herrera-Luis, Kelly Benke, Heather Volk, Christine Ladd-Acosta, and Genevieve L Wojcik. Gene–environment interactions in human health. Nature Reviews Genetics, 25(11):768–784, 2024.

[3] Shinichi Namba, Kyuto Sonehara, Yuriko N Koyanagi, Takezo Kikuchi, Takafumi Ojima, Ryuya Edahiro, Go Sato, Taiki Yamaji, Yoshihiko Tomofuji, Hiroyuki Ueda, et al. A cross-population compendium of gene–environment interactions. Nature, 651(8106):688–697, 2026.

[4] Rachel Moore, Francesco Paolo Casale, Marc Jan Bonder, Danilo Horta, Lude Franke, Inês Barroso, and Oliver Stegle. A linear mixed-model approach to study multivariate gene–environment interactions. Nature genetics, 51(1):180–186, 2019.

[5] Jessica Tyrrell, Andrew R Wood, Ryan M Ames, Hanieh Yaghootkar, Robin N Beaumont, Samuel E Jones, Marcus A Tuke, Katherine S Ruth, Rachel M Freathy, George Davey Smith, et al. Gene– obesogenic environment interactions in the uk biobank study. International journal of epidemiology, 46 (2):559–575, 2017.

[6] Huanwei Wang, Futao Zhang, Jian Zeng, Yang Wu, Kathryn E Kemper, Angli Xue, Min Zhang, Joseph E Powell, Michael E Goddard, Naomi R Wray, et al. Genotype-by-environment interactions inferred from genetic effects on phenotypic variability in the uk biobank. Science advances, 5(8):eaaw3538, 2019.

[7] Jiacheng Miao, Gefei Song, Yixuan Wu, Jiaxin Hu, Yuchang Wu, Shubhashrita Basu, James S Andrews, Katherine Schaumberg, Jason M Fletcher, Lauren L Schmitz, et al. Pigeon: a statistical framework for estimating gene–environment interaction for polygenic traits. Nature human behaviour, 9(8):1654–1668, 2025.

[8] Matthew Kerin and Jonathan Marchini. Inferring gene-by-environment interactions with a bayesian whole-genome regression model. The American Journal of Human Genetics, 107(4):698–713, 2020.

[9] Ali Pazokitoroudi, Zhengtong Liu, Andrew Dahl, Noah Zaitlen, Saharon Rosset, and Sriram Sankararaman. A scalable and robust variance components method reveals insights into the architecture of gene-environment interactions underlying complex traits. The American Journal of Human Genetics, 111(7):1462–1480, 2024. doi: 10.1016/j.ajhg.2024.05.015.

[10] Matthew R Robinson, Geoffrey English, Gerhard Moser, Luke R Lloyd-Jones, Marcus A Triplett, Zhihong Zhu, Ilja M Nolte, Jana V van Vliet-Ostaptchouk, Harold Snieder, LifeLines Cohort Study, et al. Genotype–covariate interaction effects and the heritability of adult body mass index. Nature genetics, 49(8):1174–1181, 2017.

[11] Manuela Costantino, Renée Fonseca, Zhengtong Liu, Zhenhong Huang, Sriram Sankararaman, Iain Mathieson, and Andy Dahl. Choice of phenotype scale is critical in biobank-based G×E tests. bioRxiv, 2026.

[12] Michal Sadowski, Andy W. Dahl, Noah Zaitlen, and Richard Border. The geometry of G×E: how scaling and endogenous treatment effects shape interaction direction. PLOS Genetics, 2026. doi: 10.1371/journal.pgen.1012073. bioRxiv preprint 2025; published in PLOS Genetics 2026.

[13] Nicolo Fusi, Christoph Lippert, Neil D Lawrence, and Oliver Stegle. Warped linear mixed models for the genetic analysis of transformed phenotypes. Nature communications, 5(1):4890, 2014.

[14] Zhenhong Huang, Manuela Costantino, and Andy Dahl. Optimizing phenotype scale improves genetic analyses in large-scale biobanks. bioRxiv, 2026.

[15] Carrie Zhu, Matthew J. Ming, Jared M. Cole, Michael D. Edge, Mark Kirkpatrick, and Arbel Harpak. Amplification is the primary mode of gene-by-sex interaction in complex human traits. Cell Genomics, 3(5):100297, 2023. doi: 10.1016/j.xgen.2023.100297.

[16] Arun Durvasula and Alkes L. Price. Distinct explanations underlie gene-environment interactions in the UK Biobank. The American Journal of Human Genetics, 112(3):644–658, 2025. doi: 10.1016/j.ajhg.2025.01.014.

[17] Kenneth E Westerman and Tamar Sofer. Many roads to a gene-environment interaction. The American Journal of Human Genetics, 111(4):626–635, 2024.

[18] George EP Box and David R Cox. An analysis of transformations. Journal of the Royal Statistical Society Series B: Statistical Methodology, 26(2):211–243, 1964.

[19] Nasa Sinnott-Armstrong, Sahin Naqvi, Manuel Rivas, and Jonathan K. Pritchard. GWAS of three molecular traits highlights core genes and pathways alongside a highly polygenic background. eLife, 10: e58615, 2021. doi: 10.7554/eLife.58615.

[20] Katherine S Ruth, Felix R Day, Jessica Tyrrell, Deborah J Thompson, Andrew R Wood, Anubha Mahajan, Robin N Beaumont, Laura Wittemans, Susan Martin, Alexander S Busch, et al. Using human genetics to understand the disease impacts of testosterone in men and women. Nature medicine, 26(2):252–258, 2020.

[21] Junxi Liu, Rebecca C Richmond, Jack Bowden, Ciarrah Barry, Hassan S Dashti, Iyas Daghlas, Jacqueline M Lane, Samuel E Jones, Andrew R Wood, Timothy M Frayling, et al. Assessing the causal role of sleep traits on glycated hemoglobin: a mendelian randomization study. Diabetes Care, 45(4):772–781, 2022.

[22] Heming Wang, Pavithra Nagarajan, Clint L Miller, Amy R Bentley, Raymond Noordam, Kenneth E Westerman, Michael R Brown, Aldi T Kraja, Jeffrey R O’Connell, Karen Schwander, et al. Gene-by-sleep duration interaction for glycemic traits in over 480,000 individuals. medRxiv, 2026.

[23] Jeffrey P Spence, Hakhamanesh Mostafavi, Mineto Ota, Nikhil Milind, Tamara Gjorgjieva, Courtney J Smith, Yuval B Simons, Guy Sella, and Jonathan K Pritchard. Specificity, length and luck drive gene rankings in association studies. Nature, 649(8098):918–925, 2026.

[24] Minhui Chen, Xinpei Wang, Lena Krockenberger, Rika Tyebally, Jeremy J Berg, Sebastian Pott, Jonathan Flint, Joseph E Powell, Brunilda Balliu, Xuanyao Liu, and Andy Dahl. Cell-type-specific eqtls underlie the genetic architecture of complex traits. Nature, 2026.

[25] Irene Novo, Eugenio López-Cortegano, and Armando Caballero. Highly pleiotropic variants of human traits are enriched in genomic regions with strong background selection. Human Genetics, 140(9): 1343–1351, 2021.

[26] A Kemal Topaloglu, Javier A Tello, L Damla Kotan, Mehmet N Ozbek, M Bertan Yilmaz, Seref Erdogan, Fatih Gurbuz, Fatih Temiz, Robert P Millar, and Bilgin Yuksel. Inactivating kiss1 mutation and hypogonadotropic hypogonadism. New England Journal of Medicine, 366(7):629–635, 2012.

[27] Anelia Horvath, Christoforos Giatzakis, Kitman Tsang, Elizabeth Greene, Paulo Osorio, Sosipatros Boikos, Rossella Libé, Yianna Patronas, Audrey Robinson-White, Elaine Remmers, et al. A camp-specific phosphodiesterase (pde8b) that is mutated in adrenal hyperplasia is expressed widely in human and mouse tissues: a novel pde8b isoform in human adrenal cortex. European Journal of Human Genetics, 16(10):1245–1253, 2008.

[28] Kerrin S Small, Marijana Todorčević, Mete Civelek, Julia S El-Sayed Moustafa, Xiao Wang, Michelle M Simon, Juan Fernandez-Tajes, Anubha Mahajan, Momoko Horikoshi, Alison Hugill, et al. Regulatory variants at klf14 influence type 2 diabetes risk via a female-specific effect on adipocyte size and body composition. Nature genetics, 50(4):572–580, 2018.

[29] Eleanor Wheeler, Aaron Leong, Ching-Ti Liu, Marie-France Hivert, Rona J. Strawbridge, Clara Podmore, Man Li, Jie Yao, Xueling Sim, Jaeyoung Hong, et al. Impact of common genetic determinants of Hemoglobin A1c on type 2 diabetes risk and diagnosis in ancestrally diverse populations: A transethnic genome-wide meta-analysis. PLoS Medicine, 14(9):e1002383, 2017. doi: 10.1371/journal.pmed.1002383.

[30] Anubha Mahajan, Daniel Taliun, Matthias Thurner, Neil R Robertson, Jason M Torres, N William Rayner, Anthony J Payne, Valgerdur Steinthorsdottir, Robert A Scott, Niels Grarup, et al. Fine-mapping type 2 diabetes loci to single-variant resolution using high-density imputation and islet-specific epigenome maps. Nature genetics, 50(11):1505–1513, 2018.

[31] Soren K Thomsen, Anne Raimondo, Benoit Hastoy, Shahana Sengupta, Xiao-Qing Dai, Austin Bautista, Jenny Censin, Anthony J Payne, Mahesh M Umapathysivam, Aliya F Spigelman, et al. Type 2 diabetes risk alleles in pam impact insulin release from human pancreatic β-cells. Nature genetics, 50(8):1122–1131, 2018.

[32] Christopher S Thom, Elizabeth A Traxler, Eugene Khandros, Jenna M Nickas, Olivia Y Zhou, Jacob E Lazarus, Ana PG Silva, Dolly Prabhu, Yu Yao, Chiaka Aribeana, et al. Trim58 degrades dynein and regulates terminal erythropoiesis. Developmental cell, 30(6):688–700, 2014.

[33] Richard van Wijk and Wouter W van Solinge. The energy-less red blood cell is lost: erythrocyte enzyme abnormalities of glycolysis. Blood, 106(13):4034–4042, 2005.

[34] Pierrick Wainschtein, Deepti Jain, Zhili Zheng, L Adrienne Cupples, Aladdin H Shadyab, Barbara McKnight, Benjamin M Shoemaker, Braxton D Mitchell, et al. Assessing the contribution of rare variants to complex trait heritability from whole-genome sequence data. Nature genetics, 54(3):263–273, 2022.

[35] Pierrick Wainschtein, Yuanxiang Zhang, Jeremy Schwartzentruber, Irfahan Kassam, Julia Sidorenko, Petko P Fiziev, Huanwei Wang, Jeremy McRae, Richard Border, Noah Zaitlen, et al. Estimation and mapping of the missing heritability of human phenotypes. Nature, 649(8099):1219–1227, 2026.

[36] Yuval B Simons, Kevin Bullaughey, Richard R Hudson, and Guy Sella. A population genetic interpretation of gwas findings for human quantitative traits. PLoS biology, 16(3):e2002985, 2018.

[37] David Golan, Eric S Lander, and Saharon Rosset. Measuring missing heritability: inferring the contribution of common variants. Proceedings of the National Academy of Sciences, 111(49):E5272–E5281, 2014.

[38] Florian Privé, Julyan Arbel, and Bjarni J. Vilhjálmsson. LDpred2: better, faster, stronger. Bioinformatics, 36(22-23):5424–5431, 2020. doi: 10.1093/bioinformatics/btaa1029.

[39] Joseph K Haseman and Robert C Elston. The investigation of linkage between a quantitative trait and a marker locus. Behavior genetics, 2(1):3–19, 1972.

[40] Yue Wu and Sriram Sankararaman. A scalable estimator of snp heritability for biobank-scale data. Bioinformatics, 34(13):i187–i194, 2018.

[41] Ali Pazokitoroudi, Yue Wu, Kathryn S Burch, Kangcheng Hou, Aaron Zhou, Bogdan Pasaniuc, and Sriram Sankararaman. E”cient variance components analysis across millions of genomes. Nature communications, 11(1):4020, 2020.

[42] M. F. Hutchinson. A stochastic estimator of the trace of the influence matrix for laplacian smoothing splines. Communications in Statistics - Simulation and Computation, 18(3):1059–1076, 1989. doi: 10.1080/03610918908812806.

[43] Edo Liberty and Steven W Zucker. The mailman algorithm: A note on matrix–vector multiplication. Information Processing Letters, 109(3):179–182, 2009.

[44] Charles R Henderson. Best linear unbiased estimation and prediction under a selection model. Biometrics, pages 423–447, 1975.

[45] Magnus R Hestenes and Eduard Stiefel. Methods of conjugate gradients for solving linear systems. Journal of research of the National Bureau of Standards, 49(6):409–436, 1952.

[46] Po-Ru Loh, George Tucker, Brendan K Bulik-Sullivan, Bjarni J Vilhjàlmsson, Hilary K Finucane, Rany M Salem, Daniel I Chasman, Paul M Ridker, Benjamin M Neale, Bonnie Berger, et al. E”cient bayesian mixed-model analysis increases association power in large cohorts. Nature genetics, 47(3): 284–290, 2015.

[47] Clare Bycroft, Colin Freeman, Desislava Petkova, Gavin Band, Lloyd T Elliott, Kevin Sharp, Allan Motyer, Damjan Vukcevic, Olivier Delaneau, Jared O’Connell, et al. The UK Biobank resource with deep phenotyping and genomic data. Nature, 562(7726):203–209, 2018.

[48] Cathie Sudlow, John Gallacher, Naomi Allen, Valerie Beral, Paul Burton, John Danesh, Paul Downey, Paul Elliott, Jane Green, Martin Landray, et al. UK Biobank: An Open Access Resource for Identifying the Causes of a Wide Range of Complex Diseases of Middle and Old Age. PLoS medicine, 12(3): e1001779, 2015.

[49] Jian Yang, S Hong Lee, Michael E Goddard, and Peter M Visscher. Gcta: a tool for genome-wide complex trait analysis. The American journal of human genetics, 88(1):76–82, 2011.

[50] Matteo Di Scipio, Mohammad Khan, Shihong Mao, Michael Chong, Conor Judge, Nazia Pathan, Nicolas Perrot, Walter Nelson, Ricky Lali, Shuang Di, et al. A versatile, fast and unbiased method for estimation of gene-by-environment interaction effects on biobank-scale datasets. Nature Communications, 14(1): 5196, 2023.

[51] Andy Dahl, Khiem Nguyen, Na Cai, Michael J. Gandal, Jonathan Flint, and Noah Zaitlen. A robust method uncovers significant context-specific heritability in diverse complex traits. The American Journal of Human Genetics, 106(1):71–91, 2020. doi: 10.1016/j.ajhg.2019.11.015.

