## Supplementary Materials for "Disentangling phenotype scale, amplification, and effect heterogeneity underlying polygenic GxE architecture of complex traits"

### Supplementary Information

**Supplementary Notes** **2**

**Supplementary Data** **48**

### Supplementary Notes

#### S1 Phenotypic covariance of environment-specific linear mixed model

We have the environment-specific linear mixed model as

$$\boldsymbol{y} = (\boldsymbol{X} \odot \boldsymbol{E}_1)\boldsymbol{\beta}_1 + (\boldsymbol{X} \odot \boldsymbol{E}_2)\boldsymbol{\beta}_2 + (\boldsymbol{I}_N \odot \boldsymbol{E}_1)\boldsymbol{\epsilon}_1 + (\boldsymbol{I}_N \odot \boldsymbol{E}_2)\boldsymbol{\epsilon}_2,$$

under the assumption that $(\boldsymbol{\beta}_1, \boldsymbol{\beta}_2) \perp\!\!\!\perp (\boldsymbol{\epsilon}_1, \boldsymbol{\epsilon}_2)$ and

$$\begin{bmatrix} \boldsymbol{\beta}_1 \\ \boldsymbol{\beta}_2 \end{bmatrix} \sim \mathcal{N}\left( \begin{bmatrix} \mathbf{0} \\ \mathbf{0} \end{bmatrix}, \frac{1}{M} \begin{bmatrix} \sigma_{g1}^2 \boldsymbol{I}_M & \gamma_g \boldsymbol{I}_M \\ \gamma_g \boldsymbol{I}_M & \sigma_{g2}^2 \boldsymbol{I}_M \end{bmatrix} \right), \qquad \begin{bmatrix} \boldsymbol{\epsilon}_1 \\ \boldsymbol{\epsilon}_2 \end{bmatrix} \sim \mathcal{N}\left( \begin{bmatrix} \mathbf{0} \\ \mathbf{0} \end{bmatrix}, \begin{bmatrix} \sigma_{e1}^2 \boldsymbol{I}_N & \mathbf{0} \\ \mathbf{0} & \sigma_{e2}^2 \boldsymbol{I}_N \end{bmatrix} \right).$$

Define

$$\boldsymbol{X}_1 = \boldsymbol{X} \odot \boldsymbol{E}_1, \quad \boldsymbol{X}_2 = \boldsymbol{X} \odot \boldsymbol{E}_2, \quad \boldsymbol{I}_1 = \boldsymbol{I}_N \odot \boldsymbol{E}_1, \quad \boldsymbol{I}_2 = \boldsymbol{I}_N \odot \boldsymbol{E}_2.$$

Because the covariances between $\boldsymbol{\beta}$ and $\boldsymbol{\epsilon}$ are zero, the phenotypic covariance is

$$\begin{aligned} \mathrm{Cov}[\boldsymbol{y}] &= \mathrm{Cov}[\boldsymbol{X}_1\boldsymbol{\beta}_1 + \boldsymbol{X}_2\boldsymbol{\beta}_2 + \boldsymbol{I}_1\boldsymbol{\epsilon}_1 + \boldsymbol{I}_2\boldsymbol{\epsilon}_2] \\ &= \mathrm{Cov}[\boldsymbol{X}_1\boldsymbol{\beta}_1] + \mathrm{Cov}[\boldsymbol{X}_2\boldsymbol{\beta}_2] + \mathrm{Cov}[\boldsymbol{X}_1\boldsymbol{\beta}_1, \boldsymbol{X}_2\boldsymbol{\beta}_2] + \mathrm{Cov}[\boldsymbol{X}_2\boldsymbol{\beta}_2, \boldsymbol{X}_1\boldsymbol{\beta}_1] \\ &\quad + \mathrm{Cov}[\boldsymbol{I}_1\boldsymbol{\epsilon}_1] + \mathrm{Cov}[\boldsymbol{I}_2\boldsymbol{\epsilon}_2]. \end{aligned}$$

We compute each term:

$$\begin{aligned} \mathrm{Cov}[\boldsymbol{X}_1\boldsymbol{\beta}_1] &= \boldsymbol{X}_1 \,\mathrm{Cov}[\boldsymbol{\beta}_1]\, \boldsymbol{X}_1^\top = \frac{\sigma_{g1}^2}{M}\boldsymbol{X}_1\boldsymbol{X}_1^\top, \\ \mathrm{Cov}[\boldsymbol{X}_2\boldsymbol{\beta}_2] &= \frac{\sigma_{g2}^2}{M}\boldsymbol{X}_2\boldsymbol{X}_2^\top, \\ \mathrm{Cov}[\boldsymbol{X}_1\boldsymbol{\beta}_1, \boldsymbol{X}_2\boldsymbol{\beta}_2] &= \boldsymbol{X}_1 \,\mathrm{Cov}[\boldsymbol{\beta}_1, \boldsymbol{\beta}_2]\, \boldsymbol{X}_2^\top = \frac{\gamma_g}{M}\boldsymbol{X}_1\boldsymbol{X}_2^\top, \\ \mathrm{Cov}[\boldsymbol{X}_2\boldsymbol{\beta}_2, \boldsymbol{X}_1\boldsymbol{\beta}_1] &= \left(\frac{\gamma_g}{M}\boldsymbol{X}_1\boldsymbol{X}_2^\top\right)^\top = \frac{\gamma_g}{M}\boldsymbol{X}_2\boldsymbol{X}_1^\top, \\ \mathrm{Cov}[\boldsymbol{I}_1\boldsymbol{\epsilon}_1] &= \boldsymbol{I}_1\,\sigma_{e1}^2\boldsymbol{I}_N\,\boldsymbol{I}_1^\top = \sigma_{e1}^2\mathrm{diag}(\boldsymbol{E}_1), \\ \mathrm{Cov}[\boldsymbol{I}_2\boldsymbol{\epsilon}_2] &= \sigma_{e2}^2\mathrm{diag}(\boldsymbol{E}_2). \end{aligned}$$

Therefore,

$$\begin{aligned} \mathrm{Cov}[\boldsymbol{y}] &= \frac{\sigma_{g1}^2}{M}\boldsymbol{X}_1\boldsymbol{X}_1^\top + \frac{\sigma_{g2}^2}{M}\boldsymbol{X}_2\boldsymbol{X}_2^\top + \frac{\gamma_g}{M}\left(\boldsymbol{X}_1\boldsymbol{X}_2^\top + \boldsymbol{X}_2\boldsymbol{X}_1^\top\right) \\ &\quad + \sigma_{e1}^2\mathrm{diag}(\boldsymbol{E}_1) + \sigma_{e2}^2\mathrm{diag}(\boldsymbol{E}_2) \\ &= \sigma_{g1}^2\boldsymbol{K}_{\mathrm{gxe1}} + \sigma_{g2}^2\boldsymbol{K}_{\mathrm{gxe2}} + \gamma_g\,\boldsymbol{K}_{\mathrm{gxe12}} + \gamma_g\,\boldsymbol{K}_{\mathrm{gxe21}} + \sigma_{e1}^2\boldsymbol{K}_{\mathrm{nxe1}} + \sigma_{e2}^2\boldsymbol{K}_{\mathrm{nxe2}}, \end{aligned}$$

where the component kernels are defined as

$$\begin{aligned}
\boldsymbol{K}_{\text{gxe1}} &= \frac{1}{M}(\boldsymbol{X} \odot \boldsymbol{E}_1)(\boldsymbol{X} \odot \boldsymbol{E}_1)^\top, \\
\boldsymbol{K}_{\text{gxe2}} &= \frac{1}{M}(\boldsymbol{X} \odot \boldsymbol{E}_2)(\boldsymbol{X} \odot \boldsymbol{E}_2)^\top, \\
\boldsymbol{K}_{\text{gxe12}} &= \frac{1}{M}(\boldsymbol{X} \odot \boldsymbol{E}_1)(\boldsymbol{X} \odot \boldsymbol{E}_2)^\top, \\
\boldsymbol{K}_{\text{gxe21}} &= \frac{1}{M}(\boldsymbol{X} \odot \boldsymbol{E}_2)(\boldsymbol{X} \odot \boldsymbol{E}_1)^\top, \\
\boldsymbol{K}_{\text{nxe1}} &= (\boldsymbol{I}_N \odot \boldsymbol{E}_1)(\boldsymbol{I}_N \odot \boldsymbol{E}_1)^\top, \\
\boldsymbol{K}_{\text{nxe2}} &= (\boldsymbol{I}_N \odot \boldsymbol{E}_2)(\boldsymbol{I}_N \odot \boldsymbol{E}_2)^\top.
\end{aligned}$$

Define the symmetric cross-kernel

$$\boldsymbol{K}_{\text{gxe}}^{\text{sym}} = \boldsymbol{K}_{\text{gxe12}} + \boldsymbol{K}_{\text{gxe21}}.$$

The total phenotypic covariance can then be written as

$$\text{Cov}[\boldsymbol{y}] = \sigma_{g1}^2 \boldsymbol{K}_{\text{gxe1}} + \sigma_{g2}^2 \boldsymbol{K}_{\text{gxe2}} + \gamma_g \boldsymbol{K}_{\text{gxe}}^{\text{sym}} + \sigma_{e1}^2 \boldsymbol{K}_{\text{nxe1}} + \sigma_{e2}^2 \boldsymbol{K}_{\text{nxe2}}.$$

#### S2 Estimation of variance components via Method-of-Moments

We make the following observations:

- The environment-specific GRMs $\boldsymbol{K}_{\text{gxe1}}, \boldsymbol{K}_{\text{nxe1}}$ are supported only on individuals with $\boldsymbol{E}_i = 0$.
- The environment-specific GRMs $\boldsymbol{K}_{\text{gxe2}}, \boldsymbol{K}_{\text{nxe2}}$ are supported only on individuals with $\boldsymbol{E}_i = 1$.
- Since each individual belongs to exactly one environment, we have disjoint supports:

$$\boldsymbol{E}_1 \odot \boldsymbol{E}_2 = \mathbf{0},$$

and thus

$$\boldsymbol{K}_{\text{gxe1}}\boldsymbol{K}_{\text{gxe2}} = \mathbf{0}, \quad \boldsymbol{K}_{\text{gxe1}}\boldsymbol{K}_{\text{nxe2}} = \mathbf{0}, \quad \boldsymbol{K}_{\text{nxe1}}\boldsymbol{K}_{\text{nxe2}} = \mathbf{0}, \quad \text{etc.},$$

and in particular $\boldsymbol{K}^2_{\text{gxe12}} = \boldsymbol{K}^2_{\text{gxe21}} = \mathbf{0}$.

Therefore, many cross-kernel trace terms vanish and the matrix $\boldsymbol{T}$ becomes block diagonal:

$$\boldsymbol{T} = \begin{bmatrix} \operatorname{tr}(\boldsymbol{K}^2_{\text{gxe1}}) & 0 & 0 & \operatorname{tr}(\boldsymbol{K}_{\text{gxe1}}\boldsymbol{K}_{\text{nxe1}}) & 0 \\ 0 & \operatorname{tr}(\boldsymbol{K}^2_{\text{gxe2}}) & 0 & 0 & \operatorname{tr}(\boldsymbol{K}_{\text{gxe2}}\boldsymbol{K}_{\text{nxe2}}) \\ 0 & 0 & \operatorname{tr}(\boldsymbol{K}_{\text{gxe12}}\boldsymbol{K}_{\text{gxe21}}) & 0 & 0 \\ \operatorname{tr}(\boldsymbol{K}_{\text{gxe1}}\boldsymbol{K}_{\text{nxe1}}) & 0 & 0 & \operatorname{tr}(\boldsymbol{K}^2_{\text{nxe1}}) & 0 \\ 0 & \operatorname{tr}(\boldsymbol{K}_{\text{gxe2}}\boldsymbol{K}_{\text{nxe2}}) & 0 & 0 & \operatorname{tr}(\boldsymbol{K}^2_{\text{nxe2}}) \end{bmatrix},$$

and the right-hand side is

$$\boldsymbol{q} = \begin{bmatrix} \boldsymbol{y}^\top \boldsymbol{K}_{\text{gxe1}} \boldsymbol{y} \\ \boldsymbol{y}^\top \boldsymbol{K}_{\text{gxe2}} \boldsymbol{y} \\ \boldsymbol{y}^\top \boldsymbol{K}_{\text{gxe12}} \boldsymbol{y} \\ \boldsymbol{y}^\top \boldsymbol{K}_{\text{nxe1}} \boldsymbol{y} \\ \boldsymbol{y}^\top \boldsymbol{K}_{\text{nxe2}} \boldsymbol{y} \end{bmatrix}.$$

Thus, the normal equations decouple into two independent $2 \times 2$ linear systems for the environment-specific components and one scalar equation for the cross-environment covariance:

$$\begin{bmatrix} \operatorname{tr}(\boldsymbol{K}^2_{\text{gxe1}}) & \operatorname{tr}(\boldsymbol{K}_{\text{gxe1}}\boldsymbol{K}_{\text{nxe1}}) \\ \operatorname{tr}(\boldsymbol{K}_{\text{gxe1}}\boldsymbol{K}_{\text{nxe1}}) & \operatorname{tr}(\boldsymbol{K}^2_{\text{nxe1}}) \end{bmatrix} \begin{bmatrix} \sigma^2_{g1} \\ \sigma^2_{e1} \end{bmatrix} = \begin{bmatrix} \boldsymbol{y}^\top \boldsymbol{K}_{\text{gxe1}} \boldsymbol{y} \\ \boldsymbol{y}^\top \boldsymbol{K}_{\text{nxe1}} \boldsymbol{y} \end{bmatrix},$$

$$\begin{bmatrix} \operatorname{tr}(\boldsymbol{K}^2_{\text{gxe2}}) & \operatorname{tr}(\boldsymbol{K}_{\text{gxe2}}\boldsymbol{K}_{\text{nxe2}}) \\ \operatorname{tr}(\boldsymbol{K}_{\text{gxe2}}\boldsymbol{K}_{\text{nxe2}}) & \operatorname{tr}(\boldsymbol{K}^2_{\text{nxe2}}) \end{bmatrix} \begin{bmatrix} \sigma^2_{g2} \\ \sigma^2_{e2} \end{bmatrix} = \begin{bmatrix} \boldsymbol{y}^\top \boldsymbol{K}_{\text{gxe2}} \boldsymbol{y} \\ \boldsymbol{y}^\top \boldsymbol{K}_{\text{nxe2}} \boldsymbol{y} \end{bmatrix},$$

and, for the cross-environment covariance,

$$\gamma_g = \frac{\boldsymbol{y}^\top \boldsymbol{K}_{\text{gxe12}} \boldsymbol{y}}{\operatorname{tr}(\boldsymbol{K}_{\text{gxe12}}\boldsymbol{K}_{\text{gxe21}})}.$$

Here the symmetric kernel $\boldsymbol{K}^{\text{sym}}_{\text{gxe}}$ contributes a factor of two to both the numerator ($\boldsymbol{y}^\top \boldsymbol{K}^{\text{sym}}_{\text{gxe}} \boldsymbol{y} = 2\, \boldsymbol{y}^\top \boldsymbol{K}_{\text{gxe12}} \boldsymbol{y}$, since $\boldsymbol{K}_{\text{gxe21}} = \boldsymbol{K}^\top_{\text{gxe12}}$) and the diagonal entry ($\operatorname{tr}((\boldsymbol{K}^{\text{sym}}_{\text{gxe}})^2) = 2\operatorname{tr}(\boldsymbol{K}_{\text{gxe12}}\boldsymbol{K}_{\text{gxe21}})$, using $\boldsymbol{K}^2_{\text{gxe12}} = \boldsymbol{K}^2_{\text{gxe21}} = \mathbf{0}$), and these factors cancel.

Because the MoM estimator is unconstrained, individual variance estimates may be negative and $\hat{\rho}_g$ may exceed one in finite samples, particularly when a stratum-specific genetic variance is near zero. We therefore test hypotheses (for example, $H_0 : \rho_g = 1$) using Wald statistics rather than constraining the estimates to lie within their parameter space, and we omit $\hat{\rho}_g$ when a stratum-specific genetic variance is near zero, since $\rho_g$ is poorly identified at that boundary.

#### S3 Scale dependence of amplification and heterogeneity

Here we ask whether a monotone transformation of the phenotype can remove amplification, *i.e.*, make $\sigma^2_{g1} = \sigma^2_{g2}$ on the transformed scale or force the cross-environment genetic correlation to satisfy $\rho_g = 1$. The answer is different for the two quantities, and this asymmetry motivates the two-step classification used by REGENA. Additionally, it justifies the use of a Box-Cox transform to remove scale dependence.

Let $z_i = g(y_i)$ for a strictly increasing, differentiable $g$ applied to the pooled phenotype, as in the Box–Cox scan of Section S4. Within environment $a \in \{1, 2\}$, define the transformed genetic effect as the linear projection of $z$ onto the standardized genotypes,

$$\beta^z_{a,j} = \frac{\operatorname{Cov}\big(x_j,\, g(y) \mid E = a\big)}{\operatorname{Var}(x_j \mid E = a)} = \operatorname{Cov}\big(x_j,\, g(y) \mid E = a\big),$$

which is the natural object here because it is what a method-of-moments or REML estimator recovers whether or not $z$ is exactly linear-Gaussian in $\boldsymbol{X}$. In the polygenic limit implicit in the model ($M \to \infty$ with $\beta_{a,j} = O(1/\sqrt{M})$), $x_j$ and $y \mid E = a$ are jointly Gaussian by the central limit theorem, so Stein's lemma gives

$$\operatorname{Cov}\big(x_j,\, g(y) \mid E = a\big) = \kappa_a \operatorname{Cov}\big(x_j,\, y \mid E = a\big), \qquad \kappa_a := \mathbb{E}\big[\, g'(y) \mid E = a\,\big] > 0.$$

The scalar $\kappa_a$ does not depend on the variant index $j$: a monotone transformation rescales the entire per-SNP effect vector within an environment by a single positive scalar and never rotates it,

$$\boldsymbol{\beta}^z_1 = \kappa_1 \boldsymbol{\beta}_1, \qquad \boldsymbol{\beta}^z_2 = \kappa_2 \boldsymbol{\beta}_2.$$

Although a single $g$ is applied to all individuals, $\kappa_1 \neq \kappa_2$ in general, because the expectation is taken under the environment-specific marginal distribution of $y$.

We first note that amplification is removable. Since $\sigma^{2,z}_{g,a} = \kappa^2_a \sigma^2_{g,a}$, equalizing the transformed genetic variances requires the single scalar constraint

$$\frac{\kappa_1}{\kappa_2} = \frac{\mathbb{E}[g'(y) \mid E = 1]}{\mathbb{E}[g'(y) \mid E = 2]} = \sqrt{\frac{\sigma^2_{g2}}{\sigma^2_{g1}}}.$$

Because $\kappa_a$ is a functional of $g'$ against the environment-specific marginal distribution of $y$, this constraint can generically be satisfied whenever the two environments differ in their marginal phenotype distribution. Within the Box–Cox family $f_\lambda(y) = (y^\lambda - 1)/\lambda$ used here, the ratio $\kappa_1(\lambda)/\kappa_2(\lambda)$ varies continuously with $\lambda$ and, under mild conditions, sweeps through $\sqrt{\sigma^2_{g2}/\sigma^2_{g1}}$ at some $\lambda$ in the grid. The exception is degenerate: if $y \mid E = 1$ and $y \mid E = 2$ have identical marginal distributions then $\kappa_1 = \kappa_2$ for every $g$, and amplification can be neither created nor removed.

However, the cross-environment genetic correlation is scale invariant. The transformed variance components are

$$\sigma^{2,z}_{g1} = \kappa^2_1 \sigma^2_{g1}, \qquad \sigma^{2,z}_{g2} = \kappa^2_2 \sigma^2_{g2}, \qquad \gamma^z_g = \kappa_1 \kappa_2\, \gamma_g,$$

so that

$$\rho^z_g = \frac{\gamma^z_g}{\sqrt{\sigma^{2,z}_{g1} \sigma^{2,z}_{g2}}} = \frac{\kappa_1 \kappa_2\, \gamma_g}{\kappa_1 \kappa_2 \sqrt{\sigma^2_{g1} \sigma^2_{g2}}} = \rho_g$$

exactly, for any strictly increasing differentiable $g$. The correlation depends only on the relative direction of

$\boldsymbol{\beta}_1$ and $\boldsymbol{\beta}_2$, not on their magnitudes, and transformation only rescales these vectors. Consequently, if $\rho_g < 1$ on the measured scale, no monotone relabeling of the phenotype axis can produce $\rho_g^z = 1$.

This gives a precise version of an often-informal claim:

- **Amplification can reflect the measured phenotype scale.** Unequal genetic variance across environments largely reflects the local slope of the measurement scale at different phenotype means/variances in each environment, not necessarily a qualitative gene-by-environment effect. A suitable monotone transform (e.g. Box–Cox) can typically remove it whenever the two environments differ in their marginal phenotype distribution.

- **Effect heterogeneity is scale-free.** $\rho_g < 1$ reflects whether the same variants act proportionally the same way across environments (same direction of effect, regardless of magnitude), and no reparametrization of $y$ can manufacture or erase that. It is therefore a substantially stronger, scale-invariant signature of true gene-by-environment interaction than a difference between $\sigma_{g1}^2$ and $\sigma_{g2}^2$.

We note that the argument is exact under the joint-Gaussian (infinitesimal/polygenic) approximation already invoked for $(\beta_1, \beta_2)$ and $\epsilon$ in the model. For non-infinitesimal genetic architectures, where $x_j$ and $y$ are not exactly jointly Gaussian, the result is not exact but should continue to hold to leading order via the delta method.

**Choice of reporting scale for scale-independent pairs.** For scale-independent pairs we choose a scale on which to report, and that choice identifies the operative mechanism. We search the Box–Cox grid for a scale on which only one mechanism remains detectable: either the two environments share a common genetic variance, or their genetic effects are perfectly correlated. Whichever null is attained, the complementary mechanism is what survives and it is that mechanism we report. A scale on which the variances agree indicates that the apparent amplification is an artifact of the measurement scale and that effect heterogeneity is the operative signal; a scale on which the effects align perfectly indicates the converse. When no such scale exists within the window, both mechanisms are present under every transformation scanned, and we report on the measured scale ($\lambda = 1$). Because a transformation can attenuate the genetic signal to the point where neither mechanism is detectable, we exclude any scale on which the genetic variance is not significantly different from zero in both environments. Among the remaining candidates, we report the scale nearest the original, keeping the reported estimates as close to the measured trait as the mechanism permits.

#### S4 Valid region of Box-Cox transformations

Let $z_i(\lambda) = f_\lambda(y_i)$ and assume

$$z_i(\lambda) \overset{\text{iid}}{\sim} \mathcal{N}(\mu, \sigma^2).$$

By change of variables, the density of $y_i$ is

$$p(y_i \mid \mu, \sigma^2, \lambda) = p\big(z_i(\lambda) \mid \mu, \sigma^2\big) \left| \frac{d}{dy_i} f_\lambda(y_i) \right|.$$

For the Box–Cox transform on $y_i > 0$, $\left| \frac{d}{dy_i} f_\lambda(y_i) \right| = y_i^{\lambda-1}$, yielding the log-likelihood

$$\begin{aligned} \ell(\mu, \sigma^2, \lambda) &= \sum_{i=1}^{n} \log p(y_i \mid \mu, \sigma^2, \lambda) \\ &= -\frac{n}{2} \log(2w\sigma^2) - \frac{1}{2\sigma^2} \sum_{i=1}^{n} \big(z_i(\lambda) - \mu\big)^2 + (\lambda - 1) \sum_{i=1}^{n} \log y_i. \end{aligned}$$

Profiling out $\mu$ and $\sigma^2$ using the Gaussian MLEs

$$\hat{\mu}(\lambda) = \bar{z}(\lambda) = \frac{1}{n} \sum_{i=1}^{n} z_i(\lambda), \qquad \hat{\sigma}^2(\lambda) = \frac{1}{n} \sum_{i=1}^{n} \big(z_i(\lambda) - \bar{z}(\lambda)\big)^2,$$

gives the profile log-likelihood

$$\begin{aligned} \ell_p(\lambda) &= \ell\big(\hat{\mu}(\lambda), \hat{\sigma}^2(\lambda), \lambda\big) \\ &\propto -\frac{n}{2} \log\big(\hat{\sigma}^2(\lambda)\big) + (\lambda - 1) \sum_{i=1}^{n} \log y_i. \end{aligned}$$

We retain only $\lambda$ values whose profile log-likelihood lies within a fixed window of the maximum over the grid, thereby excluding transformations that yield implausible tail behavior.

#### S5 Robustness simulations

Here we give the generative model for each scenario used to assess the calibration of REGENA. In every scenario, the baseline phenotype follows the model

$$\boldsymbol{y}' = \boldsymbol{X}\boldsymbol{\beta} + (\boldsymbol{X} \odot \boldsymbol{E})\boldsymbol{\alpha} + (\boldsymbol{I}_N \odot \boldsymbol{E})\boldsymbol{\delta} + \boldsymbol{\epsilon},$$

where $\boldsymbol{\beta}$, $\boldsymbol{\alpha}$ and $\boldsymbol{\delta}$ are the additive, gene-by-environment and noise-by-environment effects.

**Phenotype correlated with the environment.** The simulated phenotype is $\boldsymbol{y} = \frac{r}{\sqrt{1-r^2}}\boldsymbol{E} + \boldsymbol{y}'$, where $r$ is the target correlation.

**Heritable environment.** The environment was itself simulated as a heritable trait, $\boldsymbol{E} = \boldsymbol{X}\boldsymbol{\beta}_E + \boldsymbol{\epsilon}_E$, with $\boldsymbol{\beta}_E \sim \mathcal{N}(\boldsymbol{0}, \frac{\sigma^2_{g,E}}{M}\boldsymbol{I})$ and $\boldsymbol{\epsilon}_E \sim \mathcal{N}(\boldsymbol{0}, (1-\sigma^2_{g,E})\boldsymbol{I})$, where $\sigma^2_{g,E}$ is the heritability of the environment and $M$ the number of SNPs, so that trait and environment share a genetic architecture.

**Shared causal variants.** Two further scenarios relaxed the assumption that the additive and G×E components act through distinct variants. In the first, the two components were assigned the same causal SNPs. In the second, the causal SNPs of the trait were additionally taken to be those of a heritable environment.

**Collider bias.** The phenotype was simulated to be correlated with a heritable environment through an unobserved confounder[50].

**Heavy-tailed noise.** The residual $\boldsymbol{\epsilon}$ was drawn from a Student's $t$ distribution with four degrees of freedom rather than a Gaussian.

#### S6 Relation with GENIE and equivalence under the null

In the G×E models underlying GxEMM[51] and GENIE[9], we consider the linear mixed model

$$\boldsymbol{y} = \boldsymbol{X}\boldsymbol{\alpha} + (\boldsymbol{X} \odot \boldsymbol{E}_1)\boldsymbol{\beta}_1 + (\boldsymbol{X} \odot \boldsymbol{E}_2)\boldsymbol{\beta}_2 + (\boldsymbol{I}_N \odot \boldsymbol{E}_1)\boldsymbol{\epsilon}_1 + (\boldsymbol{I}_N \odot \boldsymbol{E}_2)\boldsymbol{\epsilon}_2 + \boldsymbol{\delta},$$

where each variance component is independent and

$$\boldsymbol{\alpha} \sim \mathcal{N}(\boldsymbol{0}, \tfrac{\sigma_g^2}{M}\boldsymbol{I}_M), \quad \boldsymbol{\beta}_1 \sim \mathcal{N}(\boldsymbol{0}, \tfrac{\sigma_{gxe1}^2}{M}\boldsymbol{I}_M), \quad \boldsymbol{\beta}_2 \sim \mathcal{N}(\boldsymbol{0}, \tfrac{\sigma_{gxe2}^2}{M}\boldsymbol{I}_M),$$

$$\boldsymbol{\epsilon}_1 \sim \mathcal{N}(\boldsymbol{0}, \sigma_{nxe1}^2\boldsymbol{I}_N), \quad \boldsymbol{\epsilon}_2 \sim \mathcal{N}(\boldsymbol{0}, \sigma_{nxe2}^2\boldsymbol{I}_N), \quad \boldsymbol{\delta} \sim \mathcal{N}(\boldsymbol{0}, \sigma_e^2\boldsymbol{I}_N).$$

The corresponding covariance is

$$\mathrm{Cov}[\boldsymbol{y}] = \sigma_g^2\boldsymbol{K}_{\mathrm{gxe}}^{\mathrm{sym}} + (\sigma_g^2 + \sigma_{gxe1}^2)\boldsymbol{K}_{\mathrm{gxe1}} + (\sigma_g^2 + \sigma_{gxe2}^2)\boldsymbol{K}_{\mathrm{gxe2}} + (\sigma_e^2 + \sigma_{nxe1}^2)\boldsymbol{K}_{\mathrm{nxe1}} + (\sigma_e^2 + \sigma_{nxe2}^2)\boldsymbol{K}_{\mathrm{nxe2}}. \tag{S1}$$

Comparing with our model in Equation 1, we obtain the reparameterization

$$\sigma_g^2 + \sigma_{gxe1}^2 = \sigma_{g1}^2, \qquad \sigma_g^2 + \sigma_{gxe2}^2 = \sigma_{g2}^2, \qquad \sigma_e^2 + \sigma_{nxe1}^2 = \sigma_{e1}^2, \qquad \sigma_e^2 + \sigma_{nxe2}^2 = \sigma_{e2}^2,$$

together with $\gamma_g = \sigma_g^2$ when $\gamma_g > 0$. Thus GENIE corresponds to a special case of REGENA.

**Equivalence under the null.** Under the null in our model, $\rho_g = 1$ and $\sigma_{g1}^2 = \sigma_{g2}^2$, which together imply

$$\gamma_g \;=\; \rho_g\sqrt{\sigma_{g1}^2\sigma_{g2}^2} \;=\; \sigma_{g1}^2 \;=\; \sigma_{g2}^2.$$

Matching to the GENIE parametrization, where $\gamma_g = \sigma_g^2$ and $\sigma_{g1}^2 = \sigma_g^2 + \sigma_{gxe1}^2$, $\sigma_{g2}^2 = \sigma_g^2 + \sigma_{gxe2}^2$, we obtain $\sigma_{gxe1}^2 = \sigma_{gxe2}^2 = 0$, i.e. no G×E in GENIE. Conversely, $\sigma_{gxe1}^2 = \sigma_{gxe2}^2 = 0$ implies $\rho_g = 1$ and $\sigma_{g1}^2 = \sigma_{g2}^2$. The two models thus agree under the null.

**Imperfect correlation is detected by GENIE.** Under imperfect genetic correlation, $\rho_g < 1$ while $\sigma_{g1}^2 = \sigma_{g2}^2$. Since $\rho_g = \gamma_g/\sqrt{\sigma_{g1}^2\sigma_{g2}^2}$, equality of variances implies $\gamma_g < \sigma_{g1}^2$. Translating, $\sigma_g^2 < \sigma_g^2 + \sigma_{gxe1}^2$, so $\sigma_{gxe1}^2 > 0$ (and by symmetry $\sigma_{gxe2}^2 = \sigma_{gxe1}^2 > 0$). Imperfect genetic correlation therefore corresponds to a nonzero G×E variance component in GENIE.

**GENIE cannot represent amplification.** Under amplification, $\rho_g = 1$ but $\sigma_{g1}^2 \neq \sigma_{g2}^2$ (without loss of generality $\sigma_{g1}^2 > \sigma_{g2}^2$). Under GENIE,

$$\rho_g \;=\; \frac{\sigma_g^2}{\sqrt{\sigma_g^2 + \sigma_{gxe1}^2}\,\sqrt{\sigma_g^2 + \sigma_{gxe2}^2}}.$$

Setting $\rho_g = 1$ and squaring yields

$$\sigma_g^2(\sigma_{gxe1}^2 + \sigma_{gxe2}^2) + \sigma_{gxe1}^2\sigma_{gxe2}^2 \;=\; 0.$$

Because all terms are nonnegative, this forces $\sigma^2_{gxe1} = \sigma^2_{gxe2} = 0$, which in turn implies $\sigma^2_{g1} = \sigma^2_{g2} = \sigma^2_g$. GENIE therefore cannot represent amplification, whereas REGENA explicitly allows $\rho_g = 1$ with heterogeneous genetic variances across environments. We further used simulations to confirm that GENIE detects both the absence of G×E and G×E driven by imperfect genetic correlation, but that the GxE estimates collapse to the boundary (yielding zero or invalid negative estimates) under proportional amplification (Figure S9).

**Single-environment encoding.** An alternative GENIE-style parametrization uses a single environmental indicator,

$$\boldsymbol{y} = \boldsymbol{X}\boldsymbol{\alpha} + (\boldsymbol{X} \odot \boldsymbol{E})\boldsymbol{\beta} + (\boldsymbol{I}_N \odot \boldsymbol{E})\boldsymbol{\epsilon} + \boldsymbol{\delta},$$

with $\boldsymbol{\alpha} \sim \mathcal{N}(\boldsymbol{0}, \frac{\sigma^2_g}{M}\boldsymbol{I}_M)$, $\boldsymbol{\beta} \sim \mathcal{N}(\boldsymbol{0}, \frac{\sigma^2_{gxe}}{M}\boldsymbol{I}_M)$, $\boldsymbol{\epsilon} \sim \mathcal{N}(\boldsymbol{0}, \sigma^2_{nxe}\boldsymbol{I}_N)$, $\boldsymbol{\delta} \sim \mathcal{N}(\boldsymbol{0}, \sigma^2_e\boldsymbol{I}_N)$. The implied covariance,

$$\mathrm{Cov}[\boldsymbol{y}] = \sigma^2_g\boldsymbol{K}_{\mathrm{gxe1}} + (\sigma^2_g + \sigma^2_{gxe})\boldsymbol{K}_{\mathrm{gxe2}} + \sigma^2_g\boldsymbol{K}^{\mathrm{sym}}_{\mathrm{gxe}} + \sigma^2_e\boldsymbol{K}_{\mathrm{nxe1}} + (\sigma^2_e + \sigma^2_{nxe})\boldsymbol{K}_{\mathrm{nxe2}}, \tag{S2}$$

is a restricted special case of Equation 1 under $\sigma^2_g = \sigma^2_{g1}$, $\sigma^2_g + \sigma^2_{gxe} = \sigma^2_{g2}$, $\sigma^2_e = \sigma^2_{e1}$, $\sigma^2_e + \sigma^2_{nxe} = \sigma^2_{e2}$. This parameterization enforces that both genetic and environmental variances are larger in the environment encoded as 1 and is therefore not equivalent to the more flexible model in Equation 1.

#### S7 Generalization beyond the Box–Cox family

To test whether REGENA generalizes to transformations outside the Box–Cox family, we simulated phenotypes under six nonlinear maps spanning saturating, convex, and count-valued behavior: logistic, exponential, Michaelis–Menten, and softplus transformations, together with two count models (Gamma/negative-binomial and Poisson) in which the transformed value specifies the mean of a stochastic draw. For each map, we generated a latent phenotype without G×E, $\boldsymbol{y} = \boldsymbol{X}\boldsymbol{\beta} + \boldsymbol{\epsilon}$, with additive heritability 0.2 and genetic effects shared across environments, so that $\rho_g = 1$ and $\sigma^2_{g1} = \sigma^2_{g2}$ hold by construction on the latent scale. We then applied an environment-specific location offset $\Delta_T$ followed by the transformation, $\boldsymbol{z} = T(\boldsymbol{y} + \Delta_T \boldsymbol{E})$, and analyzed $\boldsymbol{z}$ with REGENA over a Box–Cox scan of 51 values of $\lambda \in [-2, 3]$, using 50 replicates per map and UK Biobank array genotypes.

The offset is what generates the artifact: it places the two environments on differently curved stretches of the same map, so that identical latent genetic variation is compressed or stretched by different amounts on the observed scale (Figure S4). The ratio of the local slopes of $T$ between environments ranges from 0.79 (logistic, compressing) to 2.72 (Poisson, stretching). All six maps induce apparent amplification G×E on the observed scale: at $\lambda = 1$, G×E is detected in 100% of replicates for every transformation. The artifact is removed on the reconstructed scale, with detection proportion 2–8% covering the nominal 5% threshold (Figure S5 and S6). The exponential map serves as an in-family positive control, since $\lambda = 0$ inverts it exactly, and REGENA recovers a median $\lambda^* = 0.1$. The remaining recovered scales fall well inside the scan window (median $\lambda^* = 1.6$, 1.6, 0.3, 0.0 and $-0.6$ for the logistic, Michaelis–Menten, softplus, Gamma and Poisson maps). Box–Cox scan, however, cannot remove all forms of scale-induced G×E. Under a strongly saturating Michaelis–Menten map, G×E remains significant in 100% of replicates, and the recovered $\lambda^*$ is pinned to the scan endpoint in every replicate (Figure S7).

#### S8 Definition of G×E heritability

Let $\boldsymbol{\beta}_1, \boldsymbol{\beta}_2 \in \mathbb{R}^M$ denote the SNP-effect vectors in the unexposed ($E = 0$) and exposed ($E = 1$) environments, respectively. Their per-SNP variances are $\sigma_{g1}^2/M$ and $\sigma_{g2}^2/M$, and their covariance is $\gamma_g/M$. Let $w = \Pr(E = 0)$ denote the population prevalence of the unexposed environment, such that $1 - w = \Pr(E = 1)$ is the prevalence of the exposed environment. For each SNP, define the prevalence-weighted mean effect and the deviations from this mean in the two environments as

$$\mu_j = w\beta_{1j} + (1 - w)\beta_{2j}, \qquad \Delta_{1j} = \beta_{1j} - \mu_j, \qquad \Delta_{2j} = \beta_{2j} - \mu_j.$$

Writing $d_j = \beta_{2j} - \beta_{1j}$, the deviations can equivalently be expressed as $\Delta_{1j} = -(1 - w)d_j, \Delta_{2j} = wd_j$. It follows that $w\Delta_{1j} + (1 - w)\Delta_{2j} = 0$ for every realization of the SNP effects. Thus, the deviations are prevalence-weighted mean zero by construction. Define

$$\beta_j(E) = \begin{cases} \beta_{1j}, & E = 0, \\ \beta_{2j}, & E = 1, \end{cases} \qquad \Delta_j(E) = \begin{cases} \Delta_{1j}, & E = 0, \\ \Delta_{2j}, & E = 1. \end{cases}$$

The SNP effect in environment $E$ can then be written as

$$\beta_j(E) = \mu_j + \Delta_j(E).$$

Here, $\mu_j$ represents the population-average SNP effect, whereas $\Delta_j(E)$ represents the environment-specific departure from this average. Taking the phenotype to have unit variance in the target population, we define the additive heritability as the variance of the prevalence-weighted mean effect:

$$\begin{aligned} h_g^2 &= M \operatorname{Var}(\mu_j) \\ &= w^2\sigma_{g1}^2 + (1 - w)^2\sigma_{g2}^2 + 2w(1 - w)\gamma_g. \end{aligned}$$

We define the G×E heritability as the population-average variance of the environment-specific deviations:

$$\begin{aligned} h_{G\times E}^2 &= M \left[w \operatorname{Var}(\Delta_{1j}) + (1 - w) \operatorname{Var}(\Delta_{2j})\right] \\ &= w(1 - w)\left(\sigma_{g1}^2 + \sigma_{g2}^2 - 2\gamma_g\right). \end{aligned}$$

The second equality follows from

$$\begin{aligned} &w \operatorname{Var}(\Delta_{1j}) + (1 - w) \operatorname{Var}(\Delta_{2j}) \\ &\qquad = w(1 - w)^2\operatorname{Var}(d_j) + (1 - w)w^2\operatorname{Var}(d_j) \\ &\qquad = w(1 - w)\operatorname{Var}(d_j), \end{aligned}$$

together with

$$M \operatorname{Var}(d_j) = M \operatorname{Var}(\beta_{2j} - \beta_{1j}) = \sigma_{g1}^2 + \sigma_{g2}^2 - 2\gamma_g.$$

Thus, the population-level contribution of G×E depends on both the magnitude of the difference in genetic effects between environments and the prevalence of the two environments. Because the environment-specific deviations are prevalence-weighted mean zero, the mean and deviation components yield the variance decomposition

$$h_g^2 + h_{G\times E}^2 = w\sigma_{g1}^2 + (1-w)\sigma_{g2}^2,$$

which is the prevalence-weighted genetic variance in the population. When the two environments are equally prevalent, $w = 1/2$, we have

$$\mu_j = \frac{\beta_{1j} + \beta_{2j}}{2}, \qquad \Delta_{1j} = -\frac{\beta_{2j} - \beta_{1j}}{2}, \qquad \Delta_{2j} = \frac{\beta_{2j} - \beta_{1j}}{2},$$

and the heritability components reduce to

$$h_g^2 = \frac{\sigma_{g1}^2 + \sigma_{g2}^2 + 2\gamma_g}{4}, \qquad h_{G\times E}^2 = \frac{\sigma_{g1}^2 + \sigma_{g2}^2 - 2\gamma_g}{4}.$$

In addition, let $\sigma_{g1} = \sqrt{\sigma_{g1}^2}, \sigma_{g2} = \sqrt{\sigma_{g2}^2}$. Adding and subtracting $2\sigma_{g1}\sigma_{g2}$ decomposes the G×E heritability into amplification and effect-heterogeneity components:

$$h_{G\times E}^2 = w(1-w)\left(\sigma_{g1}^2 + \sigma_{g2}^2 - 2\gamma_g\right) = \underbrace{w(1-w)\left(\sigma_{g1} - \sigma_{g2}\right)^2}_{h^2_{\text{gxe,amp}}} + \underbrace{2w(1-w)\sigma_{g1}\sigma_{g2}(1-\rho_g)}_{h^2_{\text{gxe,het}}}.$$

The amplification component captures differences in the magnitudes of genetic effects between environments. It vanishes when $\sigma_{g1} = \sigma_{g2}$ and accounts for all G×E heritability when the genetic correlation is one. The effect-heterogeneity component captures differences in the relative SNP effects, as measured by imperfect genetic correlation. It vanishes when $\rho_g = 1$ and accounts for all G×E heritability when the two genetic standard deviations are equal.

#### S9 Specificity and its decomposition

We define total specificity as the fraction of the population genetic variance attributable to environment-specific deviations:

$$S = \frac{h^2_{G\times E}}{h^2_g + h^2_{G\times E}} = \frac{w(1-w)\left(\sigma^2_{g1} + \sigma^2_{g2} - 2\gamma_g\right)}{w\sigma^2_{g1} + (1-w)\sigma^2_{g2}}.$$

Thus, specificity depends not only on the difference in genetic effects between the two environments, but also on their relative prevalence in the population. Note that when $\sigma^2_{g1} = \sigma^2_{g2} = \sigma^2_g$, we have $S = 2w(1-w)(1-\rho_g)$ so that $\rho_g \approx 1$ implies $S \approx 0$ with specificity increasing as $\rho_g$ decreases to 0. In addition, if we define per-allele effect size squared $\boldsymbol{\beta} = \frac{h^2}{2Mf(1-f)}$, then specificity $S = \frac{h^2_{G\times E}}{h^2_g + h^2_{G\times E}} = \frac{\beta^2_{\mathrm{G\times E}}}{\beta^2_g + \beta^2_{\mathrm{G\times E}}}$ can be expressed as a function of per-allele effect size squared as well.

Decompose total specificity into amplification and effect heterogeneity components:

$$S_{\mathrm{amp}} = \frac{w(1-w)\left(\sigma_{g1} - \sigma_{g2}\right)^2}{w\sigma^2_{g1} + (1-w)\sigma^2_{g2}},$$

and

$$S_{\mathrm{het}} = \frac{2w(1-w)\sigma_{g1}\sigma_{g2}(1-\rho_g)}{w\sigma^2_{g1} + (1-w)\sigma^2_{g2}}.$$

By construction, $S_{\mathrm{amp}} + S_{\mathrm{het}} = S$. When the two environments are equally prevalent, $w = 1/2$, these expressions reduce to

$$S_{\mathrm{amp}} = \frac{\left(\sigma_{g1} - \sigma_{g2}\right)^2}{2\left(\sigma^2_{g1} + \sigma^2_{g2}\right)}, \qquad S_{\mathrm{het}} = \frac{\sigma_{g1}\sigma_{g2}(1-\rho_g)}{\sigma^2_{g1} + \sigma^2_{g2}},$$

recovering the equal-prevalence definitions.

#### S10 Multi-component G×E model

We extend the environment-specific linear mixed model by partitioning the genotype matrix into $K$ non-overlapping annotations. Let $\boldsymbol{X}_k \in \mathbb{R}^{N \times M_k}$ denote the standardized genotype matrix for annotation $k$. The phenotype is modeled as

$$\boldsymbol{y} = \sum_{k=1}^{K} [(\boldsymbol{X}_k \odot \boldsymbol{E}_1)\boldsymbol{\beta}_{1k} + (\boldsymbol{X}_k \odot \boldsymbol{E}_2)\boldsymbol{\beta}_{2k}] + (\boldsymbol{I}_N \odot \boldsymbol{E}_1)\boldsymbol{\epsilon}_1 + (\boldsymbol{I}_N \odot \boldsymbol{E}_2)\boldsymbol{\epsilon}_2.$$

We assume independent effects across annotations, with

$$\begin{bmatrix} \boldsymbol{\beta}_{1k} \\ \boldsymbol{\beta}_{2k} \end{bmatrix} \sim \mathcal{N}\left(\boldsymbol{0}, \frac{1}{M_k}\begin{bmatrix} \sigma^2_{g1,k}\boldsymbol{I}_{M_k} & \gamma_{g,k}\boldsymbol{I}_{M_k} \\ \gamma_{g,k}\boldsymbol{I}_{M_k} & \sigma^2_{g2,k}\boldsymbol{I}_{M_k} \end{bmatrix}\right), \quad \begin{bmatrix} \boldsymbol{\epsilon}_1 \\ \boldsymbol{\epsilon}_2 \end{bmatrix} \sim \mathcal{N}\left(\boldsymbol{0}, \begin{bmatrix} \sigma^2_{e1}\boldsymbol{I}_N & \boldsymbol{0} \\ \boldsymbol{0} & \sigma^2_{e2}\boldsymbol{I}_N \end{bmatrix}\right).$$

The phenotypic covariance becomes a sum over annotation-specific components:

$$\mathrm{Cov}[\boldsymbol{y}] = \sum_{k=1}^{K} \left(\sigma^2_{g1,k}\boldsymbol{K}_{\mathrm{gxe1},k} + \sigma^2_{g2,k}\boldsymbol{K}_{\mathrm{gxe2},k} + \gamma_{g,k}\boldsymbol{K}^{\mathrm{sym}}_{\mathrm{gxe},k}\right) + \sigma^2_{e1}\boldsymbol{K}_{\mathrm{nxe1}} + \sigma^2_{e2}\boldsymbol{K}_{\mathrm{nxe2}},$$

where

$$\begin{aligned} \boldsymbol{K}_{\mathrm{gxe1},k} &= \frac{1}{M_k}(\boldsymbol{X}_k \odot \boldsymbol{E}_1)(\boldsymbol{X}_k \odot \boldsymbol{E}_1)^\top, \\ \boldsymbol{K}_{\mathrm{gxe2},k} &= \frac{1}{M_k}(\boldsymbol{X}_k \odot \boldsymbol{E}_2)(\boldsymbol{X}_k \odot \boldsymbol{E}_2)^\top, \\ \boldsymbol{K}^{\mathrm{sym}}_{\mathrm{gxe},k} &= \frac{1}{M_k}\left[(\boldsymbol{X}_k \odot \boldsymbol{E}_1)(\boldsymbol{X}_k \odot \boldsymbol{E}_2)^\top + (\boldsymbol{X}_k \odot \boldsymbol{E}_2)(\boldsymbol{X}_k \odot \boldsymbol{E}_1)^\top\right]. \end{aligned}$$

For annotation $k$, the genetic correlation is

$$\rho_{g,k} = \frac{\gamma_{g,k}}{\sqrt{\sigma^2_{g1,k}\sigma^2_{g2,k}}}.$$

#### S11 Categorical environment encoding

Let $\boldsymbol{E}_1, \ldots, \boldsymbol{E}_K \in \{0,1\}^N$ denote one-hot indicators for a categorical environment with $K$ mutually exclusive categories. Define $\boldsymbol{X}_k = \boldsymbol{X} \odot \boldsymbol{E}_k$ and $\boldsymbol{I}_k = \boldsymbol{I}_N \odot \boldsymbol{E}_k$. We model

$$\boldsymbol{y} = \sum_{k=1}^{K} \boldsymbol{X}_k \boldsymbol{\beta}_k + \sum_{k=1}^{K} \boldsymbol{I}_k \boldsymbol{\varepsilon}_k,$$

with

$$\begin{bmatrix} \boldsymbol{\beta}_1 \\ \vdots \\ \boldsymbol{\beta}_K \end{bmatrix} \sim \mathcal{N}\left(0, \frac{1}{M} \boldsymbol{\Sigma}_g \otimes \boldsymbol{I}_M\right), \qquad \begin{bmatrix} \boldsymbol{\varepsilon}_1 \\ \vdots \\ \boldsymbol{\varepsilon}_K \end{bmatrix} \sim \mathcal{N}\big(0, \mathrm{diag}(\sigma_{e,1}^2 \boldsymbol{I}_N, \ldots, \sigma_{e,K}^2 \boldsymbol{I}_N)\big),$$

where $\boldsymbol{\Sigma}_g$ is symmetric positive semidefinite with diagonal entries $\sigma_{g,k}^2$ and off-diagonal entries $\gamma_{g,k\ell}$.

Writing $\boldsymbol{K}_{k\ell} = \frac{1}{M}(\boldsymbol{X} \odot \boldsymbol{E}_k)(X \odot \boldsymbol{E}_\ell)^\top$, $N_k = (\boldsymbol{I}_N \odot \boldsymbol{E}_k)(\boldsymbol{I}_N \odot \boldsymbol{E}_k)^\top$, and $\boldsymbol{K}_{k\ell}^{\mathrm{sym}} = \boldsymbol{K}_{k\ell} + \boldsymbol{K}_{\ell k}$ for $k < \ell$, the phenotypic covariance is

$$\mathrm{Cov}(\boldsymbol{y}) = \sum_{k=1}^{K} \sigma_{g,k}^2 \boldsymbol{K}_{kk} + \sum_{1 \le k < \ell \le K} \gamma_{g,k\ell} \boldsymbol{K}_{k\ell}^{\mathrm{sym}} + \sum_{k=1}^{K} \sigma_{e,k}^2 N_k.$$

The pairwise genetic correlation is $\rho_{g,k\ell} = \gamma_{g,k\ell} / \sqrt{\sigma_{g,k}^2 \sigma_{g,\ell}^2}$ for $1 \le k < \ell \le K$.

#### S12 Polygenic score construction

Recall $\boldsymbol{X}_1 = \boldsymbol{X} \odot \boldsymbol{E}_1$, $\boldsymbol{X}_2 = \boldsymbol{X} \odot \boldsymbol{E}_2$, and $\boldsymbol{I}_1 = \boldsymbol{I}_N \odot \boldsymbol{E}_1$, $\boldsymbol{I}_2 = \boldsymbol{I}_N \odot \boldsymbol{E}_2$. We re-write the environment-specific linear mixed model as:

$$\boldsymbol{y} = \boldsymbol{X}_1\boldsymbol{\beta}_1 + \boldsymbol{X}_2\boldsymbol{\beta}_2 + \boldsymbol{I}_1\boldsymbol{\epsilon}_1 + \boldsymbol{I}_2\boldsymbol{\epsilon}_2,$$

define the stacked design matrix and coefficients

$$\boldsymbol{X}_{\text{blk}} = \begin{bmatrix}\boldsymbol{X}_1 & \boldsymbol{X}_2\end{bmatrix}, \qquad \boldsymbol{\beta} = \begin{bmatrix}\boldsymbol{\beta}_1 \\ \boldsymbol{\beta}_2\end{bmatrix}, \qquad \boldsymbol{\eta} = \boldsymbol{I}_1\boldsymbol{\epsilon}_1 + \boldsymbol{I}_2\boldsymbol{\epsilon}_2,$$

so that $\boldsymbol{y} = \boldsymbol{X}_{\text{blk}}\boldsymbol{\beta} + \boldsymbol{\eta}$.

We rewrite the prior distribution on the environment-specific SNP effect sizes as:

$$\boldsymbol{\beta} \sim \mathcal{N}(\boldsymbol{0}, \boldsymbol{\Sigma}_\beta), \qquad \boldsymbol{\Sigma}_\beta = \frac{1}{M}\begin{bmatrix}\sigma_{g1}^2\boldsymbol{I}_M & \gamma_g\boldsymbol{I}_M \\ \gamma_g\boldsymbol{I}_M & \sigma_{g2}^2\boldsymbol{I}_M\end{bmatrix},$$

and independent noise $\boldsymbol{\eta} \sim \mathcal{N}(\boldsymbol{0}, \boldsymbol{\Sigma}_e)$ with

$$\boldsymbol{\Sigma}_e = \sigma_{e1}^2 \, \mathsf{diag}\,(\boldsymbol{E}_1) + \sigma_{e2}^2 \, \mathsf{diag}\,(\boldsymbol{E}_2)\,.$$

Then $\boldsymbol{y} \sim \mathcal{N}(\boldsymbol{0}, \boldsymbol{U})$ with $\boldsymbol{U} = \boldsymbol{X}_{\text{blk}}\boldsymbol{\Sigma}_\beta\boldsymbol{X}_{\text{blk}}^\top + \boldsymbol{\Sigma}_e$. By Gaussian conditioning,

$$\widehat{\boldsymbol{\beta}} = \mathbb{E}[\boldsymbol{\beta} \mid \boldsymbol{y}] = \boldsymbol{\Sigma}_\beta\boldsymbol{X}_{\text{blk}}^\top\boldsymbol{U}^{-1}\boldsymbol{y}.$$

Letting $\boldsymbol{\alpha} = \boldsymbol{U}^{-1}\boldsymbol{y}$ and expanding the block multiplication:

$$\widehat{\boldsymbol{\beta}}_1 = \frac{\sigma_{g1}^2}{M}\boldsymbol{X}_1^\top\boldsymbol{\alpha} + \frac{\gamma_g}{M}\boldsymbol{X}_2^\top\boldsymbol{\alpha},$$
$$\widehat{\boldsymbol{\beta}}_2 = \frac{\gamma_g}{M}\boldsymbol{X}_1^\top\boldsymbol{\alpha} + \frac{\sigma_{g2}^2}{M}\boldsymbol{X}_2^\top\boldsymbol{\alpha}.$$

The computational bottleneck is applying $\boldsymbol{U}^{-1}$, i.e., solving $\boldsymbol{U}\boldsymbol{\alpha} = \boldsymbol{y}$ at biobank scale. We solve this system using preconditioned conjugate gradients, which only requires repeated matrix–vector products $\boldsymbol{v} \mapsto \boldsymbol{U}\boldsymbol{v}$ together with a cheap preconditioner solve. We use the block-diagonal preconditioner

$$\boldsymbol{M} = \begin{bmatrix}(\sigma_{g1}^2 + \sigma_{e1}^2)\boldsymbol{I} & \boldsymbol{0} \\ \boldsymbol{0} & (\sigma_{g2}^2 + \sigma_{e2}^2)\boldsymbol{I}\end{bmatrix},$$

which makes $\boldsymbol{M}^{-1}\boldsymbol{r}$ an elementwise rescaling of residuals within each stratum, substantially reducing the number of PCG iterations in practice.

**Prediction and evaluation.** After fitting the PRS weights on the training subset, we evaluate predictive performance by comparing two linear models on a held-out calibration or test set:

$$\text{Model 1:} \qquad y \sim \text{covariates (+ ENV)},$$

$$\text{Model 2:} \qquad y \sim \text{covariates (+ ENV)} + \text{PRS}.$$

Here the environment indicator is optionally included as an additional regression covariate. We report

$$\Delta R^2 = R^2(\text{covariates} + \text{PRS}) - R^2(\text{covariates}),$$

both overall and within each environment stratum. For the stratified PRS model, the PRS supplied to the regression is environment-specific, i.e., each individual is scored using $\widehat{\boldsymbol{\beta}}_1$ or $\widehat{\boldsymbol{\beta}}_2$ according to their observed environment stratum.

**Bootstrap uncertainty for prediction metrics.** To quantify uncertainty in predictive performance, we compute nonparametric bootstrap standard errors for $R^2$(covariates), $R^2$(covariates + PRS), and $\Delta R^2$ after the PRS has been trained. Specifically, on the aligned evaluation set we resample individuals with replacement, recompute the two $R^2$ values and their difference, and use the empirical standard deviation across bootstrap replicates as the standard error. We also report percentile bootstrap confidence intervals for $\Delta R^2$. This procedure treats the trained PRS weights as fixed and therefore quantifies uncertainty in the downstream evaluation step rather than uncertainty from refitting the full genetic model.

#### S13 Environment-stratified GWAS

For each of the 20 scale-independent trait–environment pairs, we performed three genome-wide association scans using common array genotypes: a pooled scan including all individuals with non-missing phenotype and environment measurements, and separate scans within each of the two environment strata. All scans used the phenotype at the optimal scale selected by the pipeline. In the pooled scan, the environment was included as an additional covariate, such that genotype effect estimates were adjusted for the environment main effect.

All locus counts refer to LD-clumped independent loci rather than individual SNPs. We performed clumping with the following criteria. An index SNP was required to reach genome-wide significance ($p < 5 \times 10^{-8}$), and SNPs were included in an existing clump only if they also reached genome-wide significance. SNPs were assigned to a clump if they were within 250 kb of the index SNP and had $r^2 > 0.1$ with the index SNP. For all scans, including the environment-stratified scans, LD was estimated in the pooled sample for the corresponding trait–environment pair, ensuring that differences in locus counts reflected differences in association signals rather than differences in the LD reference set. A lead SNP from an environment-stratified scan was considered to define a novel locus if the pooled scan contained no SNP with $p < 5 \times 10^{-8}$ within $\pm$250 kb of the lead SNP; otherwise, the locus was classified as shared. Each novel locus was annotated to its nearest gene.

### Supplementary Tables

Table S1: **Trait names and abbreviations.** Full trait name, UK Biobank field ID and the abbreviation used throughout the figures and tables, for the 43 quantitative traits.

| Abbrev. | Trait | UKB field |
|---|---|---|
| BMI | Body mass index | 21001 |
| BMR | Basal metabolic rate | 23105 |
| Height | Height | 50 |
| WHR | Waist-hip ratio | 48/49 |
| EOS | Eosinophil count | 30150 |
| HLSR | High light scatter reticulocyte count | 30300 |
| LYM | Lymphocyte count | 30120 |
| MCH | Mean corpuscular hemoglobin | 30050 |
| MON | Monocyte count | 30130 |
| MPV | Mean platelet volume | 30100 |
| MSCV | Mean sphered cell volume | 30270 |
| PDW | Platelet distribution width | 30110 |
| PLT | Platelet count | 30080 |
| RBC | RBC count | 30010 |
| RDW | RBC distribution width | 30070 |
| Testo | Testosterone | 30850 |
| WBC | White blood cell count | 30000 |
| ALP | Alkaline phosphatase | 30610 |
| Ca | Calcium | 30680 |
| ApoA1 | Apolipoprotein A | 30630 |
| CRP | C-reactive protein | 30710 |
| DBP | Diastolic blood pressure | 4079 |
| HDL-C | HDL cholesterol | 30760 |
| LDL-C | LDL cholesterol direct | 30780 |
| SBP | Systolic blood pressure | 4080 |
| TC | Cholesterol | 30690 |
| TG | Triglycerides | 30870 |
| GLU | Glucose | 30740 |
| HbA1c | Hemoglobin A1c | 30750 |
| ALB | Albumin | 30600 |
| ALT | Alanine aminotransferase | 30620 |
| AST | Aspartate aminotransferase | 30650 |
| GGT | Gamma glutamyltransferase | 30730 |
| $FEV_1$/FVC | FEV1-FVC ratio | 20258 |
| FVC | FVC | 20257 |
| CysC | Cystatin-C | 30720 |
| Phosphate | Phosphate | 30810 |
| SCr | Creatinine | 30700 |
| SU | Urate | 30880 |
| uCr | Creatinine in urine | 30510 |
| uK | Potassium in urine | 30520 |
| uNa | Sodium in urine | 30530 |
| Urea | Urea | 30670 |

Table S2: **Novel context-specific loci per trait–environment pair.** For each of the 20 scale-independent pairs: pooled and per-stratum sample sizes, the number of independent genome-wide-significant loci in the stratified scan, the number of those that are novel (no pooled-scan signal within ±250 kb), and the number of novel loci with opposite-sign effects across strata. Trait abbreviations are defined in Table S1.

| Trait | Environment | $N_{\text{pool}}$ | $N_1$ | $N_2$ | Strat. loci | Novel | Opp. sign |
|---|---|---|---|---|---|---|---|
| BMI | Age (younger/older) | 290,277 | 149,388 | 140,889 | 109 | 5 | 0 |
| DBP | Age (younger/older) | 272,062 | 139,486 | 132,576 | 50 | 3 | 0 |
| BMI | Alcohol (light/heavy) | 231,135 | 172,628 | 58,507 | 69 | 6 | 2 |
| WHR | Alcohol (light/heavy) | 231,346 | 172,769 | 58,577 | 39 | 0 | 0 |
| SBP | Insomnia (none/sympt.) | 271,796 | 65,626 | 206,170 | 46 | 1 | 0 |
| HbA1c | Insomnia (none/sympt.) | 277,158 | 66,945 | 210,213 | 130 | 7 | 1 |
| uNa | Insomnia (none/sympt.) | 281,961 | 68,364 | 213,597 | 9 | 0 | 0 |
| ALT$^{\dagger}$ | Sex (M/F) | 277,507 | 129,441 | 148,066 | 113 | 4 | 1 |
| RBC | Sex (M/F) | 282,512 | 132,017 | 150,495 | 255 | 5 | 2 |
| BMI | Sex (M/F) | 290,277 | 135,315 | 154,962 | 97 | 7 | 0 |
| SBP | Sex (M/F) | 272,058 | 126,986 | 145,072 | 30 | 1 | 0 |
| SCr$^{\dagger}$ | Sex (M/F) | 277,464 | 129,463 | 148,001 | 136 | 4 | 0 |
| $FEV_1$/FVC | Sex (M/F) | 234,859 | 105,561 | 129,298 | 118 | 4 | 0 |
| GGT | Sex (M/F) | 277,469 | 129,449 | 148,020 | 70 | 4 | 0 |
| Testo$^{\dagger}$ | Sex (M/F) | 251,626 | 128,287 | 123,339 | 155 | 58 | 32 |
| WHR$^{\dagger}$ | Sex (M/F) | 290,661 | 135,523 | 155,138 | 97 | 21 | 12 |
| $FEV_1$/FVC | Smoking (never/ever) | 234,859 | 129,156 | 105,703 | 132 | 8 | 0 |
| DBP | Statin (non-user/user) | 272,062 | 227,920 | 44,142 | 95 | 13 | 8 |
| HbA1c | Statin (non-user/user) | 277,424 | 232,455 | 44,969 | 207 | 23 | 7 |
| BMI | Deprivation (less/more) | 231,135 | 115,650 | 115,485 | 55 | 4 | 2 |
| **Total** | | | | | **2,012** | **178** | **67** |

Table S3: **Contrast of the baseline and G×E polygenic scores for the five trait–environment pairs with the largest gains in incremental $R^2$.** We rank the $n$ held-out individuals by each score and compute the mean covariate-adjusted phenotype – residualized on the covariates and the environment indicator, in SD units of the phenotype – among the $k = \lceil 0.01\, n \rceil$ individuals in each tail. $\Delta$ is the difference between the top and bottom 1% for a given score, and the last column reports how much larger this contrast is for the G×E score than for the baseline, $\Delta_{\mathrm{G\times E}} - \Delta_{\mathrm{baseline}}$. Standard errors (in parentheses) follow from the within-tail sampling variances.

| | | | Baseline PGS | | | G×E PGS | | | |
|---|---|---|---|---|---|---|---|---|---|
| Trait × environment | $n$ | $k$ | Top 1% | Bottom 1% | $\Delta_{\mathrm{baseline}}$ | Top 1% | Bottom 1% | $\Delta_{\mathrm{G\times E}}$ | $\Delta_{\mathrm{G\times E}} - \Delta_{\mathrm{baseline}}$ |
| Red blood cell count × Sex | 56,503 | 566 | 0.594 | −0.626 | 1.220 (0.049) | 0.865 | −0.798 | 1.664 (0.056) | 0.443 (0.074) |
| Creatinine × Sex | 55,493 | 555 | 0.491 | −0.533 | 1.025 (0.047) | 0.724 | −0.710 | 1.433 (0.055) | 0.409 (0.073) |
| Testosterone × Sex | 50,326 | 504 | 0.169 | −0.102 | 0.271 (0.039) | 0.373 | −0.304 | 0.676 (0.064) | 0.405 (0.075) |
| Waist-to-hip ratio × Sex | 58,133 | 582 | 0.364 | −0.336 | 0.700 (0.046) | 0.579 | −0.525 | 1.103 (0.057) | 0.404 (0.073) |
| Waist-to-hip ratio × Alcohol | 46,261 | 463 | 0.333 | −0.328 | 0.661 (0.051) | 0.536 | −0.470 | 1.006 (0.065) | 0.346 (0.083) |

### Supplementary Figures

#### Computational scalability

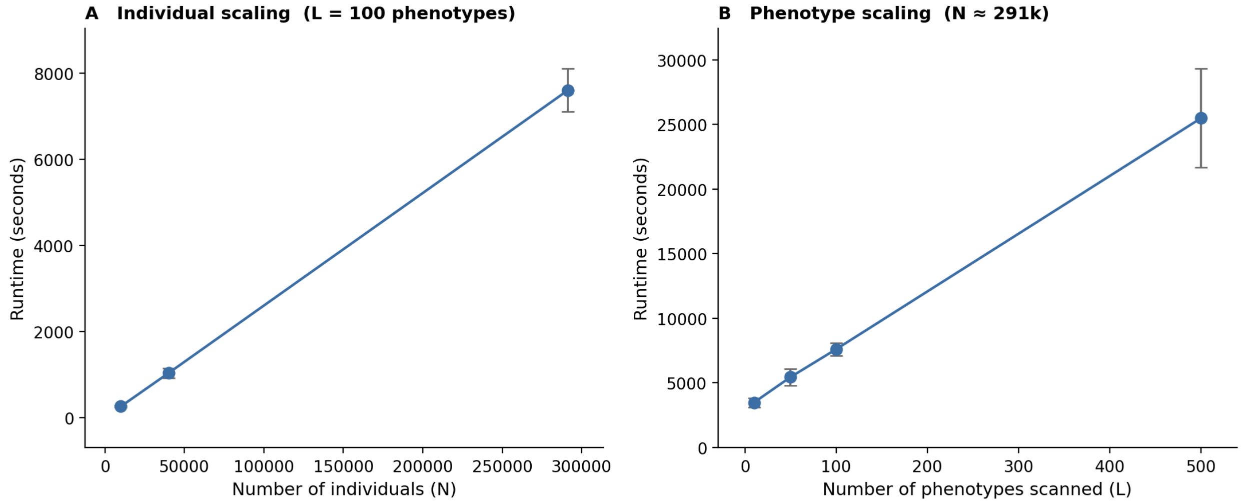

Figure S1: **Computational scalability of REGENA for biobank-scale analyses.** Wall-clock runtime was benchmarked using UK Biobank array SNPs from unrelated White British individuals ($M = 454{,}207$), with $B = 10$ random vectors and 100 jackknife blocks. Each configuration was evaluated across ten replicates; points indicate the mean runtime and error bars indicate the standard deviation. **a,** Runtime as a function of sample size $N$, with the number of phenotypes fixed at $L = 100$. **b,** Runtime as a function of the number of phenotypes $L$, using the full sample of $N = 291{,}273$ individuals.

#### Simulations: Box–Cox scan under the null

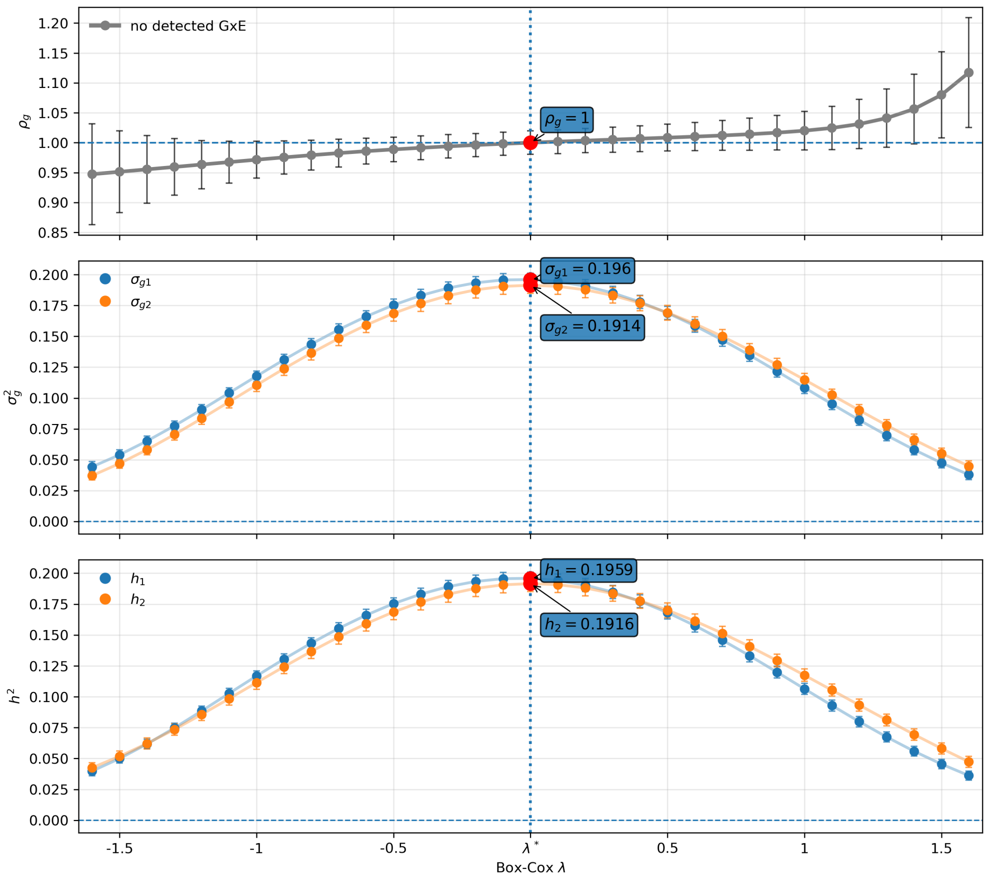

Figure S2: **REGENA estimates across the Box–Cox scan under the null.** Phenotypes were simulated with no G×E ($\sigma^2_{g1} = \sigma^2_{g2} = 0.2$, $\rho_g = 1$) on the log scale ($\lambda = 0$) and then transformed by Box–Cox with $\lambda$ ranging from $-1.5$ to 1.5. REGENA was applied at each grid point. **Top:** estimated cross-environment genetic correlation $\hat{\rho}_g$. **Middle:** estimated environment-specific genetic variances $\hat{\sigma}^2_g$. **Bottom:** estimated environment-specific heritabilities $\hat{h}^2$. Red points mark the optimal $\lambda^*$; error bars denote standard errors.

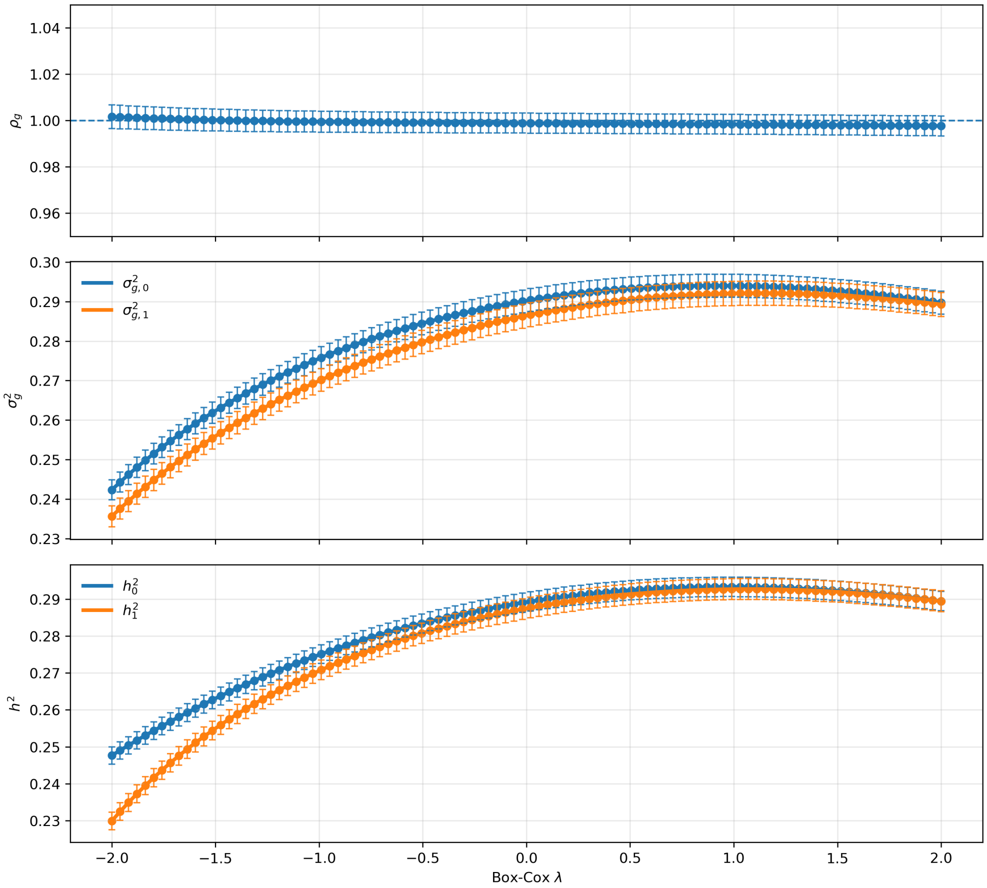

Figure S3: **Stability of REGENA estimates across random seeds.** Phenotypes were simulated under the null with $\sigma^2_{g1} = \sigma^2_{g2} = 0.3$ and $\rho_g = 1$, and REGENA was applied across a Box–Cox grid of $\lambda \in [-2, 2]$. Each panel shows the mean estimate (point) and standard error across 100 random seeds (error bars) of $\hat{\rho}_g$, the environment-specific genetic variances, and the environment-specific heritabilities.

#### Simulations: transformations beyond the Box–Cox family

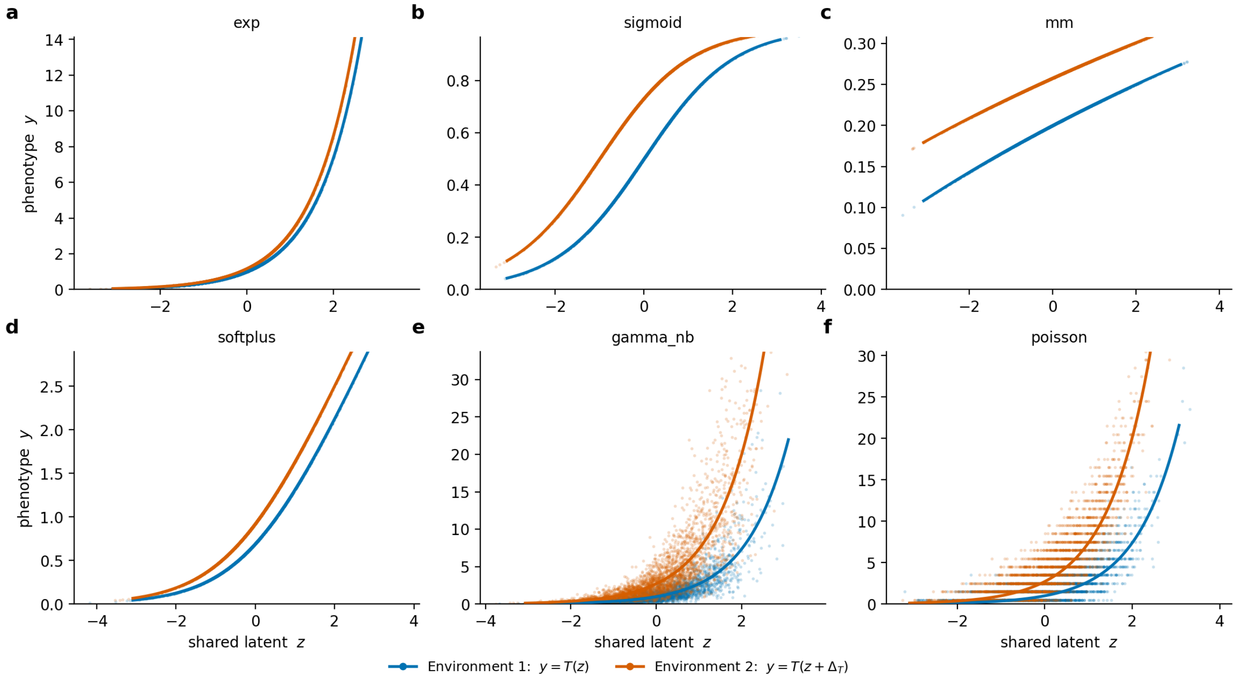

Figure S4: **Construction of the transformed phenotypes.** Observed phenotype $y$ against the shared latent phenotype $z$, which is identically distributed in the two environments. Writing $u = z + \Delta_T E$, the transformations are **a**, $y = e^u$; **b**, $y = 1/(1 + e^{-u})$; **c**, $y = x/(K + x)$ with $x = u + 6$ and $K = 24$; **d**, $y = \log(1 + e^u)$; **e**, $y \sim$ Gamma with shape $k = 4$ and mean $\mu = e^u$; **f**, $y \sim$ Poisson$(\mu) + 0.5$ with $\mu = e^u$ (environment offsets $\Delta_T = 0.155$, 1, 2.33, 0.417, 1 and 1, respectively). Lines show the transformation applied in each environment, $T(z)$ in environment 1 (blue) and $T(z + \Delta_T)$ in environment 2 (orange); in **e** and **f**, where $y$ is a stochastic draw, they show the conditional mean $\mathbb{E}[y \mid z]$. Points are 3,000 individuals sampled per environment from one replicate.

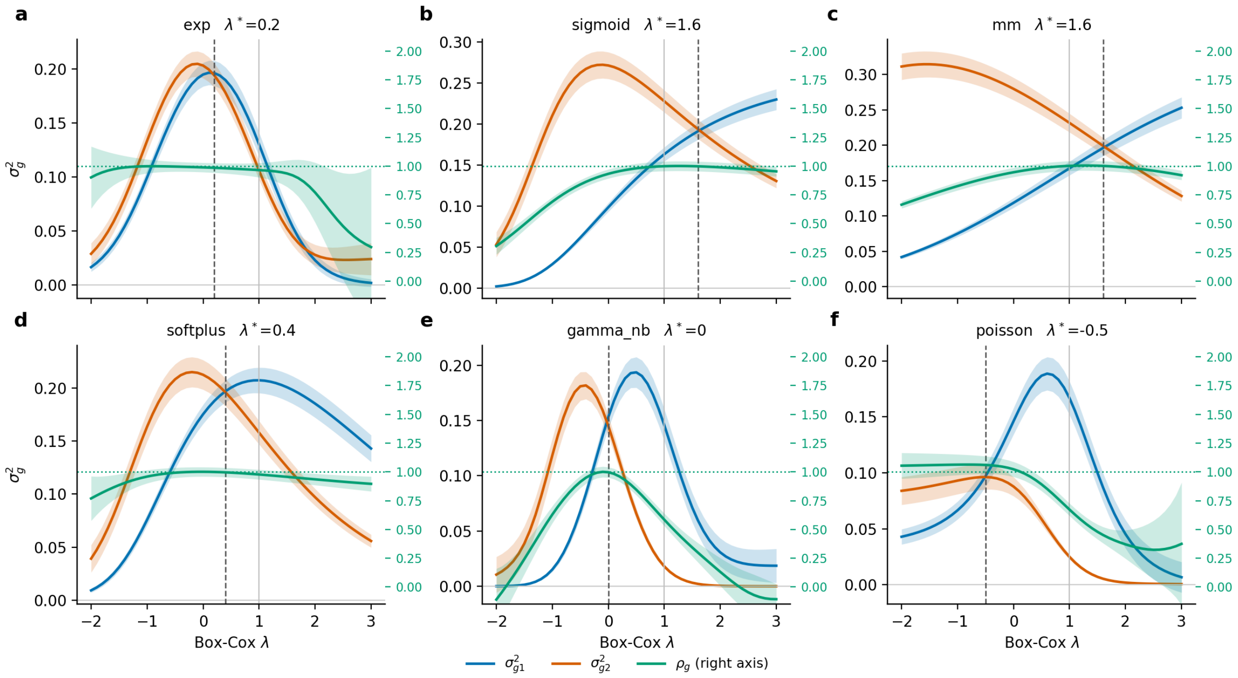

Figure S5: **Genetic variance components across the Box–Cox scan.** REGENA estimates of $\sigma^2_{g1}$ (blue) and $\sigma^2_{g2}$ (orange, left axis) and of $\rho_g$ (green, right axis) at each of the 51 scan points $\lambda \in [-2, 3]$, for one replicate of each transformation. Transformations and environment offsets $\Delta_T$ are as in Figure S4: **a**, exponential; **b**, logistic; **c**, Michaelis–Menten; **d**, softplus; **e**, Gamma; **f**, Poisson. Bands are 95% confidence intervals (±1.96 SE). Vertical lines mark the observed scale $\lambda = 1$ (solid grey) and the reconstructed scale $\lambda^*$ (dashed; given in each panel title); the dotted green line marks $\rho_g = 1$.

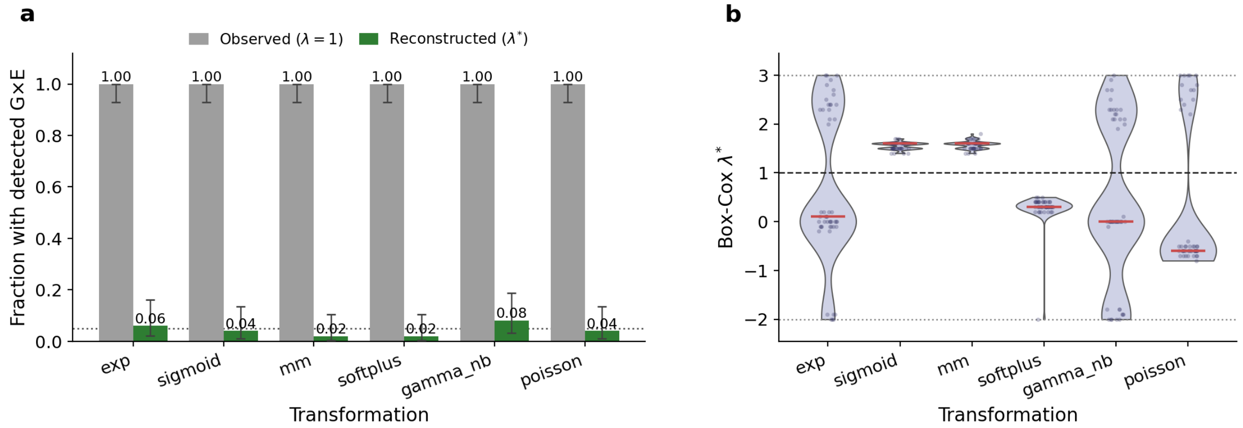

Figure S6: **Detection of G×E and the reconstructed scales.** **a**, Fraction of the 50 replicates in which G×E is detected for each transformation, at the observed scale $\lambda = 1$ (grey) and at the reconstructed scale $\lambda^*$ (green). A replicate is counted as detected if $p < 0.025$ for either $H_0 : \rho_g = 1$ or $H_0 : \sigma^2_{g1} = \sigma^2_{g2}$. Error bars are 95% binomial confidence intervals; the dotted line marks $\alpha = 0.05$. **b**, Distribution of $\lambda^*$ across replicates, shown as a violin with individual replicates jittered over it and the median in red; the dashed line marks $\lambda = 1$ and the dotted lines the scan bounds.

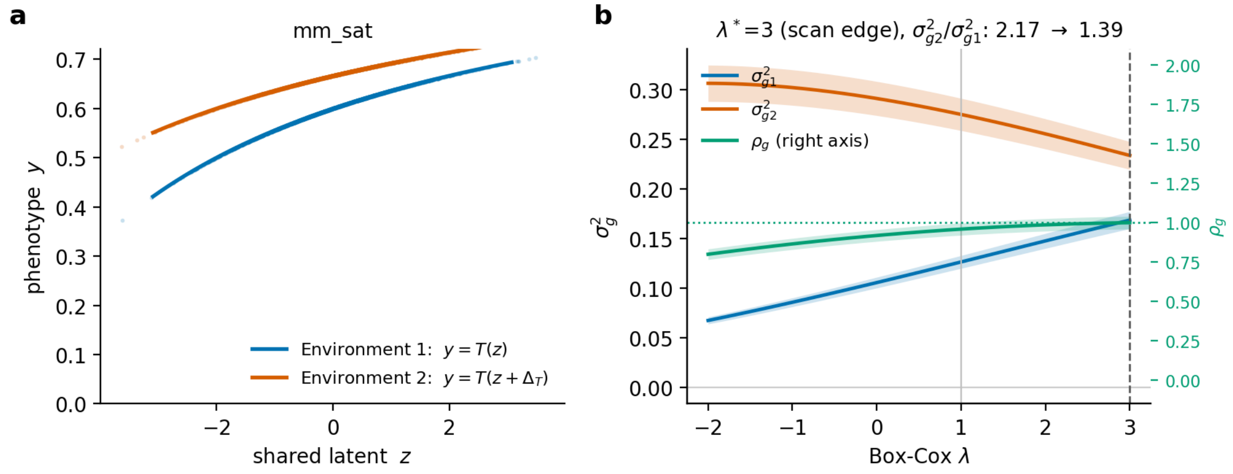

Figure S7: **Scale-induced G×E that the Box–Cox scan does not remove.** The transformation is $y = x/(K + x)$ with $x = u + 6$, $K = 4$ and $u = z + \Delta_T E$, $\Delta_T = 2$; relative to Figure S4**c**, the smaller $K$ and larger offset place environment 2 further onto the plateau of the map. **a**, Observed phenotype against the shared latent phenotype, with lines showing the transformation in environment 1 (blue) and environment 2 (orange) and points showing 3,000 individuals sampled per environment from one replicate. **b**, $\sigma^2_{g1}$ (blue), $\sigma^2_{g2}$ (orange, left axis) and $\rho_g$ (green, right axis) across the scan for the same replicate; the dashed line marks $\lambda^*$, which lies at the upper bound of the scan window in every replicate.

#### Simulations: robustness to confounding

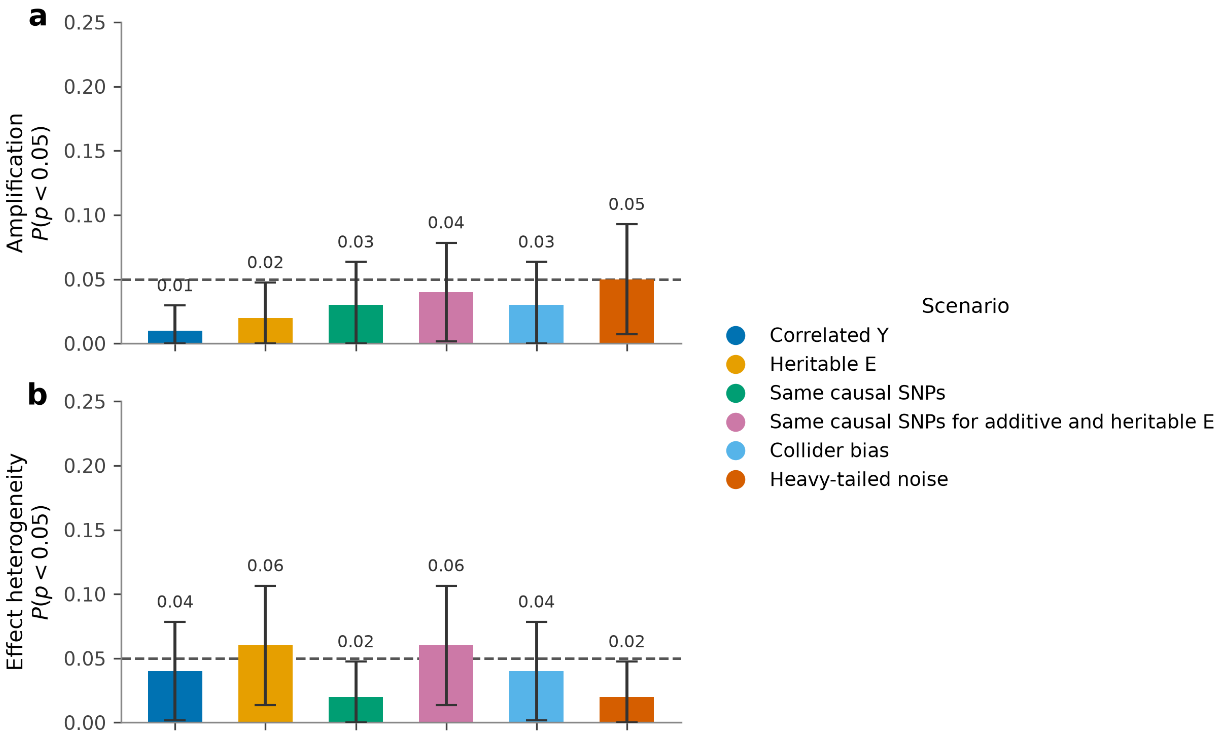

Figure S8: **False-positive rates of the two G×E tests under six models of confounding.** Bars give the fraction of 100 null replicates rejected at $p < 0.05$ (dashed line) for (**a**) the amplification test $p(\sigma^2_{g1} = \sigma^2_{g2})$ and (**b**) the effect-heterogeneity test $p(\rho_g = 1)$, one colour per confounding scenario. Error bars are 95% binomial confidence intervals.

#### Comparison with GENIE

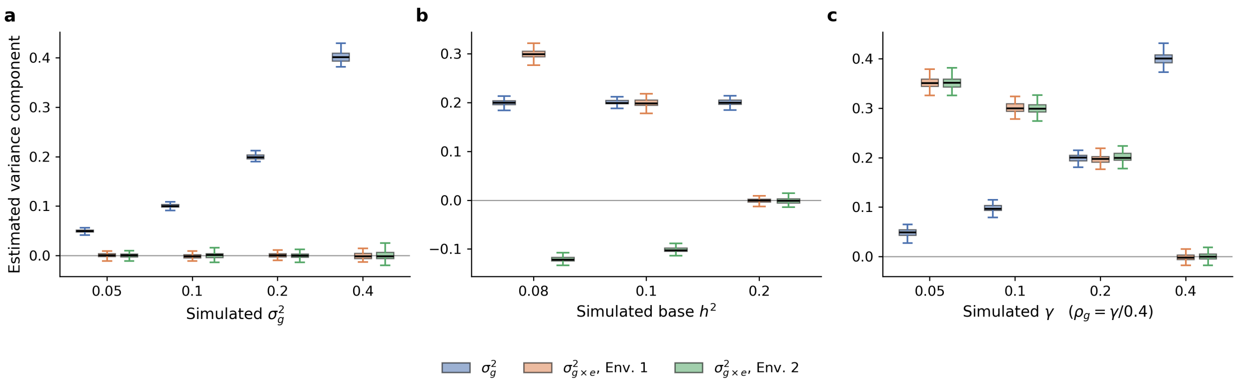

Figure S9: **GENIE captures imperfect genetic correlation but not amplification in fixed-scale simulations.** We simulated phenotypes at a fixed transformation parameter $\lambda$ under three architectures: **a,** a null model; **b,** an amplification model; and **c,** an effect heterogeneity model. Boxplots show estimates of the baseline genetic variance component, $\sigma_g^2$, and two GENIE interaction components across simulation replicates.

#### Mechanisms of G×E across environments

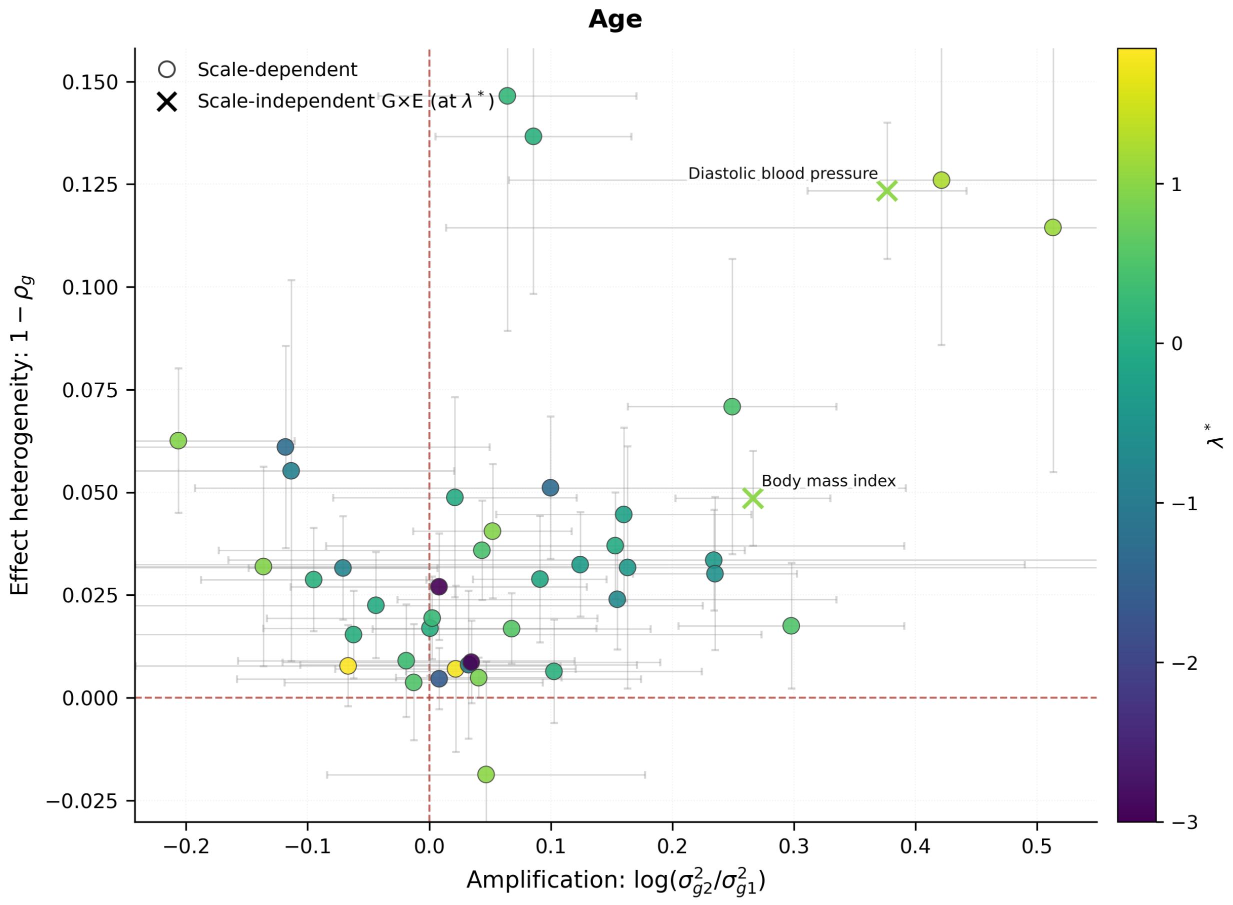

Figure S10: **Mechanisms of gene-by-age interaction across 43 quantitative traits in the UK Biobank.** Age was dichotomized into younger and older strata. REGENA was applied to each trait–age pair with a Box–Cox scan over $\lambda \in [-3, 3]$. Each trait is summarized on two axes evaluated at the optimal $\lambda^*$: effect heterogeneity ($1 - \rho_g$; y-axis) and amplification ($\log(\sigma^2_{g1}/\sigma^2_{g2})$; x-axis). Scale-dependent pairs are shown as dots and scale-independent pairs as markers. Colors indicate the selected $\lambda$. Points near the origin indicate weaker G×E, whereas departures along the x- or y-axis indicate amplification- or heterogeneity-type G×E, respectively.

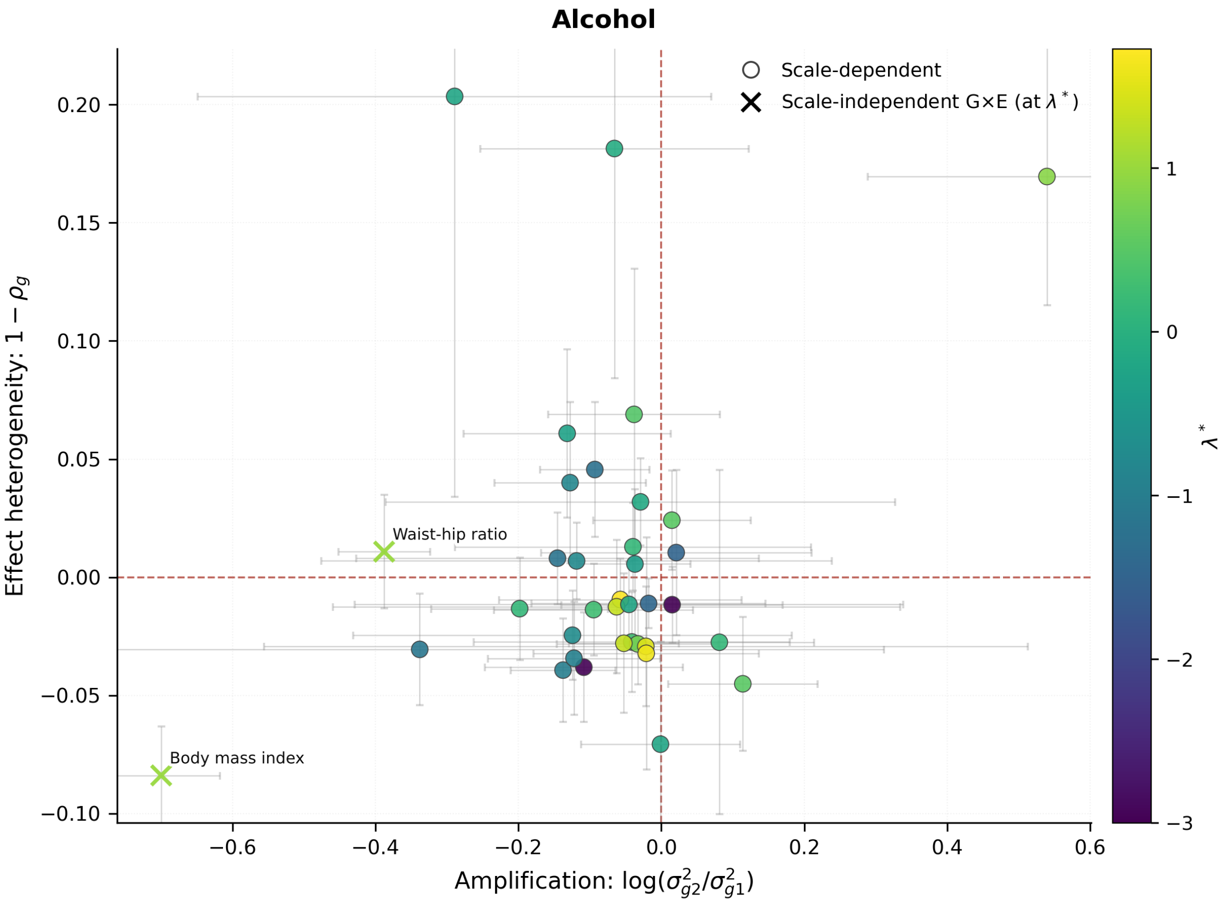

Figure S11: **Mechanisms of gene-by-alcohol-consumption interaction across 43 quantitative traits in the UK Biobank.** REGENA was applied to each trait–alcohol pair with a Box–Cox scan over $\lambda \in [-3, 3]$. Each trait is summarized on two axes evaluated at the optimal $\lambda^*$: effect heterogeneity ($1 - \rho_g$; y-axis) and amplification ($\log(\sigma^2_{g1}/\sigma^2_{g2})$; x-axis). Scale-dependent pairs are shown as dots and scale-independent pairs as markers. Colors indicate the selected $\lambda$.

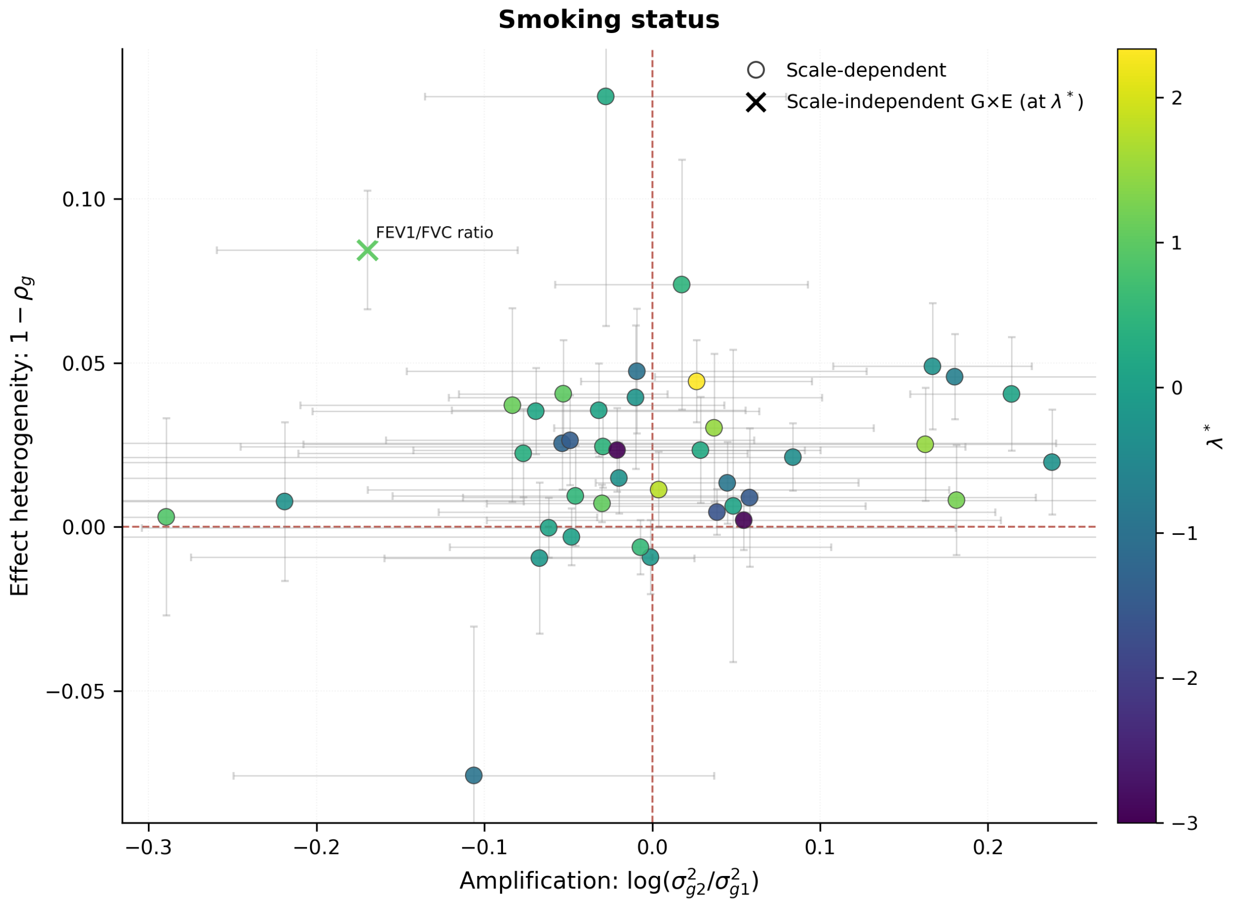

Figure S12: **Mechanisms of gene-by-smoking-status interaction across 43 quantitative traits in the UK Biobank.** REGENA was applied to each trait–smoking-status pair with a Box–Cox scan over $\lambda \in [-3, 3]$. Each trait is summarized on two axes evaluated at the optimal $\lambda^*$: effect heterogeneity ($1 - \rho_g$; y-axis) and amplification ($\log(\sigma^2_{g1}/\sigma^2_{g2})$; x-axis). Scale-dependent pairs are shown as dots and scale-independent pairs as markers. Colors indicate the selected $\lambda$.

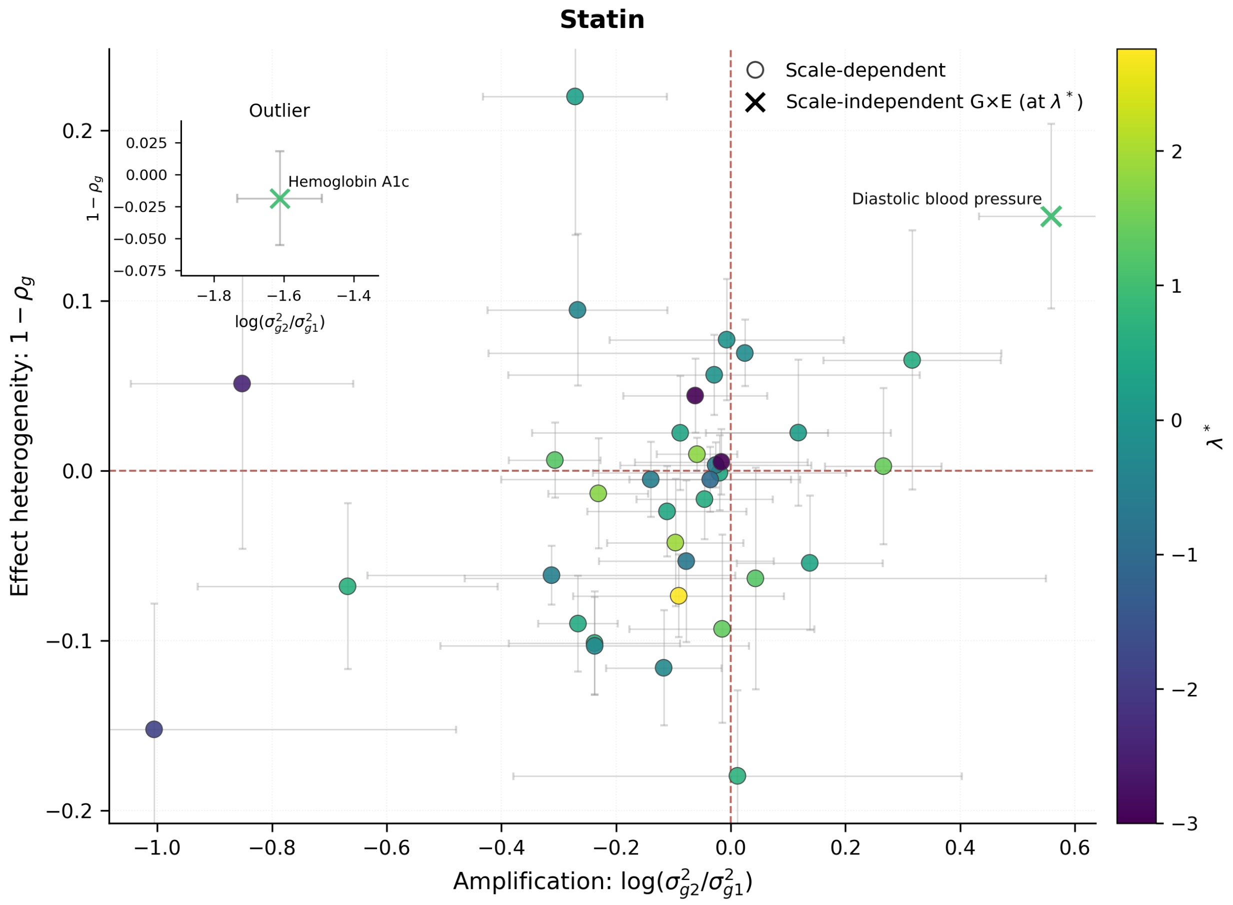

Figure S13: **Mechanisms of gene-by-statin-use interaction across 43 quantitative traits in the UK Biobank.** REGENA was applied to each trait–statin-use pair with a Box–Cox scan over $\lambda \in [-3, 3]$. Each trait is summarized on two axes evaluated at the optimal $\lambda^*$: effect heterogeneity ($1-\rho_g$; y-axis) and amplification ($\log(\sigma^2_{g1}/\sigma^2_{g2})$; x-axis). Scale-dependent pairs are shown as dots and scale-independent pairs as markers. Colors indicate the selected $\lambda$.

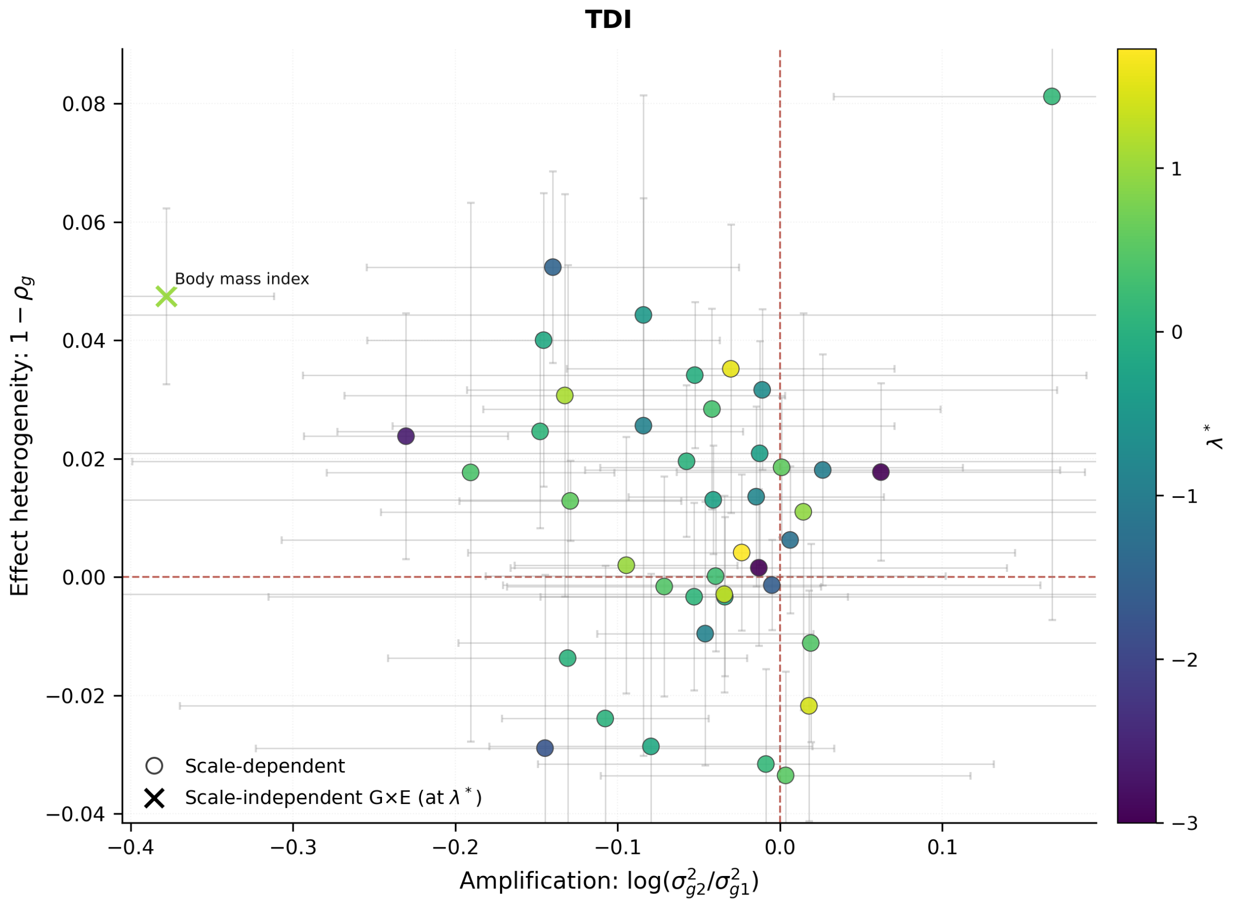

Figure S14: **Mechanisms of gene-by-Townsend-deprivation-index interaction across 43 quantitative traits in the UK Biobank.** The Townsend deprivation index (TDI) was dichotomized into low- and high-deprivation strata. REGENA was applied to each trait–TDI pair with a Box–Cox scan over $\lambda \in [-3, 3]$. Each trait is summarized on two axes evaluated at the optimal $\lambda^*$: effect heterogeneity ($1-\rho_g$; y-axis) and amplification ($\log(\sigma^2_{g1}/\sigma^2_{g2})$; x-axis). Scale-dependent pairs are shown as dots and scale-independent pairs as markers. Colors indicate the selected $\lambda$.

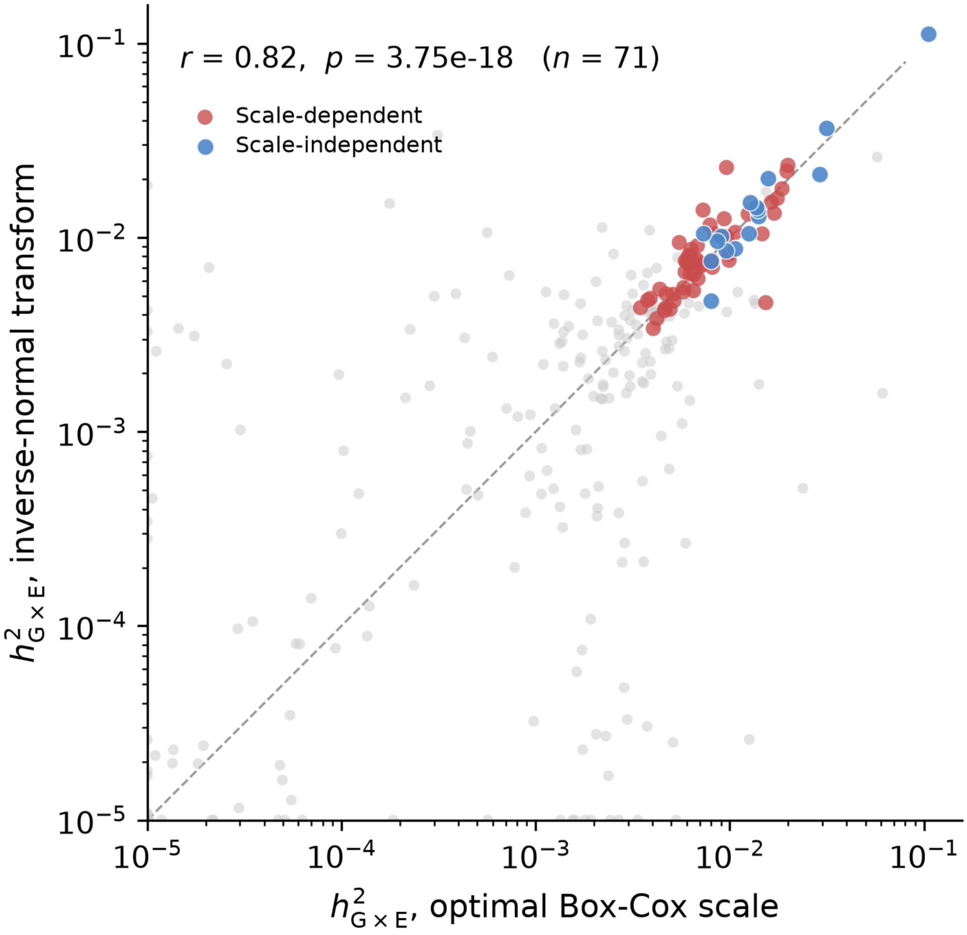

Figure S15: **Concordance of $h^2_{\mathrm{G\times E}}$ across the working scale and INT.** Per-pair $h^2_{\mathrm{G\times E}}$ after a rank-based inverse-normal transform (INT; $y$) against the estimate at the pair's working scale ($x$), on log–log axes; the dashed line is $y = x$. Red, scale-dependent trait–environment pairs that are marginally significant ($p < 0.05$) under both transforms; blue, scale-independent pairs meeting the same criterion. Pearson's $r$ over these pairs is annotated on the panel.

#### MAF–LD partitioned G×E heritability and specificity

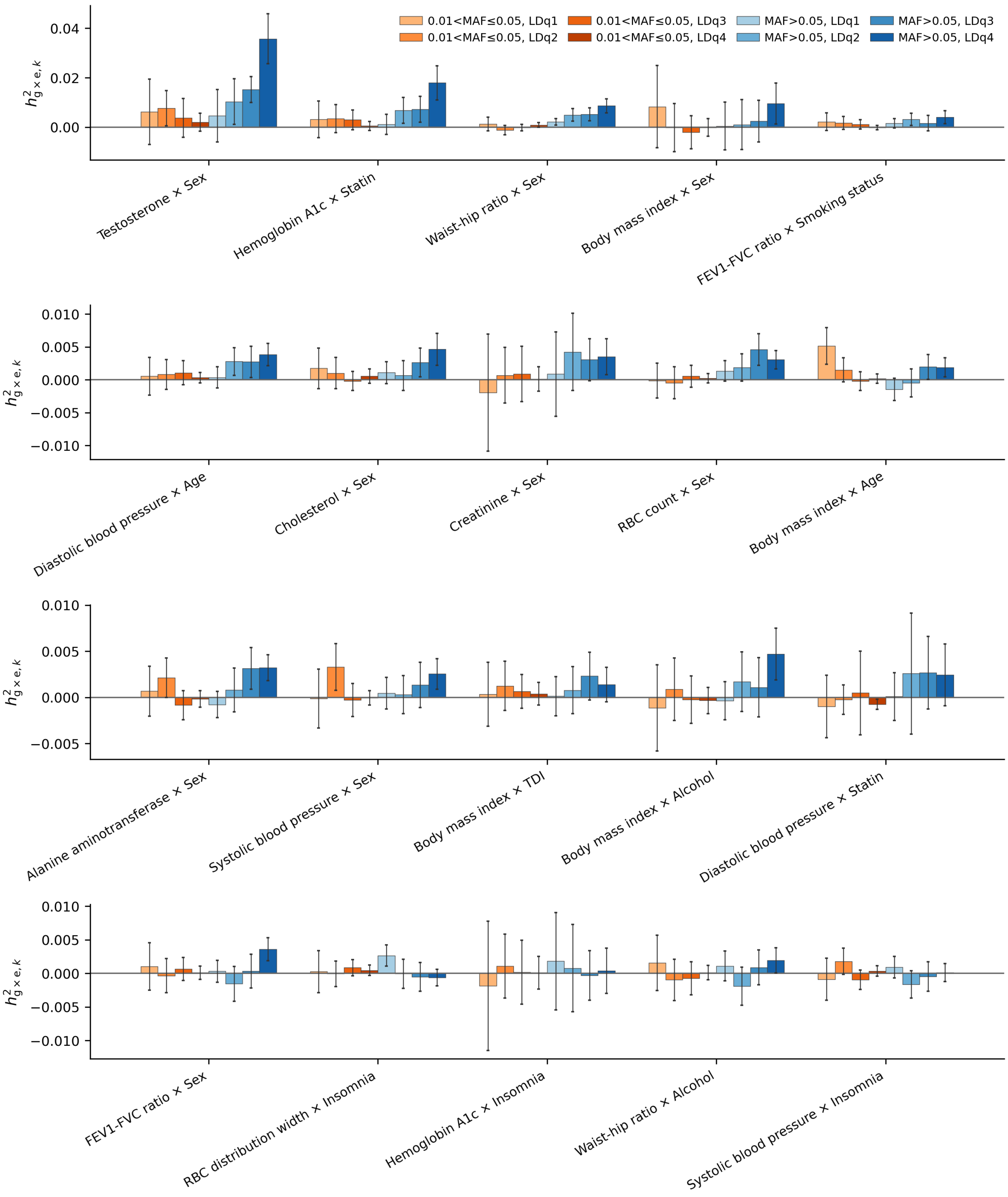

Figure S16: **G×E heritability across MAF and LD bins.** Per-bin G×E heritability for 20 trait-environment pairs with scale-independent G×E. Bins are eight array MAF by LD-quartile bins.

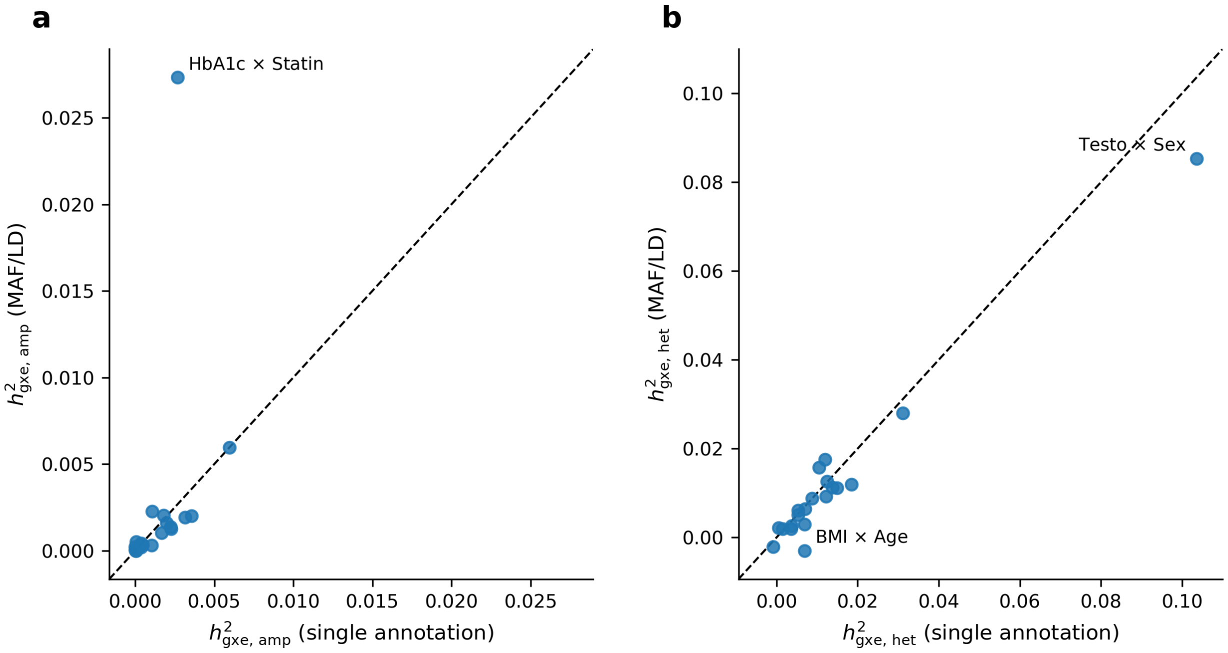

Figure S17: **Concordance of G×E heritability between single-annotation and MAF/LD-partitioned models.** The G×E heritability splits into an amplification and a heterogeneity term, $h^2_{G\times E} = w(1-w)(\sigma_{g1}-\sigma_{g2})^2 + 2w(1-w)\sigma_{g1}\sigma_{g2}(1-\rho_g)$. We compared the estimates of **a**, amplification and **b**, heterogeneity across the two models, for the 20 trait–environment pairs with scale-independent G×E. The $x$ axis is the single-annotation fit and the $y$ axis the eight-bin MAF × LD fit, summing $\sigma^2_{g1}$, $\sigma^2_{g2}$ and $\gamma_g$ over bins before splitting into the two terms. Dashed lines are the identity; pairs falling furthest from it are labeled.

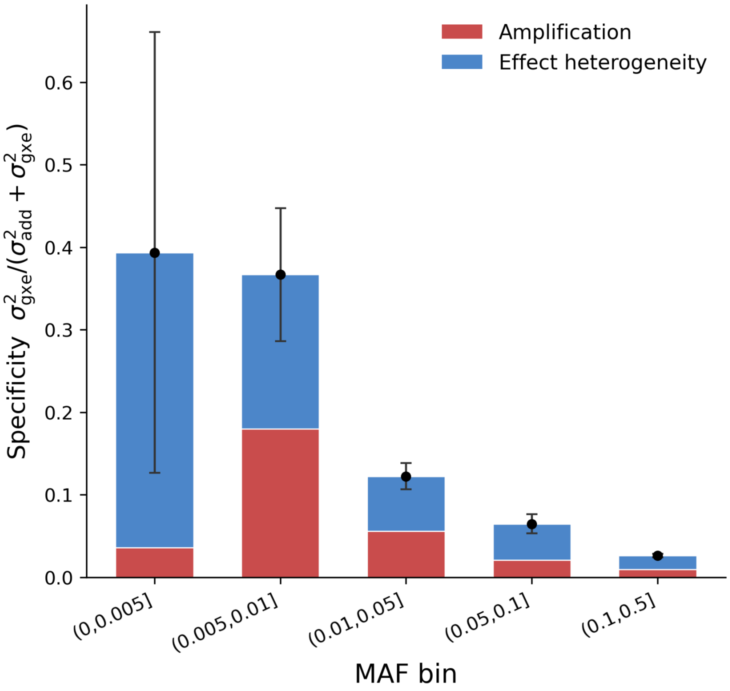

Figure S18: **Decomposition of G×E specificity on the observed scale.** MAF-stratified specificity of G×E heritability, meta-analyzed across all trait–environment pairs with specificity evaluated on the observed scale ($\lambda = 1$) for every pair. Stacked bars show the contributions of amplification specificity, $S_{\text{amp}}$, and effect-heterogeneity specificity, $S_{\text{het}}$. Points denote the total specificity estimates and error bars indicate 95% confidence intervals.

#### G×E polygenic scores

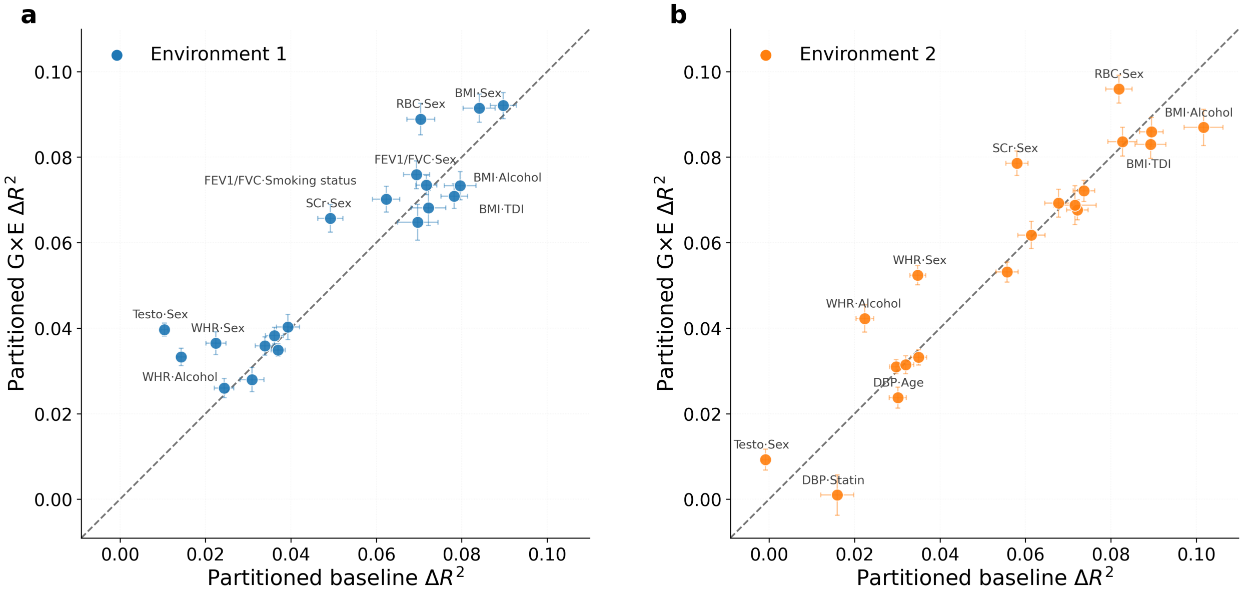

Figure S19: **Environment-specific versus baseline polygenic score prediction within each environmental stratum.** Both panels use the 20 scale-independent trait–environment pairs and compare a MAF/LD-partitioned G×E score with a non-G×E baseline that pools the environmental strata. Incremental $R^2$ ($\Delta R^2$) is the out-of-sample gain over a covariates-only model (sex, 20 genetic PCs, environment indicator), evaluated within a single stratum; both scores share the covariate set and the train/test split. The error bar denotes one standard error from a nonparametric bootstrap over held-out individuals; the panels share axis limits. **a,** Environment 1. **b,** Environment 2. The dashed line is $y = x$; points above it indicate pairs where the G×E score achieves higher $\Delta R^2$ than the baseline.

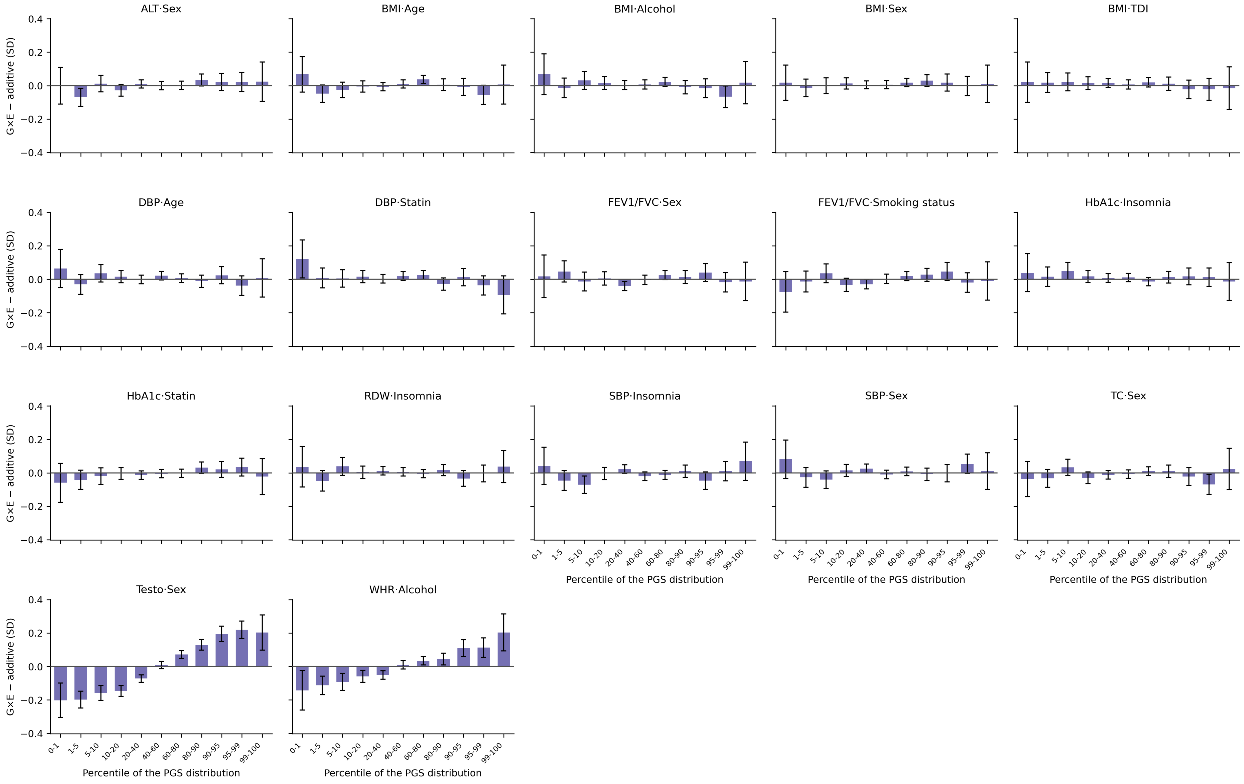

Figure S20: **Difference in covariate-adjusted phenotype between G×E and additive polygenic scores across the score distribution.** One panel per scale-independent trait–environment pair, with waist-hip ratio by sex, creatinine in urine by sex, and RBC count by sex not shown. Held-out individuals were binned by percentile of each score's own distribution. Bars give the mean covariate-adjusted phenotype under the G×E score minus that under the additive score in each bin, in phenotype SD units. Error bars are 95% confidence intervals within the pair.

#### Environment-stratified GWAS

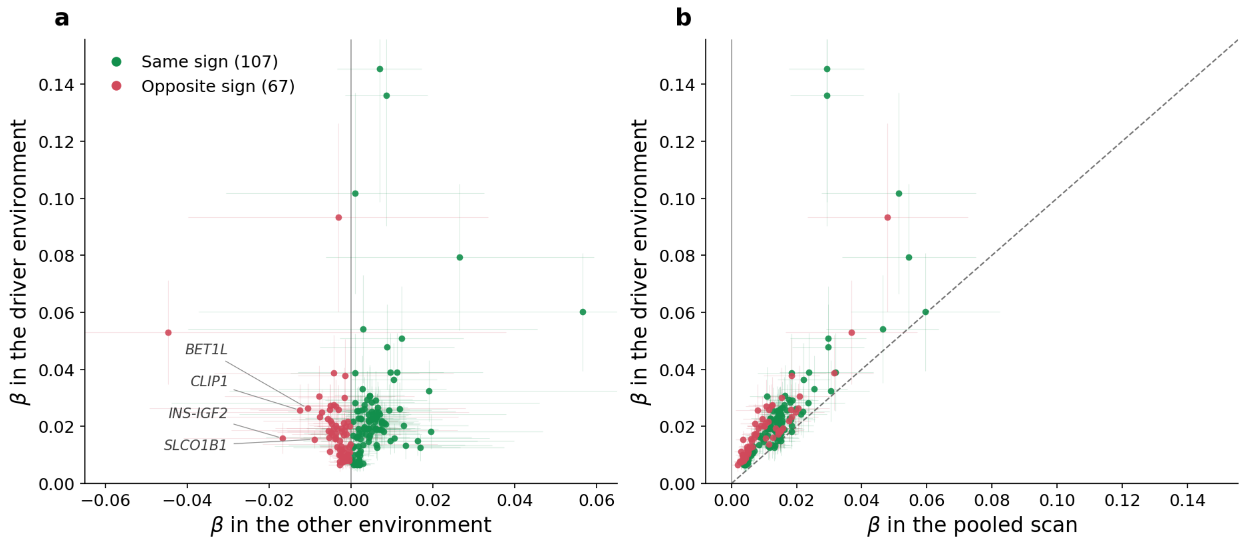

Figure S21: **Effect sizes of the loci recovered only by environment-stratified GWAS.** Each point is one of the 174 novel independent loci found across the 20 trait–environment pairs with scale-independent G×E. The environment in which a locus was significant is its driver environment; effects are expressed in pooled phenotype standard deviations and oriented so that the driver-environment effect is positive. Error-bars are 95% CIs on both axes. **a**, effect in the other environment against effect in the driver environment. Points left of the vertical line have effects of opposite sign in the two environments (red, $n = 67$); points to its right share the same sign (green, $n = 107$). The four opposite-sign loci with the strongest reversal are named. **b**, effect in the pooled scan against effect in the driver environment, with the dashed identity line. Nearly all loci lie above the identity: pooling the two environments dilutes the effect, most severely for the opposite-sign loci, whose strata partially cancel and which cluster closest to zero on the pooled axis.

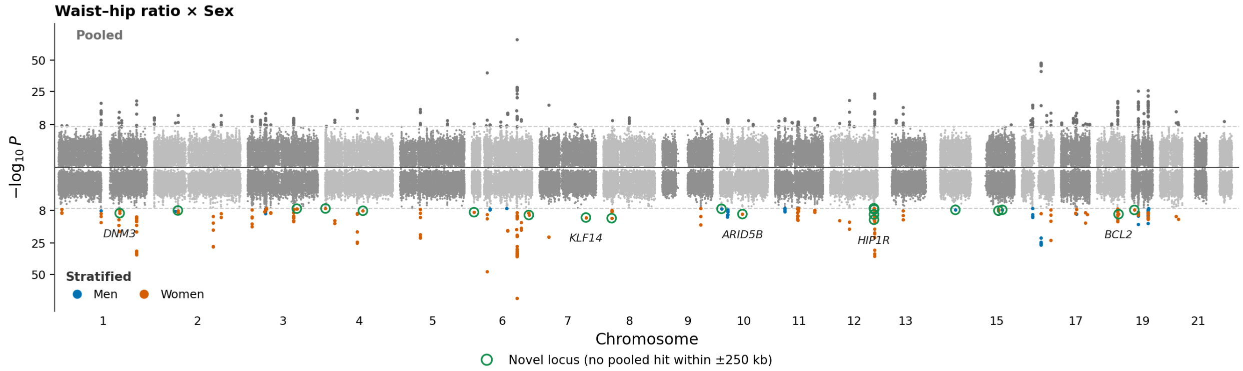

Figure S22: **Pooled and sex-stratified GWAS of waist–hip ratio.** Mirror Manhattan plot for waist–hip ratio × sex at its optimal scale ($\lambda^* = 1.85$): the pooled scan above the axis, the sex-stratified scan below, each variant plotted at the smaller of its two within-environment $p$-values and colored by the driving sex. Dashed lines mark $p = 5 \times 10^{-8}$. Open green circles mark the 21 novel loci.

#### Categorical environment encoding

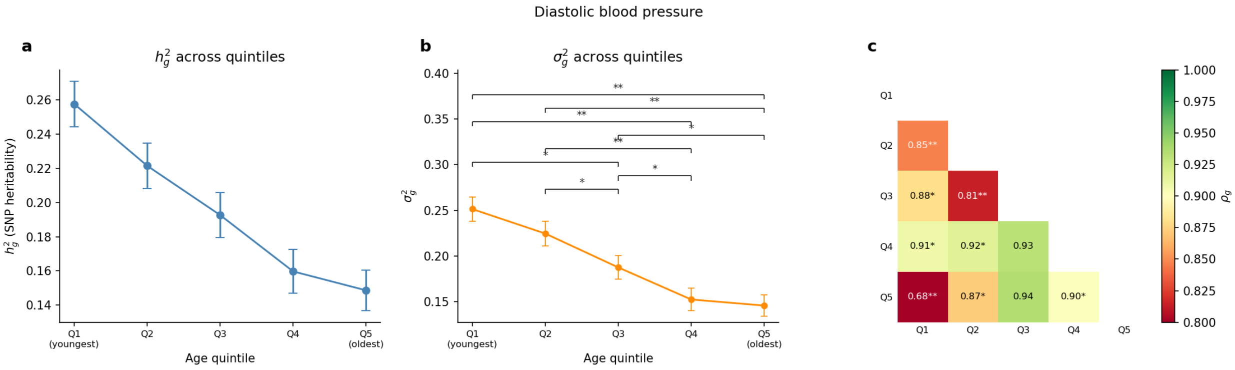

Figure S23: **Gene-by-age interaction for diastolic blood pressure under a five-stratum categorical encoding of age.** Age was discretized into quintiles (Q1, youngest, to Q5, oldest), and REGENA was applied to diastolic blood pressure across the five strata. **a,** environment-specific SNP heritability $\hat{h}_g^2$. **b,** environment-specific genetic variances $\hat{\sigma}_g^2$. Brackets indicate pairwise tests of variance equality. **c,** matrix of pairwise cross-environment genetic correlations $\hat{\rho}_g$, with point estimates shown in each cell. Single ($*$) and double ($**$) asterisks denote marginal ($p < 0.05$) and Bonferroni-significant ($p < 0.05/(43 \times 7)$) tests against $H_0 : \sigma_{g,i}^2 = \sigma_{g,j}^2$ (middle) or $H_0 : \rho_g = 1$ (right). Error bars denote standard errors.

### Supplementary Data

**Data S1. G×E variance components for all trait–environment pairs.** Environment-specific genetic variances, cross-environment genetic correlation, and Wald tests of $H_0: \sigma^2_{g1} = \sigma^2_{g2}$ and $H_0: \rho_g = 1$ at the reporting scale $\lambda^*$, with the scale classification from the Box–Cox scan.

**Data S2. Novel context-specific loci from environment-stratified GWAS.** Lead SNP, nearest gene, stratum-specific effect sizes, standard errors and $p$ values, the smallest pooled-scan $p$ value within $\pm 250$ kb, and whether effects have opposite signs across strata.
